# Rare variant effect estimation and polygenic risk prediction

**DOI:** 10.1101/2024.06.23.24309366

**Authors:** Kisung Nam, Minjung Kho, Wei Zhou, Bhramar Mukherjee, Seunggeun Lee

## Abstract

Due to their low frequency, estimating the effect of rare variants is challenging. Here, we propose RareEffect, a method that first estimates gene- or region-based heritability and then each variant effect size using an empirical Bayes approach. Our method uses a variance component model, popular in rare variant tests, and is designed to provide two levels of effect sizes, gene/region-level and variant-level, which can provide better interpretation. To adjust for the case-control imbalance in phenotypes, our approach uses a fast implementation of the Firth bias correction. We demonstrate the accuracy and computational efficiency of our method through extensive simulations and the analysis of UK Biobank whole exome sequencing data for 100 traits. Additionally, we show that the effect sizes obtained from our model can be leveraged to improve the performance of polygenic scores, outperforming recently developed methods for rare variant polygenic scoring.

## Introduction

With the availability of extensive sequencing data in biobanks^1^, the study of rare variants has become more feasible than ever before. Rare variants have been identified as potential causative factors for numerous complex diseases^2-8^, and their exploration is crucial in unraveling the genetic risk factors of complex traits^9^. To identify the association between rare variants and complex traits, gene or region-based tests^10,11^, including the Burden test, the sequence kernel association test (SKAT)^12^ and its adaptive optimized version (SKAT-O)^13^, have been commonly used. Recently, several methods, including STAAR^14^, SAIGE-GENE^15^, SAIGE-GENE+^16^, and DeepRVAT^17^ are developed to run region-based tests in biobank scale data.

To elucidate the effect of rare variants on complex diseases and traits and utilize them for risk prediction, in addition to calculating p-values for association tests, effect size estimation is required. However, estimating the effect size of rare variants remains a challenge. The low minor allele frequencies make single variant-based estimations unstable. Popular association tests like SKAT and SKAT-O are score tests, so only provide p-values. Although the Burden test provides a gene-burden effect size, this may not accurately reflect the true effect of rare variants in the presence of null variants and variants with opposing association directions. Recently, several methods have been developed to estimate the effect sizes of rare variants, aiming to overcome the limitations of burden test approaches^18-20^. However, these methods often require excessive computational resources, making their application to large-scale datasets impractical. Alternative approaches estimate burden effect sizes by weighting variants according to their predicted functional impact or by directly computing gene impairment scores from predicted variant functionality^17,21,22^. However, as variant function prediction remains an evolving field, such direct weighting strategies may be prone to reduced accuracy.

To address the challenges, we introduce RareEffect, a novel method that estimates gene or region-based heritability and subsequently calculates each variant’s effect size using an empirical Bayes approach. Utilizing a variance component model, similar to that in the SKAT test, our method offers dual-level effect size estimation, region-level and variant-level, for enhanced interpretability. Unlike the Burden approach, our model flexibly estimates effect sizes in a data-driven manner. To reduce the computational burden for estimating variance components, we implemented the Factored Spectrally Transformed Linear Mixed Models (FaST-LMM)^23^, which leverages the low rank of the genetic relatedness matrix. We also utilize the strategy to collapse ultra-rare variants, as used in SAIGE-GENE+, to reduce the sparsity of the genotype matrix and improve power of estimating the effect of ultra-rare variants^16^. For binary traits, we additionally apply a fast implementation of the Firth bias reduction method to stably estimate the effect sizes.

From simulation studies, we showed that the proposed method is computationally fast and reliably estimates each gene heritability. We also showed that the proposed approach outperformed linear regression or ridge regression in terms of root mean squared error (RMSE) for estimating the individual-variant level effect sizes. From the UK Biobank (UKB) Whole Exome Sequencing (WES) data analysis of 100 traits, we demonstrate that exonic rare variants can explain additional phenotypic variabilities, but the degree differs by phenotypes. We also demonstrated that our approach explains phenotypic variability more effectively than recently developed methods based on predicted variant functional impact or gene-impairment scores, such as AlphaMissense^21^, PrimateAI-3D^22^ and DeepRVAT^17^. These findings provide strong evidence for the practical utility of our method in leveraging rare variant data for risk prediction and heritability estimation.

## Results

### Overview of methods

Our proposed method encompasses three steps. The overview of the method is outlined in **Fig. 1** and is described in detail in the Methods section.

**Fig. 1.**
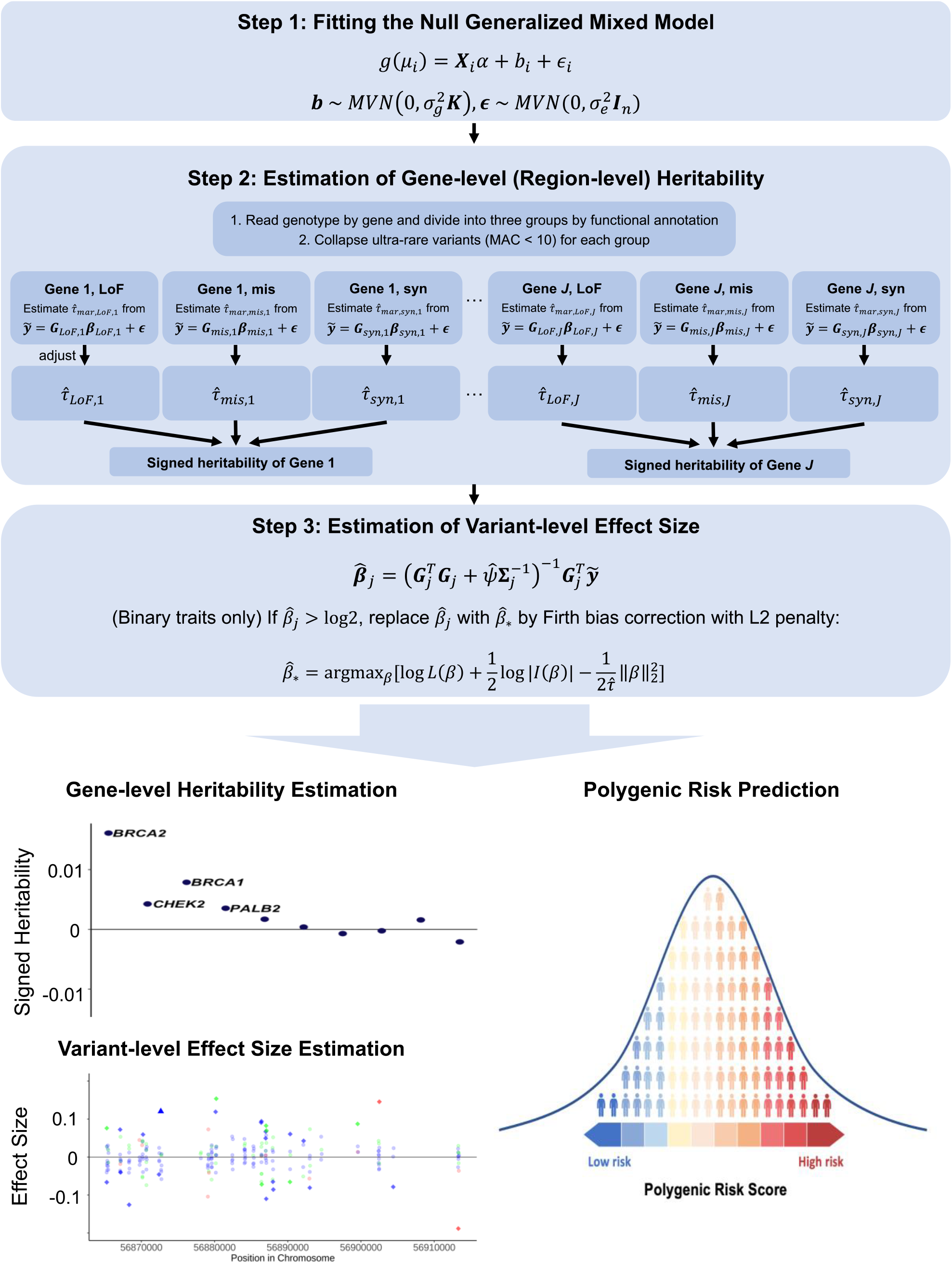
Overview of RareEffect. RareEffect encompasses three steps. In step 1, we fit a null GLMM using AI-REML approach, and obtain residuals for the subsequent steps. In step 2, we divide variants by gene and its functional annotation (LoF, missense, and synonymous). We first estimate the variance component of each group and adjust them using MoM approach. In step 3, we calculate the variant-level effect size using BLUP estimates. For binary traits, Firth bias correction is additionally applied to adjust the case-control imbalance. Through RareEffect, we provide region-level and variant-level effect sizes for enhanced interpretability and improved risk prediction performance.

In step 1, we fit a null linear or logistic mixed-effect model without genotypes to estimate the covariate effects. This step involves fitting the model using the average information restricted maximum likelihood (AI-REML)^24^ approach, which is utilized in the SAIGE^25^ and GMMAT^26^ framework. Residuals will be used as covariate-adjusted phenotypes in the subsequent steps.

In step 2, we model the effect of rare variants (MAF≤0.01) as random effects and estimate the variance components, and hence heritability. To mitigate the computational demands, we adopt the Factored Spectrally Transformed Linear Mixed Models (FaST-LMM)^23^. Given *n* ≫ *k*, FaST-LMM reduces the computation cost from *O*(*n*^3^) of conventional mixed model algorithm to *O*(*nk*^2^) where *n* is the number of samples in the dataset and *k* is the number of genetic variants in a single group (See **Methods**). Additionally, we leveraged the sparsity in genotypes. To incorporate the fact that genetic effects vary by functional annotations, we fit the model separately for distinct categories, and then combine them to calculate gene or region-level heritability. For whole exome analysis, we include three functional categories: (1) Loss-of-function (LoF) variants; (2) missense variants; and (3) synonymous variants. Within each category, the ultra-rare variants, defined as those with a minor allele count (MAC) of lower than 10, are collapsed into a single variant, as employed in SAIGE-GENE+^16^. As estimating multiple variance components due to distinct functional groups is not feasible with FaST-LMM, we marginally calculate variance component for each functional group separately, and then adjust them using method of moments (MoM) approach (See **Methods**).

In step 3, following the estimation of variance components, we calculate the effect size of each variant using the Best Linear Unbiased Predictor (BLUP) estimates^27-30^. We further estimate the prediction error variance (PEV) of the effect size estimates to assess the reliability of the variant-level effect sizes. For binary traits, we implemented Firth bias correction^31^ as a subsequent step. This correction mechanism mitigates bias and rectifies abnormal estimates, especially in scenarios where the case-control ratio is imbalanced. Recognizing the imperative of scalability in large-scale biobank data analyses, we developed a fast implementation of Firth correction, which reduces computation complexity from *O*(*Mnk*_*F*_) to *O*(*n*_*nz*_*k*_*F*_) where *M*is the average number of iterations for convergence of Firth corrected beta, *n*_*nz*_is the number of individuals with non-zero genotype, and *k*_*F*_ denotes the number of variants that needs to be corrected (See **Methods**).

### UK Biobank WES data analysis

We applied our method to 100 traits in the UKB, including clinically important quantitative traits such as cholesterol levels and major diseases such as type 2 diabetes and various cancers. The full list of analyzed traits is provided in **Supplementary Table 1**. For LDL cholesterol (LDL-C) level, we adjusted the pre-medication levels by dividing the raw LDL-C level by 0.7 for individuals on cholesterol-lowering medication^32^.

We computed gene-level effect sizes by leveraging the estimated heritability derived from the mixed effects model. Recognizing the inherent unsigned nature of heritability, we assign a sign by incorporating variant-level effect sizes of loss-of-function (LoF) variants within each gene. This allows us to discern the direction of the effect of the gene on the trait (See **Methods**). As expected, the gene-based association test p-values and the magnitude of the gene-level effect size showed a substantial correlation (**Fig. 2** and **Supplementary Figure 1**). Incorporating gene-level effect size and direction on top of the gene-based association tests can add significant value to genetic analyses. For example, the signed heritability clearly shows that the impairment of APOC3 function increases HDL cholesterol (HDL-C) level but decreases triglycerides (**Fig. 2(a)** and **2(c)**).

**Fig. 2.**
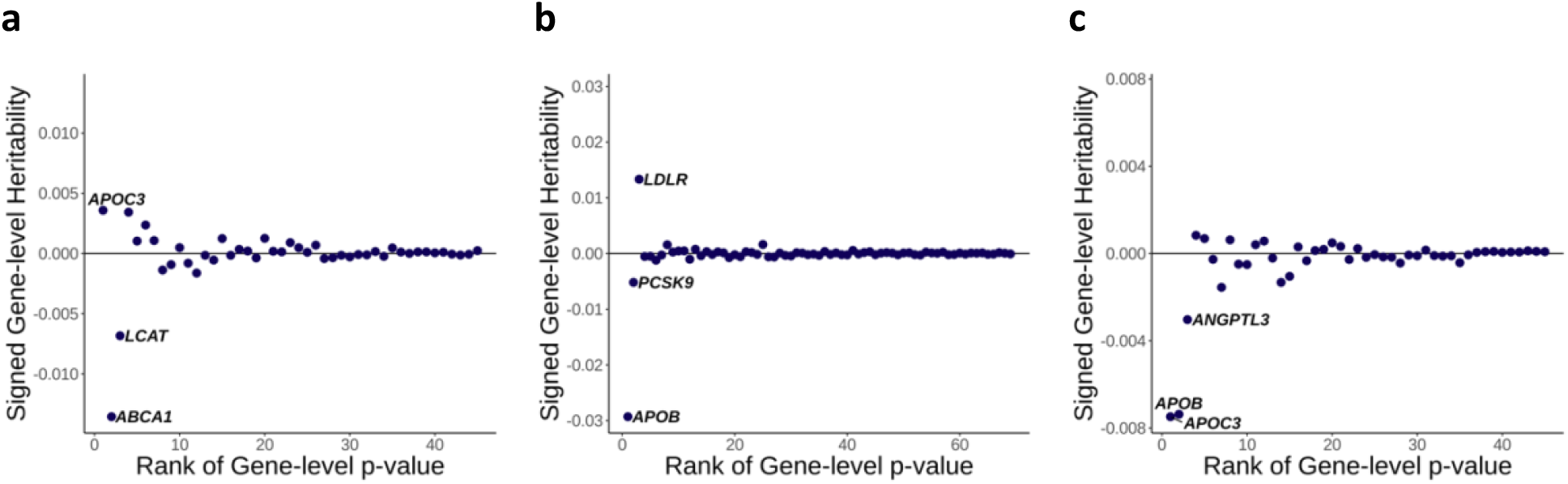
Estimated signed heritability for lipid phenotypes using 392,748 White British samples in UK Biobank whole exome sequencing data. Signed gene-level heritability from RareEffect for (a) HDL cholesterol level, (b) LDL cholesterol level, and (c) triglycerides level. Gene-level p-values were obtained from SAIGE-GENE+, and we included genes with p-values < 2.5 × 10^−6^. The *x*-axis represents the rank order of genes based on their gene-level p-values. Lower ranks correspond to genes with more significant p-values. The *y*-axis shows the signed gene-level heritability for each gene. Signed heritability indicates the direction (positive or negative) and magnitude of the genetic contribution of the gene to the trait.

We estimated the variant-level effect sizes of exonic variants and presented two genes, *APOC3* and *SLC12A3*, on HDL-C levels as examples (**Fig. 3**). As expected, variants demonstrating significant associations, in terms of p-values, also exhibited larger effect sizes. Using RareEffect, we observed that the majority of variants in *APOC3* displayed positive effect sizes. The Burden and SKAT-O p-values from SAIGE-GENE+ were both highly significant (Burden p-value = 1 × 10^−298^ and SKAT-O p-value = 1 × 10^−300^). In contrast, variants in *SLC12A3* exhibited both positive and negative effect sizes. Consequently, Burden p-value was not significant (Burden p-value = 0.01), but the SKAT-O p-value was significant (SKAT-O p-value = 7 × 10^−12^). This distinction in the directionality of effect sizes cannot be discerned through the burden approach.

**Fig. 3.**
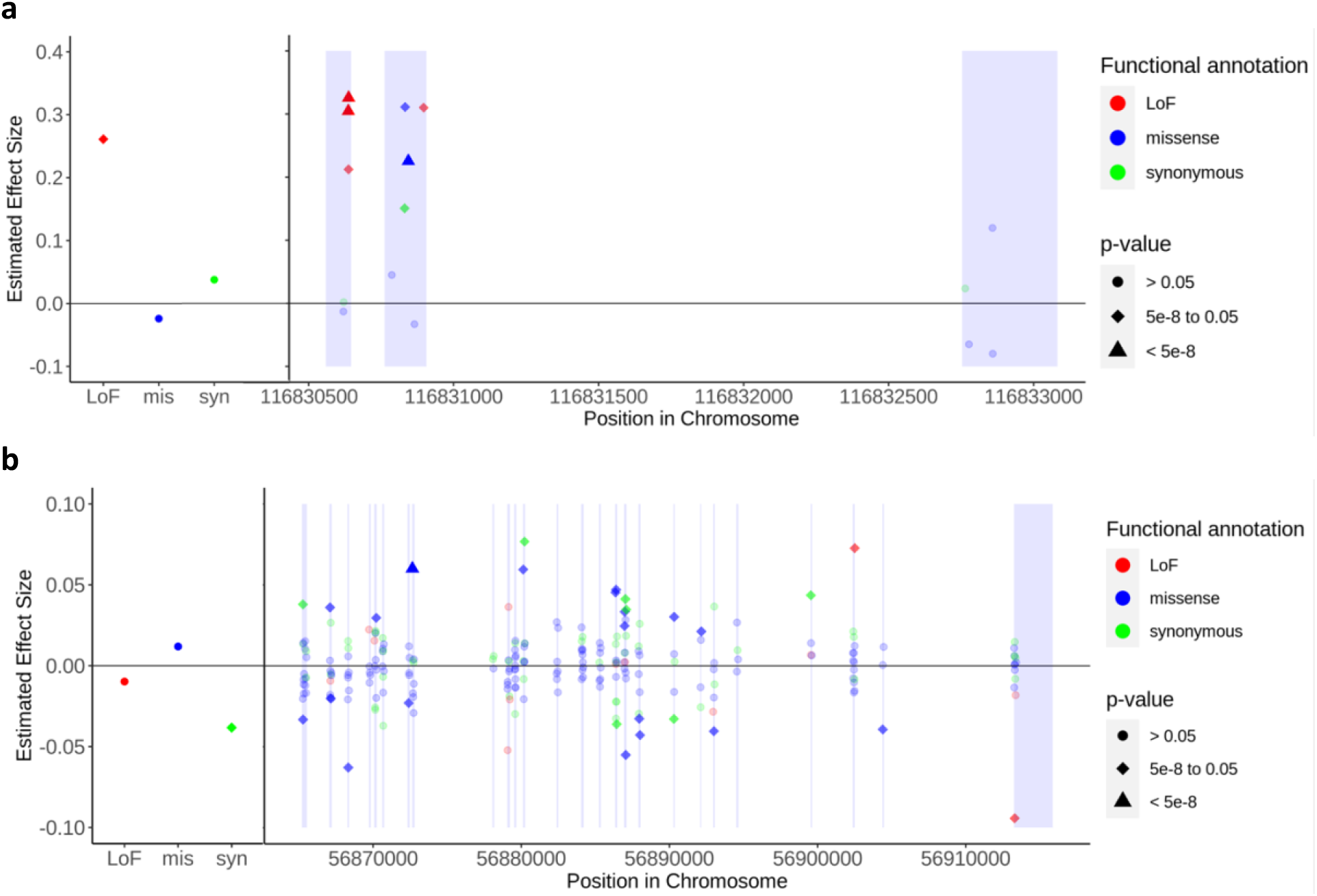
Variant-level effect size on HDL cholesterol for variants in *APOC3* gene (chromosome 11) and *SLC12A3* gene (chromosome 16). Variant-level effect size on HDL cholesterol level for (a) *APOC3* and (b) *SLC12A3* genes. Gene-level p-values were obtained from SAIGE-GENE+, and we included genes with p-values < 2.5 × 10^−6^. The left panel shows the effect size of collapsed ultra-rare variants categorized by their functional annotations: loss-of-function (LoF), missense, and synonymous, respectively. The right panel displays the variant-level effect size of rare variants. Variants are color-coded based on their functional annotation: red for LoF, blue for missense, and green for synonymous. The shapes of the points indicate the significance of the variants: circles represent p-values > 0.05, diamonds represent p-values between 5 × 10^−8^ and 0.05, and triangles represent p-values < 5 × 10^−8^. Single-variant p-values were obtained from SAIGE, while the p-values of collapsed variants were derived from linear regression by regressing 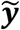 on each collapsed variant. The exon region is shaded.

We extended our analyses to estimate polygenic risk scores using rare variants with effect sizes estimated from RareEffect (*PRS*_*RE*_). The UK Biobank data were randomly split into training and test sets (ratio = 8:2), and *PRS*_*RE*_was constructed using the genes with p-values< 2.5 × 10^−6^ from SAIGE-GENE+ in the training set (See **Methods**). We included top 10 genes when the number of genes with p-values < 2.5 × 10^−6^ is smaller than 10. We further integrated these *PRS*_*RE*_with PRS derived from common variants (*PRS*_*common*_) using SBayesRC^33^ to evaluate the practical utility of our approach. When combining the PRS from common and rare variants, we constructed a composite score, a linear combination of PRS from common and rare variants. We evaluated the predictive performance in terms of *R*^2^.

Our RareEffect PRS (*PRS*_*RE*_) showed higher *R*^2^ for 97% of tested quantitative traits than *PRS*_*burden*_, a comparative approach that relied on per-allele effect sizes derived from burden tests (**Fig. 4, Supplementary Table 2-4**, and **Supplementary Figure 2**). In addition, combining the common variant PRS with the *PRS*_*RE*_ to construct a composite score improved prediction performance in a number of phenotypes (**Supplementary Note A** and **Supplementary Table 2**). The improvement became pronounced for lipid phenotypes among individuals deemed at high risk. For instance, when predicting HDL and LDL cholesterol levels, *PRS*_*RE*_achieved *R*^2^ of 0.4737 and 0.6174 when restricting individuals with top/bottom 0.5% in terms of *PRS*_*RE*_. When the composite score was used for risk prediction, *R*^2^ were 0.6619 and 0.7012 when restricted to top/bottom 0.5% individuals. In contrast, common variants only PRS model had lower *R*^2^ (0.6028 for HDL and 0.6046 for LDL) for top/bottom 0.5% individuals. Notably, the sub-groups identified as high-risk by *PRS*_*common*_ and *PRS*_*RE*_were substantially distinct (**Supplementary Figure 3**), underscoring the complementary nature of rare variants in detecting individuals at elevated disease risk. Additionally, we observed that our model showed higher predictability compared to the Burden approach which showed *R*^2^ of 0.3684 and 0.2362 for HDL and LDL with top/bottom 0.5% in terms of PRS using burden score (*PRS*_*burden*_), respectively. We observed the similar pattern when PRS-CS^34^ was used for *PRS*_*common*_ (**Supplementary Table 2**).

**Fig. 4.**
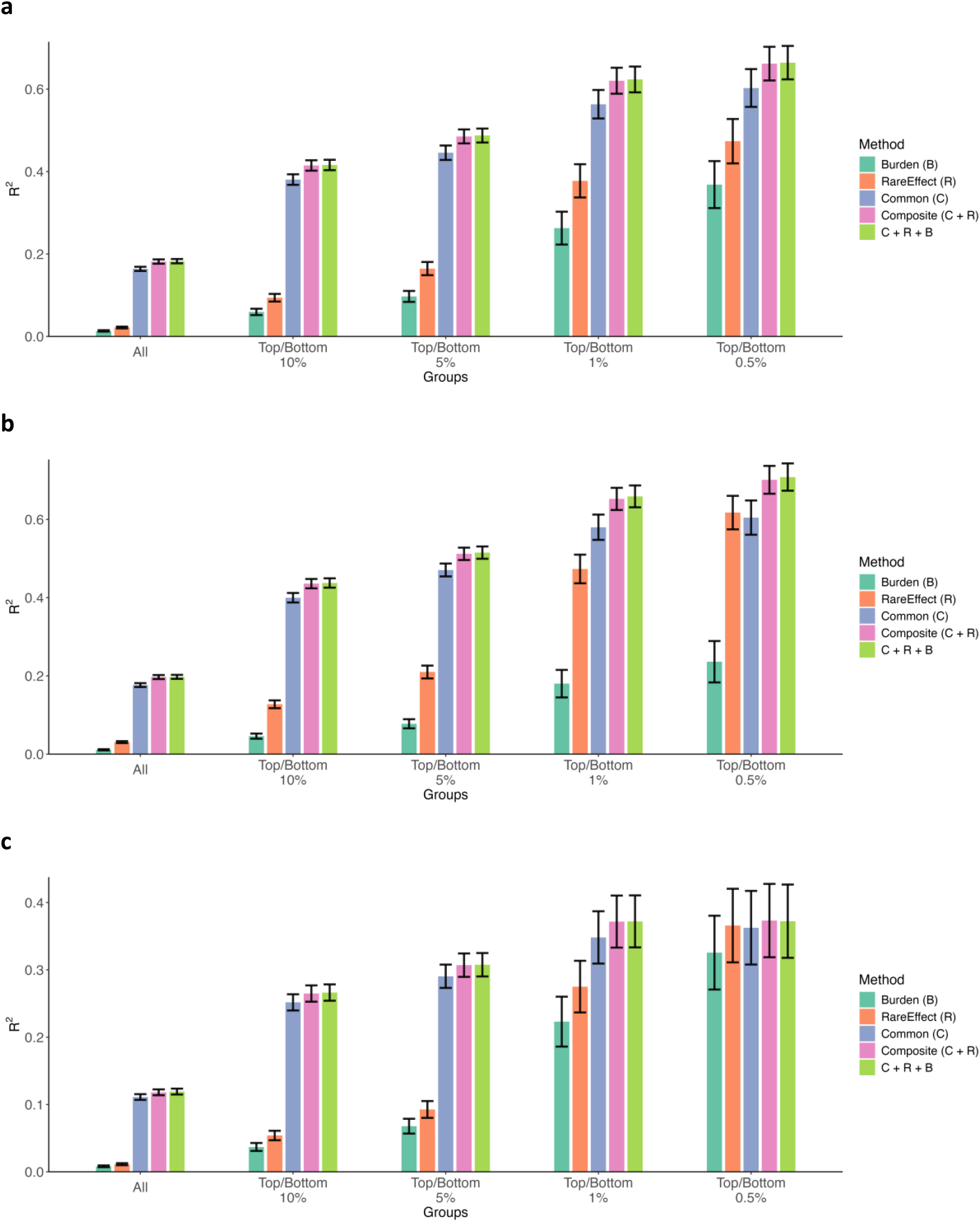
Comparison of performance of risk prediction models for lipid phenotypes using 314,198 White British samples (80% of the whole White British samples) in UK Biobank whole exome sequencing data. Predictive performance was evaluated using the coefficient of determination (*R*^2^) of the risk prediction models for (a) HDL cholesterol level, (b) LDL cholesterol level, and (c) Triglycerides level. We evaluated the performance of five models using: (1) *PRS*_*burden*_only, (2) *PRS*_*RE*_only, (3) *PRS*_*common*_ only, (4) composite score, (5) *PRS*_*common*_ + *PRS*_*RE*_+ *PRS*_*burden*_ by five subgroups: (1) all samples, (2) samples with top/bottom 10% PRS, (3) samples with top/bottom 5% PRS, (4) samples with top/bottom 1% PRS, and (5) samples with top/bottom 0.5% PRS. The black vertical lines represent the 95% confidence interval of the *R*^2^ estimates.

Our method exhibited marginally lower predictive performance for binary traits, as measured by enrichment of cases among high-risk groups and AUC, compared to the burden approach. We observed that for chosen binary traits, there were fewer genes associated with the trait, and their signals appeared weaker when contrasted with tested continuous traits. Nonetheless, our method offers potential benefits, as it can enhance predictability by combining *PRS*_*RE*_with *PRS*_*common*_ and *PRS*_*burden*_, which yielded better results. (**Supplementary Table 3, 4** and **Supplementary Note A**).

To further evaluate the utility of RareEffect, we compared its performance against other recent rare variant PRS methods incorporating functional annotations, including PrimateAI-3D, AlphaMissense, and DeepRVAT (**Supplementary Note B**). Across five phenotypes tested, RareEffect consistently demonstrated higher predictive accuracy, both as a standalone rare variant PRS and when combined with common variant PRS. For instance, in the case of LDL cholesterol, RareEffect achieved an *R*^2^ of 0.0340, more than twice that of the other methods. Moreover, when evaluating prediction performance in individuals at the extremes of the PRS distribution (top/bottom 0.5%), RareEffect yielded the highest improvement in composite score accuracy, with an *R*^2^ gain of 0.1066, which is four times larger than the next best approach PrimateAI-3D (*R*^2^ gain 0.0273) (**Supplementary Table 5** and **Supplementary Figure 4, 5**).

We further examined the relationship between phenotype outliers and the PRS in identifying individuals at high risk for common diseases^18^. We first defined the phenotype outliers as individuals with phenotype value exceeding a certain z-score cutoff and calculated the proportion of individuals with high PRS among phenotype outliers. Specifically, for LDL cholesterol levels, *PRS*_*RE*_successfully pinpointed individuals at phenotypic extremes, who exhibited a tenfold higher likelihood of possessing a *PRS*_*RE*_falling within 0.1^st^ percentile compared to the baseline population (**Supplementary Figure 6**). *PRS*_*RE*_and *PRS*_*common*_ utilize distinct set of variants and show minimal correlation (**Supplementary Table 6**). Therefore, integrating these models into a unified framework enables the identification of a significantly larger cohort at high risk than achievable through *PRS*_*common*_ alone.

### Simulation study

To assess the estimation accuracy of our method, we conducted extensive simulations under diverse scenarios (see **Methods**) for both binary and quantitative traits. To mimic real data, we utilized actual genotypes from the UKB dataset, specifically the array-genotyped data for common variants (MAF≥0.01) and the UKB WES data for rare variants (MAF≤0.01).

We compared the performance of variant-level effect size estimation from our method against other existing approaches such as linear regression, which is used for standard single-variant association test, or ridge regression using multiple evaluation metrics, including bias, mean absolute error (MAE), root mean squared error (RMSE), and coverage rate. For quantitative traits, our method consistently demonstrated a lower RMSE of 0.2101 on average, outperforming the comparative methods which showed RMSE of 0.2271 (ridge regression) to 0.2228 (linear regression) (**Supplementary Table 7, 8**) when estimating the effect size. For binary traits, our method also exhibited lower estimation error particularly in scenarios of low disease prevalence compared to ridge regression (**Supplementary Table 9**). Conversely, ridge regression showed marginally reduced RMSE compared to our method in instances of high disease prevalence; however, the difference in estimation accuracy remains modest.

To evaluate the prediction performance, we compared true *G*_*rare*_ *β* of simulated data and predicted 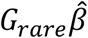, where *G*_*rare*_ include all rare variants across 50 causal genes. The results showed that RareEffect achieved lower MAE and RMSE on average compared to the comparative methods (**Supplementary Table 10**). We note that RareEffect, similar to Ridge regression, exhibited a higher calibration slope, reflecting the inherent shrinkage of the method. However, because polygenic risk score (PRS) models are typically subjected to model calibration prior to application, we emulated this by calibrating the slope using a subset of samples via regression model. As expected, after regression calibration, all methods achieved a calibration slope of approximately 1.

We also evaluated the performance of RareEffect on the estimation of gene-level heritability comparing with RARity^35^, which uses adjusted R^2^ (**Supplementary Note CE**). Our analysis shows that RareEffect tends to perform better when the true heritability is small and less biased than RARity, but RARity can perform better when true heritability is high (**Supplementary Table 11**). Additional analysis of different sample sizes showed that RareEffect stably estimate heritability with sample size > 100,000 (**Supplementary Table 12**).

### Computation and memory cost

Analyzing 166,960 samples from the UKB WES data to estimate the effect size of the *DOCK6* gene, which contains 4,114 rare variants, we observed that the computation time for RareEffect with a simulated phenotype was approximately 90% lower than that of ridge regression. Specifically, RareEffect completed the analysis in 4.2 seconds, compared to 44.6 seconds for ridge regression (**Supplementary Figure 7)**. The memory usage for RareEffect during the analysis was 1.14GB for *DOCK6* gene (**Supplementary Figure 8**). For binary traits, an additional step of performing Firth bias correction is required. We observed that the normal Firth bias correction process took 708 seconds to analyze a gene with 250 variants (after collapsing) across 342,409 individuals. However, by implementing our fast version of Firth correction, the computation time was dramatically reduced to 2.9 seconds (**Supplementary Figure 9**).

## Discussion

Our study introduces a novel method aimed at estimating the effect size of rare variants and can be extended to estimate gene-level effect size by employing a two-stage framework of generalized linear mixed models. By leveraging the variant-level effect size estimates obtained through our approach, we can examine the collective impact of rare variants within a gene and quantify their contribution to the overall heritability of complex traits.

In order to obtain accurate estimates for effect sizes while optimizing computational efficiency, we employed several techniques in our analysis. First, we utilized the FaST-LMM^23^ algorithm to expedite computation and reduce memory usage. Unlike GWAS settings where FaST-LMM relies on subsampling variants to reduce rank of genetic relatedness matrix, RareEffect operates in a regime where the number of variants per gene (*k*) is inherently much smaller than the sample size (*n*). This allows us to apply the FaST-LMM algorithm without approximation, reducing the time complexity while retaining exact computation. Second, we implemented the optimized version of Firth bias correction by utilizing the sparsity of genotype matrix and skipping the computation of hat matrix at every iteration. Third, we employed a collapsing strategy that reduces the sparsity of the data, similar to the approach employed in SAIGE-GENE+. These algorithmic approaches accelerate the estimation process and enhance computational scalability, particularly when dealing with large-scale datasets.

Prediction error variance (PEV) estimated in our method can be used to calculate confidence intervals. However, because RareEffect employs a shrinkage-based approach, and confidence intervals constructed using these estimates may not reliably cover the true effect sizes. In particular, we observed that the coverage rate for true causal variants was generally low, which reflects a known limitation of shrinkage estimators^36-38^. Nonetheless, for null variants, the confidence intervals consistently captured zero (**Supplementary Table 7**), indicating that the method accurately identifies non-causal variants. Furthermore, among the cases where the confidence intervals failed to cover the effect size of true causal variant, more than 99.999% of the intervals were positioned closer to zero than the true beta value, suggesting that these failures tend to occur in a conservative direction (**Supplementary Table 13**). Therefore, although the overall coverage rate is reduced, this does not appear to increase type I error but rather reflects a conservative bias in the effect size estimation. Among the methods compared, multiple linear regression exhibited lower coverage for null variants, suggesting potential inflation of type I error rates. Additionally, since variant-level effect size estimation involves multiple statistical tests, multiplicity adjustment may be necessary. Standard approaches such as Bonferroni adjustment^39^ or false discovery rate (FDR) control^40^ can be applied, and multiplicity-corrected confidence intervals for odds ratios^41^ can also be considered.

To assess genetic differences by sex, we compared estimated gene-level heritability between genetic sexes for three lipid traits (HDL, LDL and TG) (**Supplementary Note D). Supplementary Table 14** shows estimated heritability for top 10 genes. Overall, the estimated heritabilities are generally consistent with very high Pearson correlation.

In principle, the framework of RareEffect can be extended to other variant types such as copy number variants (CNVs) and intergenic variants, provided appropriate grouping strategies are defined. For intergenic variants or CNVs, the key challenge lies in defining biologically meaningful groups—such as proximity to regulatory elements or shared structural features— after which RareEffect’s variance component modeling can be applied.

Beyond its immediate applications in effect size estimation, our proposed method can enhance polygenic risk prediction models. Traditionally, these models have relied on common variants. Our analysis reveals that the correlation between *PRS*_*RE*_and phenotype values in the general population is not substantial (**Supplementary Figure 10**), so can provide complementary information.

AI-based methods^22,42^ have been developed to enhance the pathogenicity prediction of rare variants. Although these approaches can help to identify effect sizes of pathogenic variants and can be used for risk prediction, they may not be as effective for identifying beneficial or gain-of-function variants. As shown in our study, these methods have substantially low predictive performance than RareEffect. Additionally, the performance of these methods can be limited when applied to non-protein-altering variants. Our approach can accommodate the predictively pathogenic variants identified by AI-based models by forming them into a separate category, thereby enhancing performance.

RareEffect uses the MoM estimator to account for LD within a gene when estimating variance components. While this approach effectively adjusts for LD in most cases, it presents a challenge when the overall genetic effect of a gene is small. In such cases, the MoM estimator can produce negative variance component estimates, making the adjustment infeasible (**Supplementary Table 15**). When this occurs, the unadjusted marginal variance component estimates need to be used, which can lead to a slight overestimation of gene-level heritability. Although this issue is relatively infrequent and primarily affects genes with minimal contribution to heritability, it remains an important consideration when interpreting results.

We note that RareEffect is specifically designed to estimate gene-level heritability rather than exome-wide heritability. Estimating exome-wide heritability would require aggregating gene-level heritability estimates across all genes, but doing so involves additional assumptions. For instance, RARity conducted extensive pruning to adjust for long-range LD. Similar approach should be used to apply RareEffect to estimate exome-wide heritability.

In our study, gene-level effect directions were estimated using MAF-weighted effect sizes of predicted loss-of-function (LoF) variants. This approach performed well in both simulation studies and real data analyses, yielding results concordant with the true effect directions in simulations and with burden test results in real data (**Supplementary Note E**). The method assumes that predicted LoF variants within a gene have consistent effect directions. While this assumption would generally hold, exceptions may arise due to true biological heterogeneity or misclassification of functional impact. In such cases, discrepancies between the estimated and true gene-level effect directions may occur, potentially influencing downstream analyses.

Our study has several limitations. While RareEffect demonstrates comparable or superior performance in estimating the effect size for simulated binary phenotypes, our evaluation revealed only marginal enhancements in predictive performance, compared to the common variant PRS, across tested disease phenotypes, and these improvements were generally not statistically significant (**Supplementary Table 16**). This could be attributed to the trait’s reliance on a limited number of ultra-rare variants, which our method collapses into super-variants, thereby complicating the estimation of variant-level effect sizes for true causal variants. In addition, the binary phenotypes analyzed in this study generally had a much smaller number of significantly associated genes, which likely limited the potential benefit from rare variant PRS in these traits. Despite its efficacy in effect size estimation, RareEffect’s ability to improve prediction accuracy remains constrained, highlighting a potential area for future refinement and investigation for risk prediction of binary traits. Additionally, it is important to note that RareEffect is based on BLUP estimate, characterized by shrinkage properties, leading to biased estimates of effect sizes. To reduce bias, we also have implemented adaptive ridge for L0-regularization approach^43^ in the top of BLUP estimation. (**Supplementary Note F**). Adaptive Ridge provided improvement in bias (**Supplementary Table 7)**, but phenotype prediction was not improved. Further investigation is warranted and left for future work.

In summary, our results demonstrate that incorporating information from rare variants enables the accurate estimation of gene-level and variant-level effect sizes, as well as the identification of high-risk individuals who might remain undetected by conventional polygenic risk scores (PRS) methods relying on common variants.

## Methods

### Generalized linear mixed model

We denote the phenotype of the *i*th individual using *y*_*i*_ for both quantitative and binary traits in a study with sample size *n*. ***X*** represents the *n* × (*p* + 1) vector with *p* covariates including the intercept, and ***G***_*j*_ is the *n* × *k* matrix representing the minor allele counts for *k* rare variants in gene or region *j*. The generalized linear mixed model (GLMM) can be expressed as:

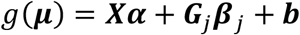

where ***µ*** is the mean phenotype, 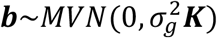 is the random effect, and ***K*** is an *n* × *n* genetic relatedness matrix (GRM). And *g* is the link function which is an identity function for quantitative traits with error term 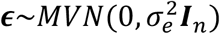 and a logit function for binary traits. The parameter **α** is a (*p* + 1) × 1 vector of fixed effect coefficients and ***β***_*j*_ is a *k* × 1 vector of the random genetic effect.

### Fitting the null generalized mixed model (step 1)

We used the average information restricted maximum likelihood (AI-REML) algorithm to fit the null GLMM (i.e., *H*_0_: ***β***_*j*_ = **0**) as in SAIGE step 1.

### Estimation of the gene-level (region-level) heritability (step 2)

We estimate the effect size of rare variants using the following linear mixed model:

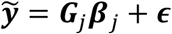

Where 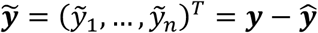 for quantitative traits, 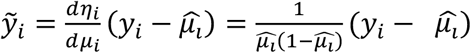, a working residual from iteratively reweighted least squares (IRWLS) for binary traits, and 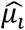 is the mean phenotype for individual *i*, which can be obtained from step 1. When obtaining ***G***_*j*_, as in SAIGE-GENE+, we collapsed ultra-rare variants with minor allele count (MAC) ≤ 10 by each gene and functional group to reduce the sparsity. We further implemented an option to collapse all loss-of-function (LoF) rare variants into a single column^44^, irrespective of their minor allele count, adopting the burden approach. This approach is predicated on the assumption that all rare LoF variants share a common effect size and direction. For binary traits, when we use working residuals, the variance of 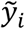 differs by individual, so we cannot apply the optimization technique of FaST-LMM. Therefore, we divide both sides by the square root of variance of 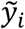 to make the variance be the same across individuals. We estimate the effect size using the modified model, for binary traits:

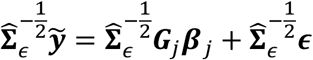

where 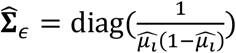.

In this model, the prior distribution of ***β***_*j*_ are assumed to follow *M VN*(0, τΣ), and the noise ϵ is assumed to follow *M VN*(0, ψ***I***_*n*_) for quantitative traits, while we assume 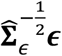 follows *M VN*(0, *ψ****I***_*n*_) for binary traits. When there is no prior knowledge of the correlation within ***β***, Σ is set to be an identity matrix. But in general, Σ does not have to be an identity or a diagonal matrix.

To estimate the variance component parameters (τ, *ψ*), we use factored spectrally transformed linear mixed models (FaST-LMM) algorithm. Let 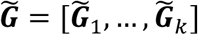 be an *n* × *k* genotype matrix of the region with 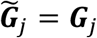 for quantitative traits and 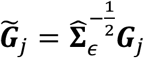 for binary traits. The variance of 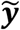 (quantitative traits) or 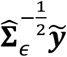 (binary traits) can be written as

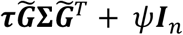

Traditional approaches to estimate the variance components require either calculating inverse matrix or conducting spectral decomposition of the *n* × *n* matrix 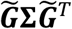, so the time complexity is of *O*(*n*^3^). In contrast, FaST-LMM algorithm uses the fact that 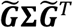 has rank at most *k*, so to reduce the computation complexity. Suppose 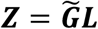 is an *n* × *k* matrix where ***L*** is a Cholesky factor of Σ. Then 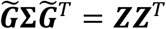. FaST-LMM carries out singular value decomposition on ***Z*** and calculate likelihood for (τ, *ψ*). With given *n* ≫ *k*, calculation of ***Z*** and its singular value decomposition requires only *O*(*nk*^2^) of time complexity. And we further improved the computation efficiency utilizing the sparsity of ***Z***^45^. In biobank-scale data, *n* is in the hundreds of thousands, and *k* is in the tens to hundreds on average which means *n* ≫ *k*.

Using the above optimization technique, we estimate the variance components by each group. Consider one group (LoF, missense or synonymous) in a single gene *j* in the model:

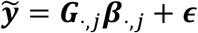

We first marginally estimate the maximum-likelihood estimator (MLE) of variance components τ_*LoF,j*_, τ_*mis,j*_, and τ_*syn,j*_. As the marginal estimates do not account for LD among variants in different groups, we adjust the estimates using method of moments (MoM) approaches^46^. The MoM estimator can be obtained by solving the following linear system:

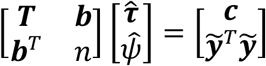

where ***T*** is a 3 × 3 matrix for joint estimation or a scalar (1 × 1 matrix) for marginal estimation with entries 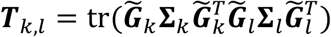 where *k, l* ∈ {1, 2, 3} for joint estimation. ***b*** is a 3-vector for joint estimation or a scalar for marginal estimation with entries 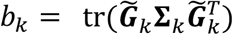, ***c*** is a 3-vector for joint estimation or a scalar for marginal estimation with entries 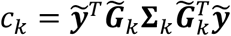, and 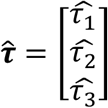 where (τ_1_, τ_2_, τ_3_) denotes (τ_*LoF,j*_, τ_*mis,j*_, τ_*syn,j*_) for a single gene *j*, respectively.

After estimating the marginal and joint MoM estimator of variance components, we adjust the MLE of the marginal variance components by:

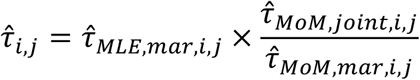

where *i* ∈ {*LoF, mis, syn*}. The MoM estimator of the variance component was not directly utilized in our study since the MoM estimator could yield negative variance components. In cases of the variance component estimated by the MoM is negative, we used marginal variance component without adjustment. Both MoM and likelihood-based approaches exhibited the similar trends of variance components (**Supplementary Figure 11**). Additionally, we assume that the variance explained by rare variants in a single gene is negligibly small^35^ compared to the total variance of 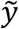, therefore, 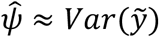.

We estimated the heritability from rare variants of gene *j* using these adjusted variance components. In a joint model,

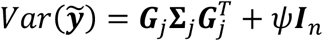

Where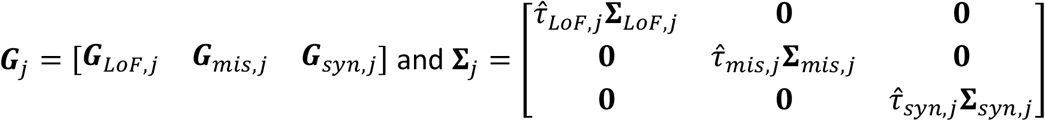.

Therefore,

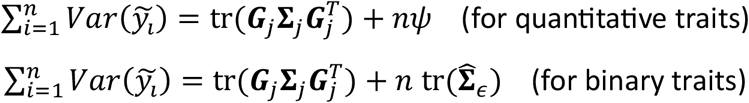

Subsequently, the heritability from rare variants of gene *j* can be denoted as:

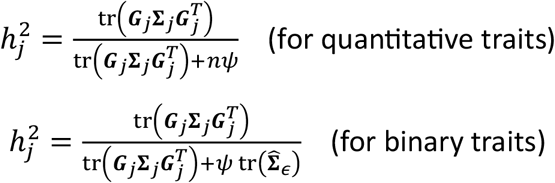

Additionally, to determine the direction of the gene-level effect, we obtained the sign of the linear combination of the effect sizes of loss-of-function variants in a gene, weighted by their MAFs:

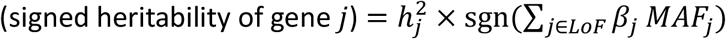

This measure gives the information of the magnitude of genetic effects from rare variants in a single gene and its direction of effects.

### Estimation of the variant-level effect size (step 3)

The effect sizes at variant-level resolution are estimated using the adjusted variance components in the previous step by:

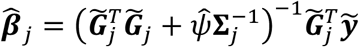

for each gene or region.

We further estimated the prediction error variance (PEV) by:

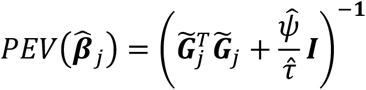

for each gene or region. Using this PEV, we can obtain confidence intervals for effect sizes.

For binary traits, the Firth bias correction^31^ is a more accurate method to estimate SNP effect sizes^47-49^, particularly in cases marked by a significant case-control imbalance. We incorporate this correction into our analytical framework. Additionally, we introduce an L2 penalty term to account for the prior distribution of ***β***. The Firth corrected effect estimates can be calculated numerically by optimizing the following objective function:

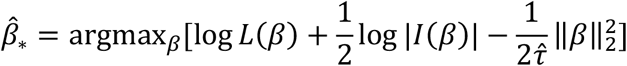

where *L* denotes the likelihood function and *I* is the Fisher information.

To improve computational efficiency, we developed the fast implementation of Firth bias correction. First, we compute the hat matrix, 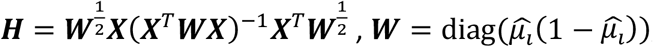, only once rather than recalculating it at each iteration, assuming that the iterative weights (***W***) change slowly as a function of the mean µ_*i*_ ^26,50^. We compared, using both simulation and real data, the difference in effect size estimates between computing the hat matrix once and computing it at each iteration. Our findings indicate minimal differences in the effect size estimates (**Supplementary Figure 12**). Given that Firth correction is applied to each variant, we iterate the hat matrix calculation for *k*_*F*_ times, where *k*_*F*_ represents the number of variants that need to be corrected. This strategy results in a reduction in the computational complexity with Firth correction from *O*(*Mnk*_*F*_) to *O*(*nk*_*F*_), where *M*is the average number of iterations needed for Firth corrected effect size convergence. Second, we extend Firth correction to accommodate sparse genotype matrices. Specifically, we restrict our computations to individuals with non-zero genotypes when determining the score and Fisher information. This further reduces the time complexity of the Firth correction, now at *O*(*n*_*nz*_*k*_*F*_), where *n*_*nz*_denotes the number of individuals with non-zero genotype. Considering that we are estimating the effect size of rare variants with MAF < 0.01, we observed that *n*_*nz*_< 0.01*n* which implies that leveraging sparsity makes the estimation of effect sizes more than 100 times faster compared to a non-sparse approach.

We note that Firth correction is used when the absolute value of estimated effect size 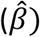 surpasses a predefined threshold. In this study, we used threshold of log 2 ≈ 0.693 for simulation studies and real data analysis.

RareEffect utilizes a shrinkage-based framework for estimating variant-level effect sizes, which inherently introduces bias. To mitigate this limitation, we incorporated an optional extension based on an adaptive ridge approach that provides a computationally efficient approximation to L0-regularization (**Supplementary Note F**).

### Rare-variant PRS calculation

We further calculated polygenic risk scores using the variant-level effect sizes of rare variants (*PRS*_*RE*_) in genes with gene-level p-value from SAIGE-GENE+ lower than 2.5 × 10^−6^. *PRS*_*RE*_of individual *i* can be calculated as:

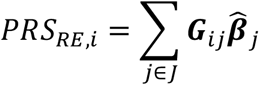

where *J* denotes a set of genes with gene-level p-value lower than 2.5 × 10^−6^. Additionally, we combined these *PRS*_*RE*_with *PRS*_*common*_ only. We applied PRS-CS^34^ and SBayesRC^33^ to obtain the variant-level weights for the calculation the *PRS*_*common*_. We compared the predictive performance of the *PRS*_*RE*_to *PRS*_*burden*_. *PRS*_*burden*_ were obtained in two different ways: (1) collapsing rare variants by functionality of the variants (LoF, missense, and synonymous), which is labeled as “Burden 1” in Supplementary Table, and (2) collapsing all rare variants into one super-variant regardless of functionality, labeled as “Burden 2” in Supplementary Table. In general, Burden 1 exhibited superior predictive performance compared to Burden 2 across most cases. Therefore, we used Burden 1 as the baseline for comparison in our analysis. After collapsing, we fitted the following linear model to estimate the per-allele effect sizes:

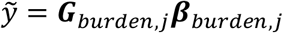

The burden PRS are also calculated as a linear combination of per-allele effect sizes and the collapsed genotypes of each individual.

We compared the predictive performance in terms of *R*^2^ for quantitative traits, and the area under receiver operating characteristic curve (AUROC) for binary traits of the following linear models:

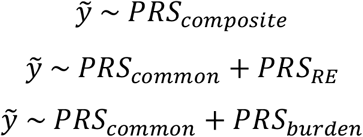

where *PRS*_*composite*_ is a linear combination of *PRS*_*common*_ and *PRS*_*RE*_with weights trained from the training set.

Additionally, we compared the performance of several recent methods for calculating rare variant PRS, including DeepRVAT, AlphaMissense, PrimateAI-3D, and RareEffect (See **Supplementary Note B**)

### UK Biobank data analysis

In this study, we used WES data of 393,247 White British participants in the UK Biobank. The UK Biobank is a UK-based prospective cohort of ∼500,000 individuals aged 40 to 69 at enrollment. We split the train and test data 8:2 randomly for the PRS evaluation. We applied quality control (QC) procedures prior to the analysis. We first removed redundant samples and individuals with sex mismatch or sex chromosome aneuploidy. Additionally, we further removed variants with a missingness rate across individuals > 0.1, HWE p-value < 10^−15^, and monomorphic variants. We generated group files, which define the list of variants in genes and its functional annotation, by using the loss-of-function transcript effect estimator (LOFTEE)^51^. We regarded a variant as loss-of-function (LoF) only in case of it is labeled as a high-confidence (HC) LoF variant, and variants with low-confidence (LC) were regarded as missense variants.

Using the data after QC, we applied our method to five quantitative traits (HDL cholesterol, LDL cholesterol, triglycerides, height, and body mass index) and five binary traits (breast cancer, prostate cancer, lymphoid leukemia, type 2 diabetes, and coronary atherosclerosis). We defined the disease by mapping ICD-10 codes to Phecodes using the PheWAS R package^52^.

### Simulation study

To generate outcome phenotypes, we used the following model for quantitative and binary traits:

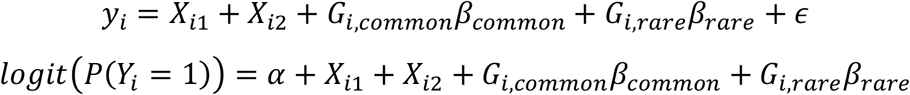

where *X*_*i*1_ and *X*_*i*2_ are covariates, and *G*_*i,common*_ and *G*_*i,rare*_ are genotype vectors of common variants and rare variants of *i*th individual, respectively. The intercept *α* for binary traits is determined by the disease prevalence. The covariates *X*_*i*1_ and *X*_*i*2_ were simulated from Bernoulli(0.5) and *N*(0, 1), respectively. For common variant effect, we randomly selected *L* =30,000 LD-pruned common variants with MAF > 1%, and assumed that the effect size of single common variant follows 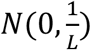. We selected 50 causal genes in UKB WES 200k data for generation of phenotypes.

We used eight different scenarios regarding rare variants: (1) proportion of causal variants, (2) effect size of causal variants, and (3) direction of effect within a single gene. For the proportion of causal variants, we assumed (1) 20% of LoF, 10% of missense, and 2% of synonymous variants, or (2) 30% of LoF, 10% of missense, and 2% of synonymous variants among rare variants that are not ultra-rare are causal. For ultra-rare variants, we assumed that the proportion of causal variants are three times higher than the non-ultra-rare variants, that is, (1) 60% of LoF, 30% of missense, and 6% of synonymous ultra-rare variants, or (2) 90% of LoF, 30% of missense, and 6% of synonymous ultra-rare variants are causal. Regarding the effect size of causal variants, we assumed that the absolute effect sizes of causal variants are (1) |0.5 log_10_ *MAF*| for LoF variants, and |0.25log_10_ *MAF*| for missense and synonymous variants, or (2) |0.3 log_10_ *MAF*| for LoF variants, and |0.15log_10_ *MAF*| for missense and synonymous variants. We further assumed that the effect directions are (1) the same among all causal variants in a single gene, or (2) same for 100% of LoF, 80% of missense, and 50% of synonymous variants, while remaining variants have the opposite direction of effect. For eight combinations of scenarios, we repeated the simulation for 1,000 times. To evaluate the prediction performance, we calculated *G*_*rare*_ *β*_*rare*_ and predicted 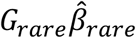. Additional model calibration was conducted using a linear regression model of *G*_*rare*_ *β*_*rare*_ and 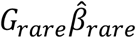 using 10% of simulated samples.

### Computation cost evaluation

We evaluated the computation time and memory usage using simulated data as described above, comprising 166,960 individuals of White British ancestry from the UKB WES 200k dataset. Additionally, we examined computation time and memory usage across subsets with sample sizes of 10k, 30k, 50k, and 100k. For each generative scenario, we reported the mean of 5 attempts for computation times and memory usage, comparing them with multiple linear regression, simple linear regression (as in GWAS), and ridge regression. The evaluation for linear regression and ridge regression was done using lm and glm functions in R, respectively.

## URLs

SAIGE (including SAIGE-GENE+): https://github.com/saigegit/SAIGE

DeepRVAT: https://github.com/PMBio/deeprvat

AlphaMissense scores: https://zenodo.org/records/8208688

PrimateAI-3D scores: https://primateai3d.basespace.illumina.com/

SBayesRC (GCTB): https://cnsgenomics.com/software/gctb

PRS-CS: https://github.com/getian107/PRScs

RARity: https://github.com/GMELab/RARity

## Data Availability

The analysis results for 100 phenotypes of UKB WES data analysis results are available at: https://storage.googleapis.com/leelabsg/RareEffect/RareEffect_effect_size.zip (variant-level effect size) and https://storage.googleapis.com/leelabsg/RareEffect/RareEffect_h2.zip (gene-level signed heritability).

## Code availability

RareEffect is implemented as a part of SAIGE software, which is an open-source R package, available at https://github.com/saigegit/SAIGE. RareEffect is available in SAIGE version 1.3.7 or higher.

### Author Contribution

K.N. and S.L. designed experiments. K.N. performed experiments and analyzed the UKB WES data. K.N. and S.L. implemented the software with input from W.Z.. M.K. and B.M. provided helpful advice. K.N. and S.L. wrote the manuscript with input from all co-authors.

## Acknowledgements

This research was supported by the Brain Pool Plus (BP+) Program through the National Research Foundation of Korea (NRF) funded by the Ministry of Science and ICT (2020H1D3A2A03100666) and the grants funded by the Ministry of Food and Drug Safety, Republic of Korea (Grant Number: 23212MFDS202). Minjung Kho was supported by the NRF grant funded by the Korean government (MSIT) (NRF-2022R1C1C2006474). This research was conducted using the UK Biobank Resource under application number 45227.

## Supplementary Figures

1. Estimated signed heritability for 10 tested traits
2. Predictive performance for 10 tested traits
3. Relationship between common variant PRS (*PRS*_*common*_) and RareEffect PRS (*PRS*_*RE*_)
4. Comparison of the performance of rare variant PRS methods for lipid phenotypes
5. Comparison of the performance of rare variant PRS methods when combined with common variant PRS (SBayesRC) for lipid phenotypes
6. Enrichment of high-risk individuals based on RareEffect PRS (*PRS*_*RE*_), common variant PRS (*PRS*_*common*_) and the composite score among phenotype outliers
7. Computation time of RareEffect, linear regression, and ridge regression
8. Memory usage of RareEffect by number of variants
9. Computation time of the fast implementation Firth bias correction and the standard Firth correction
10. Relationship between RareEffect PRS (*PRS*_*RE*_) and phenotype values
11. Comparison of estimated variance components between RareEffect and the MoM estimator
12. Comparison of estimated effect sizes between computing the hat matrix at every iteration and computing it only once in Firth bias correction
13. Distribution of RareEffect PRS

## Supplementary Tables

1. List of 100 analyzed traits in the UK Biobank
2. Predictive performance measured by R^2^ and AUC for the analyzed phenotypes in the UK Biobank
3. Predictive performance measured by mean/prevalence difference and enrichment for the analyzed phenotypes in the UK Biobank
4. Number of phenotypes showing improvement compared to common PRS performance
5. Comparison of predictive performance between RareEffect and DeepRVAT, AlphaMissense, and PrimateAI-3D
6. Pearson correlation between PRS from common variants and RareEffect PRS
7. Estimation performance for simulated continuous data across scenarios
8. p-values from paired t-tests for the difference in MSE
9. Estimation performance for simulated binary data across scenarios
10. Predictive performance for simulated data across scenarios
11. Estimation performance for gene-level heritability
12. Estimated gene-level heritability by sample size
13. Proportion of conservative estimates in confidence interval coverage failures
14. Estimated signed heritability by genetic sex for the top 10 genes
15. Proportion of negative MoM estimates of variance components
16. p-values from DeLong’s test for differences in AUROC
17. Predictive performance for continuous phenotypes in the UK Biobank after excluding the top 3 genes
18. Comparison of predictive performance stratified by genetic sex
19. Direction of estimated gene-level heritability
20. Comparison of predictive performance with adaptive ridge adjustment

