## Supplementary Notes for "Rare variant effect estimation and polygenic risk prediction"

#### A. Detailed results of the Comparison of PRS common and composite score in UK Biobank

We analyzed 61 continuous and 39 binary traits, constructing both a PRS based on common variants ( $PRS_{common}$ ) and a composite score integrating $PRS_{common}$ and $PRS_{RE}$ ($PRS_{composite}$). To assess performance improvements, we focused on individuals at the predictive extremes, restricting our analyses to the top and bottom 10%, 5%, 1%, and 0.5% of predicted risk scores.

For continuous traits, in addition to $R^2$, we evaluated the difference in standardized means between the high- and low-predicted groups. This additional metric was included because $R^2$ estimates can become unstable in phenotypic extremes due to increased variability in phenotypes. As shown in **Supplementary Table 4,** among the 61 continuous traits, 21, 12, and 6 phenotypes exhibited >5%, >10%, and >20% improvements in $R^2$, respectively, when using $PRS_{composite}$ compared to $PRS_{common}$. Similar trends were observed when using the standardized mean difference. The largest improvement was observed for Aspartate Aminotransferase (AST), with a 31% increase in $R^2$ and a 22% increase in the mean difference in top/bottom 0.5% individuals, respectively.

For binary traits, we assessed case enrichment (e.g., prevalence) within the high-risk groups and calculated the AUC among high- versus low-risk groups. Among the 39 binary traits, 7, 2, and 2 traits showed >5%, >10%, and >20% improvements in case enrichment when evaluated the enrichment among top 0.5% high risk individuals, respectively. When incorporating $PRS_{burden}$ into the $PRS_{composite}$, the corresponding numbers increased to 9, 4, and 2 traits, respectively. However, using AUC as the evaluation metric, we observed minimal improvements. Among the phenotypes, Myeloproliferative disease showed the largest improvement with 60% improvement in $PRS_{composite}$ in the enrichment among top 0.5%.

We additionally evaluated how much the prediction performance of $PRS_{composite}$ would decline when excluding the top three most significant genes from the score calculation for continuous traits (**Supplementary Table 17**). This analysis was not performed for binary phenotypes, as there were very few genome-wide significant genes identified, making such an assessment less meaningful. The degree of performance change varied substantially across different phenotypes. Focusing on the composite score of PRS-CS and RareEffect for individuals in the top and bottom 0.5% of the PRS distribution, we found that for HDL cholesterol, the $R^2$ increased from 0.5417 (using common PRS alone) to 0.6002 even after excluding the top three genes, compared to 0.6237 when including these genes—indicating only a modest drop in performance. In contrast, for LDL cholesterol, the $R^2$ increased from 0.5823 (common PRS alone) to just 0.5842 when the top three genes were excluded, whereas including these genes resulted in a substantially higher $R^2$ of 0.6809. These results suggest that the contribution of top genes to prediction accuracy differs notably depending on the phenotype.

### B. PRS method comparisons

We compared the performance of several recent methods for calculating rare variant PRS, including DeepRVAT[1], AlphaMissense[2], PrimateAI-3D[3,4], and RareEffect for the prediction of three cholesterol phenotypes. For DeepRVAT, we used the seed genes identified through its own gene discovery pipeline, while for RareEffect, AlphaMissense, and PrimateAI-3D, we defined significant genes based on SAIGE-GENE+[5] Cauchy-combined SKAT-O p-values, using a significance threshold of $2.5 \times 10^{-6}$ (corresponding to a Bonferroni correction for 20,000 genes).

To compute gene-level burden scores, DeepRVAT used its pretrained model to derive gene impairment scores, averaging the outputs from its ensemble of 30 models. We note that the phenotypes we tested were initially used to pretrain the DeepRVAT model. The all 34 annotations used in pretraining were used to calculate gene impairment scores. For AlphaMissense and PrimateAI-3D, the burden for each gene was computed as the weighted sum of rare variants within the gene, using each method's annotation scores as weights. Variants without available AlphaMissense or PrimateAI-3D scores were excluded from the burden calculation.

Consistent with the burden-based approach described in the Methods section, we calculated rare variant PRS by performing a linear combination of gene-level burden scores (or gene impairment scores in the case of DeepRVAT) with their estimated burden effect sizes. The resulting rare variant PRS was then combined with the common variant PRS using weights learned in the training set, and performance was evaluated on the test set using the final composite score. Due to the complexity of the pipeline and software compatibility issues, we were unable to run DeepRVAT on the DNAnexus platform. As a result, our evaluation was limited to the previous release of UK Biobank data (UKB WES 200K), which could be downloaded to a local server. We note that the gene impairment scores of DeepRVAT were derived from a pretrained model provided. Only the effect size of each gene impairment scores were estimated using the UKB WES 200K training set ($n = 133{,}125$). For all the other methods, UKB WES 470K training set ($n = 314{,}198$) were used to estimate each gene burden score effect sizes. Performance evaluation for all methods was carried out on the UKB WES 200K test set ($n = 33{,}125$).

### C. Gene-level heritability estimation evaluation

To evaluate the gene-level heritability estimation, we compared our method with RARity[6], a method to estimate heritability using adjusted $R^2$ (**Supplementary Table 11**). Our analysis revealed that RARity tends to underestimate the heritability of true causal genes, whereas RareEffect produces estimates that are closer to the true heritability values. This discrepancy appears to arise from RARity's exclusion of extremely rare variants, such as singleton and doubleton variants, when calculating heritability. In contrast, RareEffect incorporates these ultra-rare variants, leading to more accurate heritability estimates for causal genes. In terms of variance, RARity showed lower variance and overall lower RMSE in heritability estimates compared to RareEffect, indicating that adjusted $R^2$-based approaches perform well for heritability estimation. However, it is important to note that RARity is specifically designed for heritability estimation and does not provide variant effect size estimates, which is the primary objective of RareEffect.

Additionally, we assessed the impact of sample size on gene-level heritability estimates obtained using RareEffect. Since RareEffect collapses ultra-rare variants based on a minor allele count (MAC) threshold, smaller sample sizes result in a larger proportion of rare variants being classified as ultra-rare and subsequently collapsed. This process can lead to an underestimation of gene-level heritability, as fewer variants contribute individually to the variance estimation. To quantify the bias introduced by varying sample sizes, we conducted simulations and found that a sample size of at least 100,000 individuals is recommended to obtain reliable heritability estimates when using a MAC collapsing threshold of 10 (**Supplementary Table 12**).

### D. Sex difference analysis

To assess the influence of genetic sex on effect heterogeneity, we conducted an analysis using three real phenotypes from the UK Biobank: HDL, LDL, and triglycerides. We first stratified the training set into male and female groups and separately estimated gene-level heritability and effect sizes for each group. To evaluate the difference in gene-level heritability estimates, we calculated the Pearson correlation of the estimated values for the top 10 genes between the two groups. The correlations were 0.973 for HDL-C, 0.965 for LDL-C, and 0.846 for triglycerides (**Supplementary Table 14**). Additionally, we constructed polygenic risk scores (PRS) using variant-level effect sizes and assessed whether there were differences in prediction performance between males and females. Our analysis did not reveal any notable differences in PRS performance between males and females. We observed a slightly higher PRS accuracy, in terms of $R^2$, in females across all three phenotypes (**Supplementary Table 18**). This minor difference is likely due to the slightly larger sample size of females in the dataset, which may contribute to more stable estimates.

### E. Gene-level effect direction analysis

To validate the approach of inferring gene-level effect directions using the MAF-weighted sum of LoF variant effect sizes, we conducted both simulation studies and empirical comparisons using UK Biobank data. Since the true gene-level effect directions are unknown in real data, we compared the gene-level effect directions estimated by RareEffect with the directions of per-allele effect sizes obtained from gene burden tests. For the burden tests, we applied two different approaches: (1) collapsing all rare variants within the gene, and (2) collapsing only the loss-of-function (LoF) variants within the gene. In contrast, for the simulation data, where the true gene-level effect directions were predefined, we directly compared these true directions with the gene-level effect directions estimated by RareEffect. In simulations, the accuracy of inferred directions increased with true effect size, ranging from 87% to 92% concordance. In real data, the direction estimated by RareEffect matched the burden test direction in approximately 80% of significant genes when all rare variants were collapsed, and this agreement increased to 98% when limited to LoF variants (**Supplementary Table 19**).

### F. Implementation of adaptive ridge for L0-regularization

Since RareEffect estimates variant-level effect sizes using a shrinkage method, it inherently introduces bias. As a result, when evaluated using metrics such as bias, RareEffect appears to perform worse compared to unbiased estimators like the ordinary least squares (OLS) estimator. To address this limitation, we implemented an optional extension that applies an adaptive ridge method[7], which provides a computationally efficient approximation to L0-regularization. This approach leverages the variance component estimates obtained in Step 2 of the RareEffect framework.

The objective of L0-regularization is to maximize the penalized log-likelihood function:

$$\widehat{\boldsymbol{\beta}} = \operatorname{argmax}_{\beta}[\log L(\boldsymbol{\beta}) - \boldsymbol{\lambda} \cdot \|\boldsymbol{\beta}\|_{L_0}]$$

where $\boldsymbol{\lambda}$ is a vector with elements $\lambda_i = \frac{\psi}{\tau_i}$, $i \in \{LoF, mis, syn\}$ and $\psi$ and $\tau$ are the variance components estimates in Step 2. In order to avoid any numerical instabilities, we used the following formula to update the weights:

$$w_j = \begin{cases} \delta^{-2} \exp\left[-\frac{2}{\gamma}\log\left(1 + \left|\frac{\beta_j}{\delta}\right|^{\gamma}\right)\right] & \text{if } \left|\widetilde{\beta_j}\right| \leq \delta \\ \left|\widetilde{\beta_j}\right|^{-2} \exp\left[-\frac{2}{\gamma}\log\left(1 + \left|\frac{\beta_j}{\delta}\right|^{\gamma}\right)\right] & \text{if } \left|\widetilde{\beta_j}\right| > \delta \end{cases}$$

We used $\delta = 10^{-5}$ and $\gamma = 2$ in the study.

Using adaptive ridge for variant-level effect size estimation led to improvements in bias, MAE, and RMSE of $\widehat{\boldsymbol{\beta}}$ compared to the original estimates (**Supplementary Table 7**). However, when these adaptive ridge estimates were used for phenotype prediction, they did not offer a substantial advantage over the original RareEffect estimates (**Supplementary Table 20**).
