## Supplementary Figures for "Rare variant effect estimation and polygenic risk prediction"

#### Supplementary Figure 1. Estimated signed heritability for 10 tested traits.

Signed gene-level heritability from RareEffect for 10 traits. Gene-level p-values were obtained from SAIGE-GENE+, and we included genes with p-values < $2.5 \times 10^{-6}$. The $x$-axis represents the rank order of genes based on their gene-level p-values. Lower ranks correspond to genes with more significant p-values. The $y$-axis shows the signed gene-level heritability for each gene. Signed heritability indicates the direction (positive or negative) and magnitude of the genetic contribution of the gene to the trait.

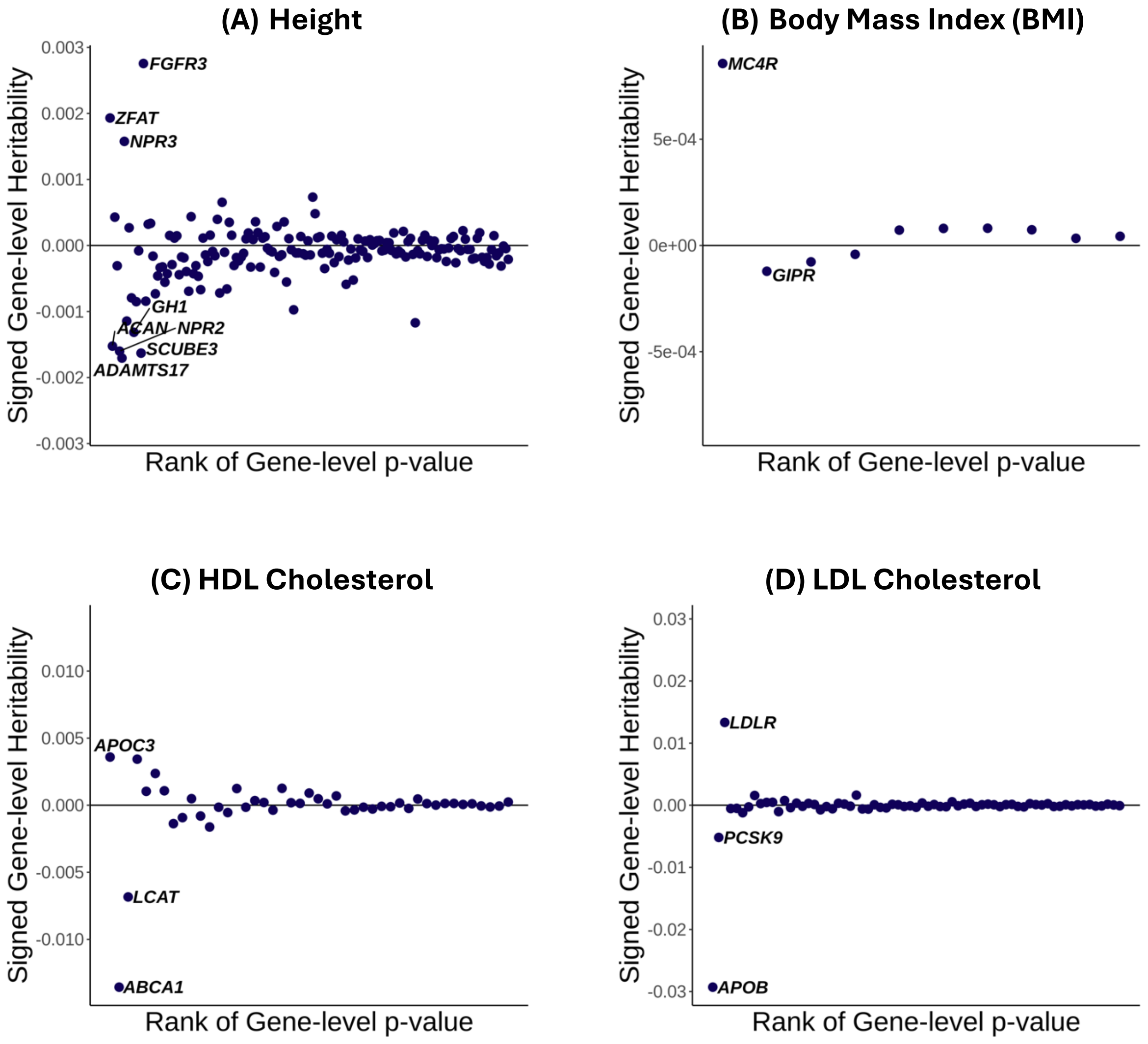

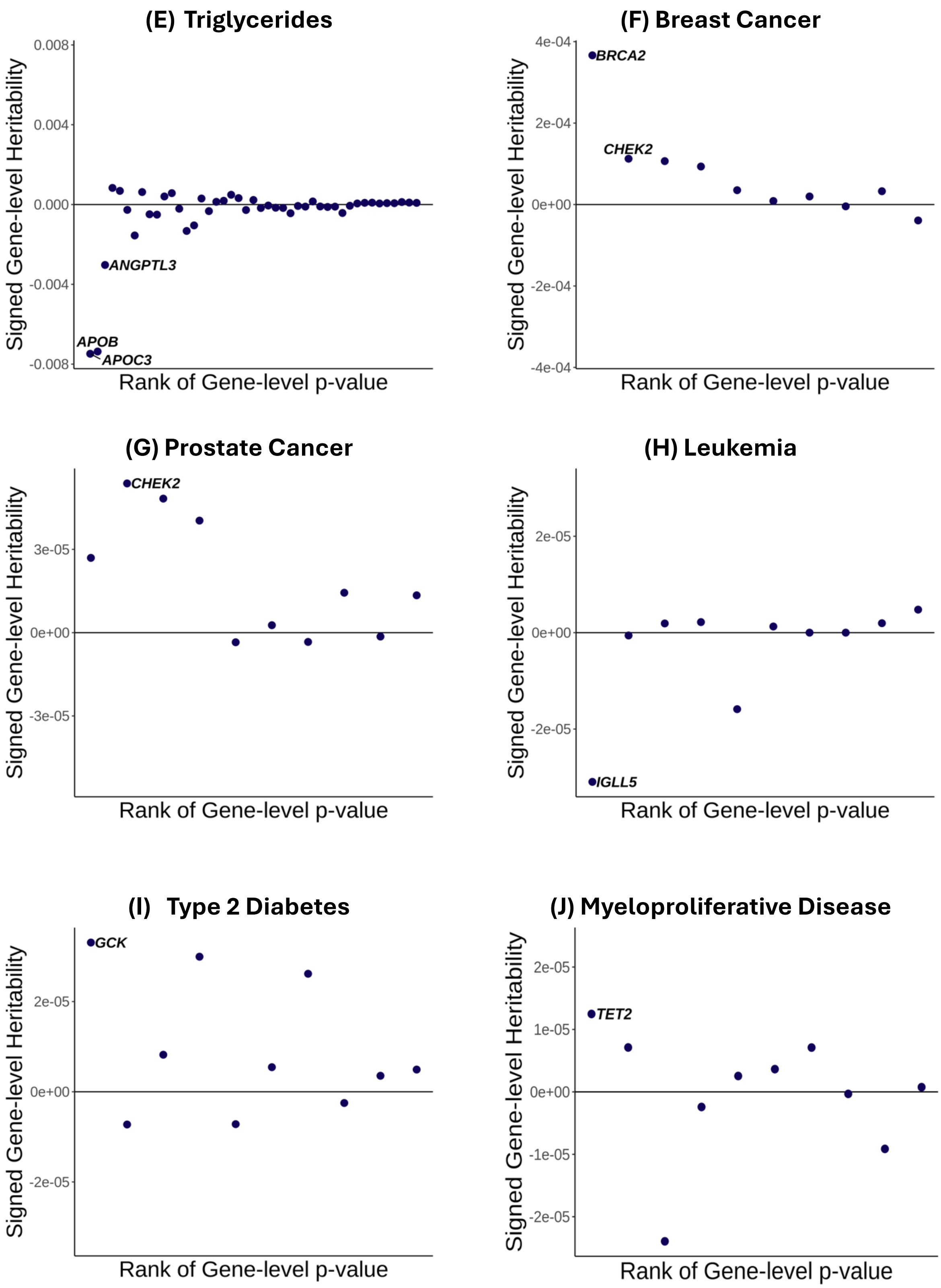
(E) Triglycerides
Signed Gene-level Heritability
0.008
0.004
0.000
-0.004
-0.008
ANGPTL3
APOB
APOC3
Rank of Gene-level p-value
(F) Breast Cancer
Signed Gene-level Heritability
4e-04
2e-04
0e+00
-2e-04
-4e-04
BRCA2
CHEK2
Rank of Gene-level p-value
(G) Prostate Cancer
Signed Gene-level Heritability
3e-05
0e+00
-3e-05
CHEK2
Rank of Gene-level p-value
(H) Leukemia
Signed Gene-level Heritability
2e-05
0e+00
-2e-05
IGLL5
Rank of Gene-level p-value
(I) Type 2 Diabetes
Signed Gene-level Heritability
2e-05
0e+00
-2e-05
GCK
Rank of Gene-level p-value
(J) Myeloproliferative Disease
Signed Gene-level Heritability
2e-05
1e-05
0e+00
-1e-05
-2e-05
TET2
Rank of Gene-level p-value

**Supplementary Figure 2. Predictive performance for 10 selected traits.**

We evaluated model performance using $R^2$ (left panel) and mean difference (right panel) for continuous traits, and AUC (left panel) and case enrichment (right panel) for binary traits. Five models were assessed: (1) $PRS_{burden}$ only, (2) $PRS_{RE}$ only, (3) $PRS_{common}$ only, (4) composite score, (5) $PRS_{common} + PRS_{RE} + PRS_{burden}$, and performance was evaluated across five subgroups: (1) all samples, (2) samples with top/bottom 10% PRS, (3) samples with top/bottom 5% PRS, (4) samples with top/bottom 1% PRS, and (5) samples with top/bottom 0.5% PRS. The black vertical lines represent the 95% confidence interval. For all 100 trait results, please see Supplementary Table 2 and 3.

**(A) HDL Cholesterol**

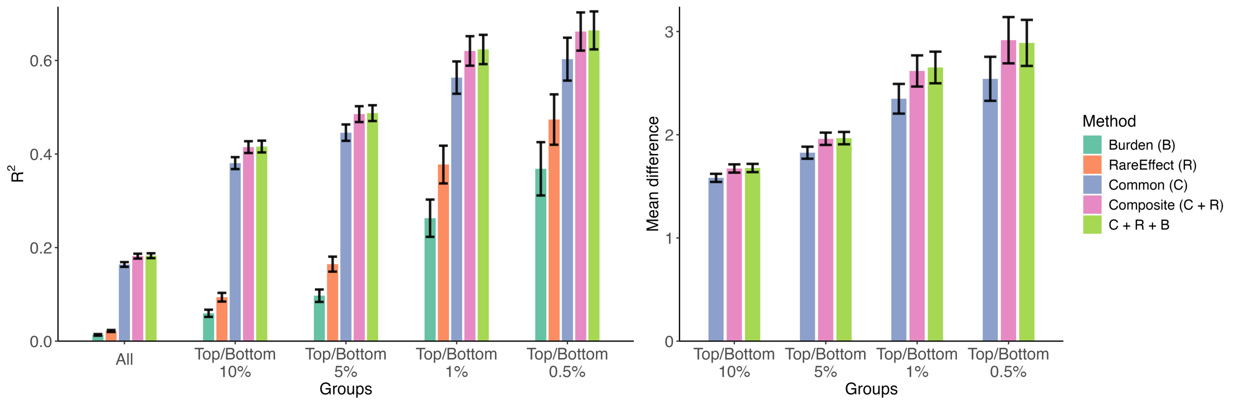


**(B) LDL Cholesterol**

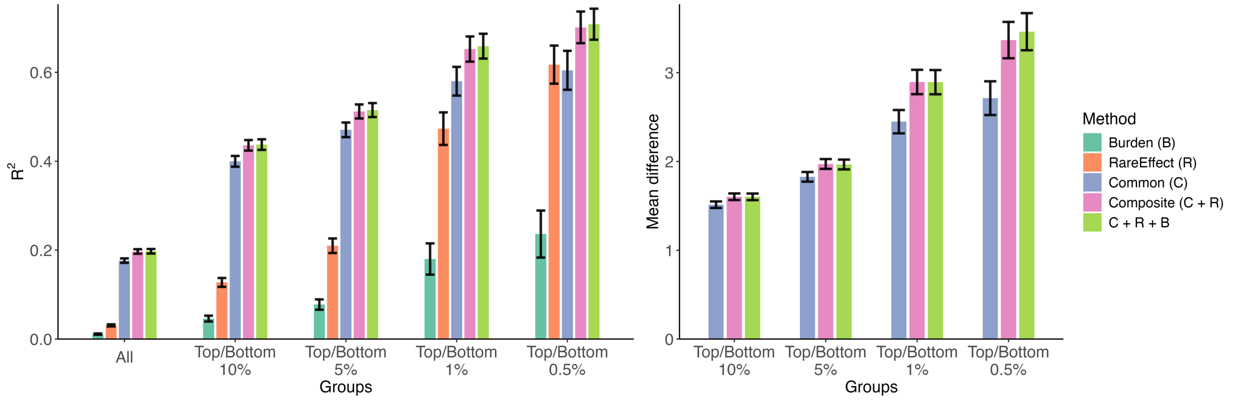

### (C) Triglycerides

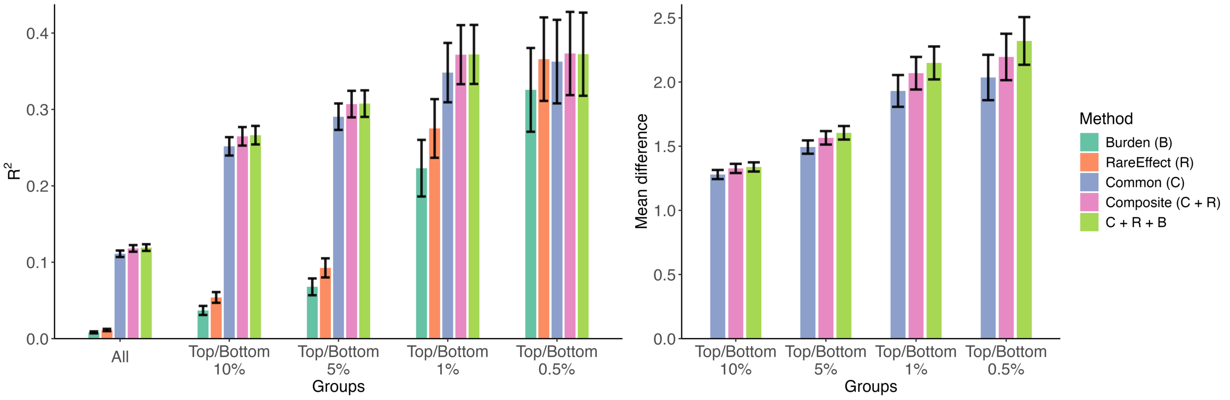


### (D) Standing Height

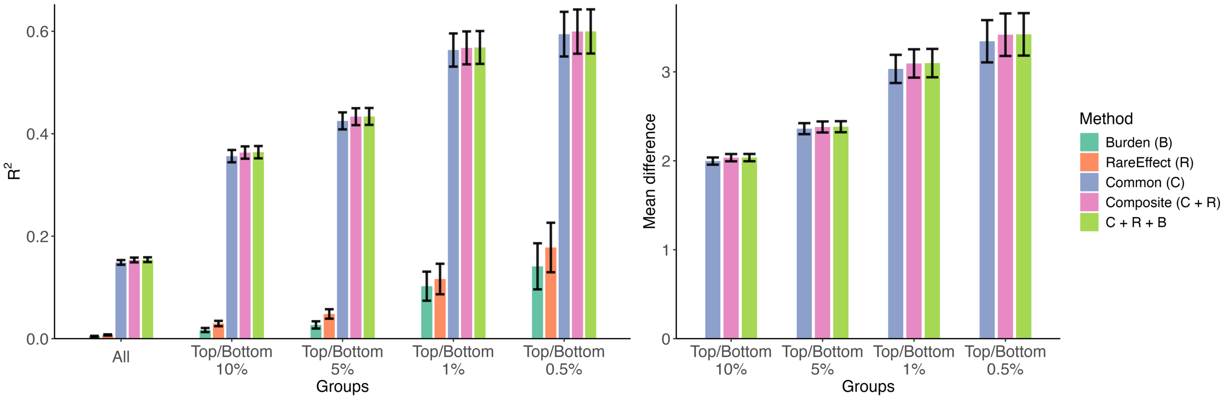


### (E) Body Mass Index (BMI)

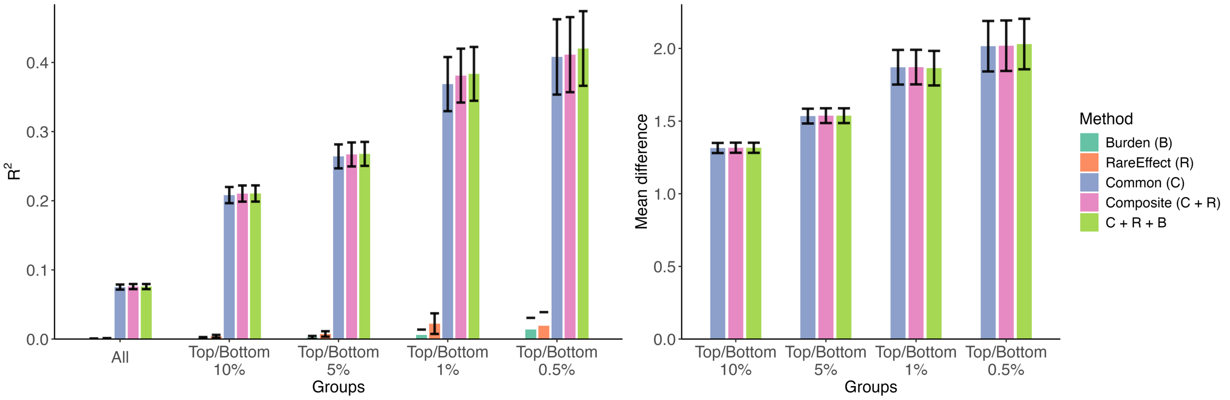

### (F) Breast Cancer

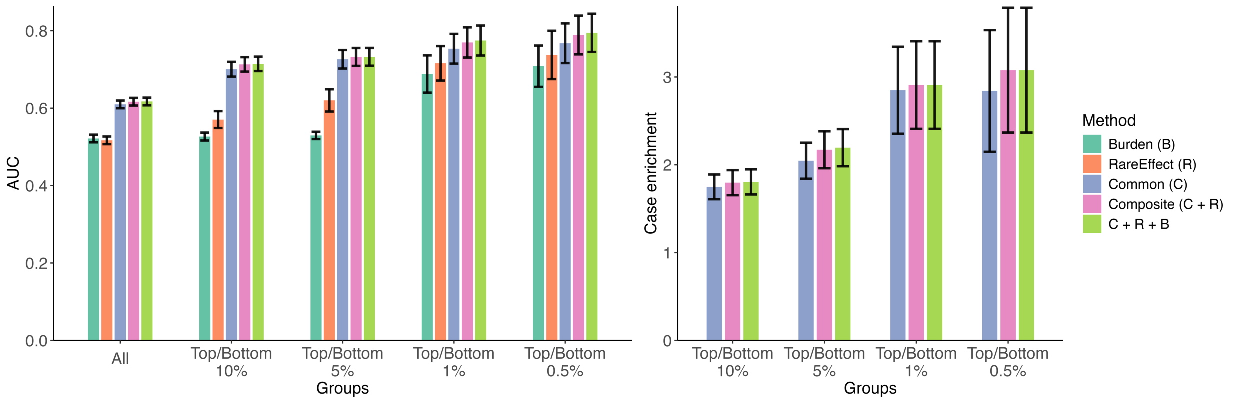


### (G) Prostate Cancer

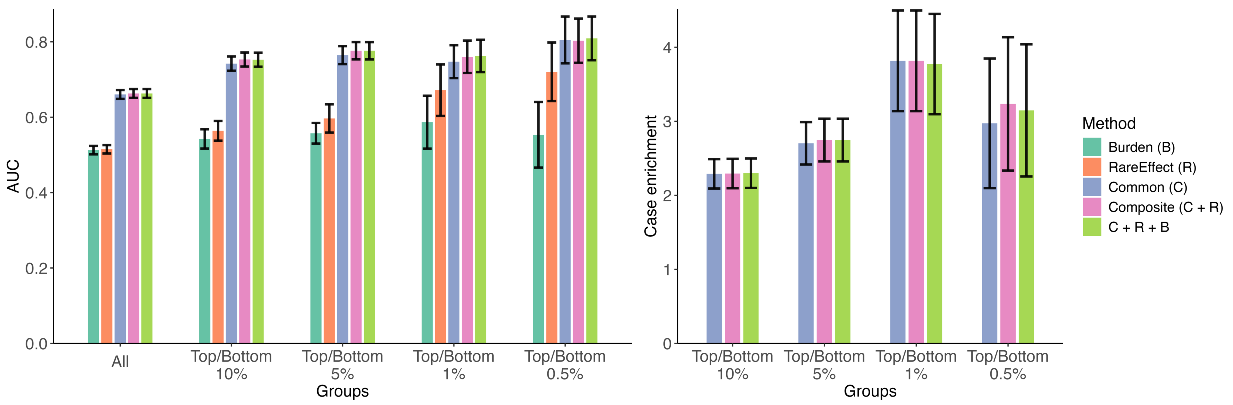


### (H) Leukemia

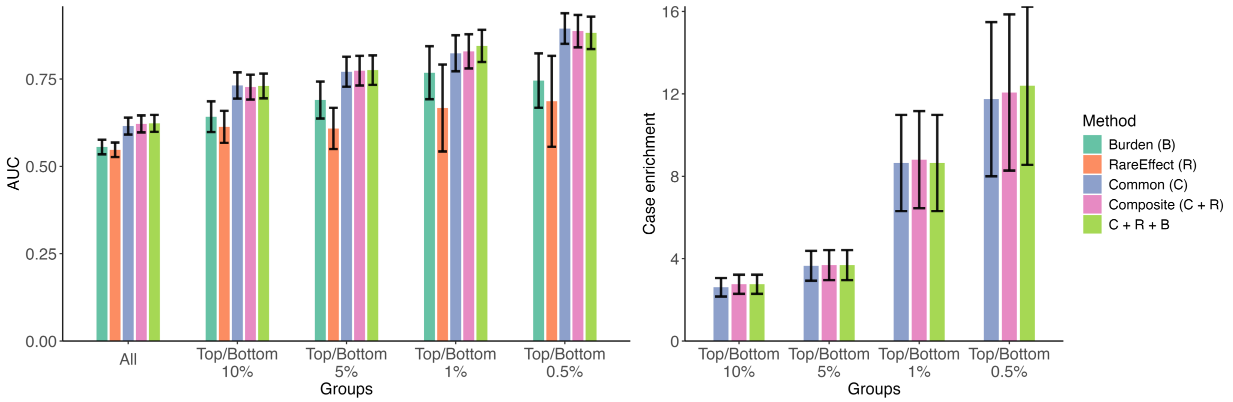

### (I) Type 2 Diabetes

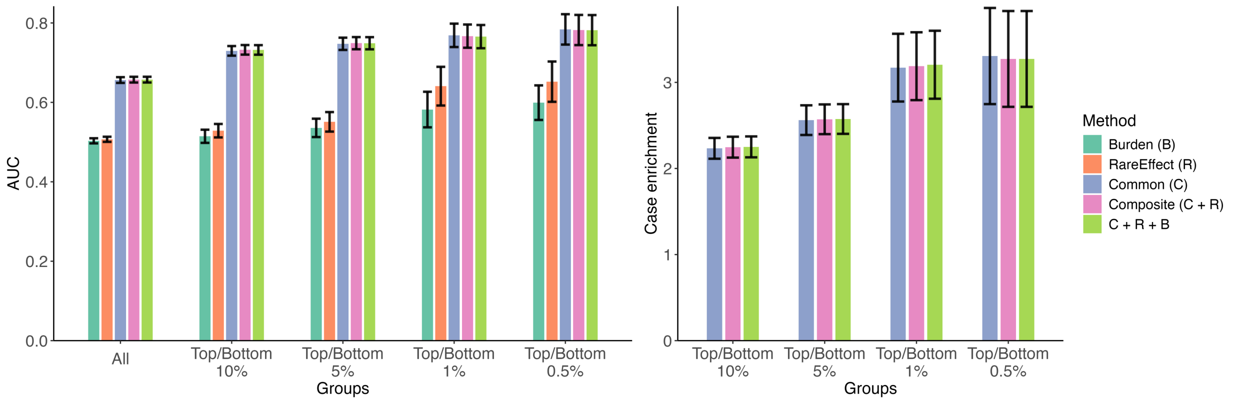


### (J) Myeloproliferative Disease

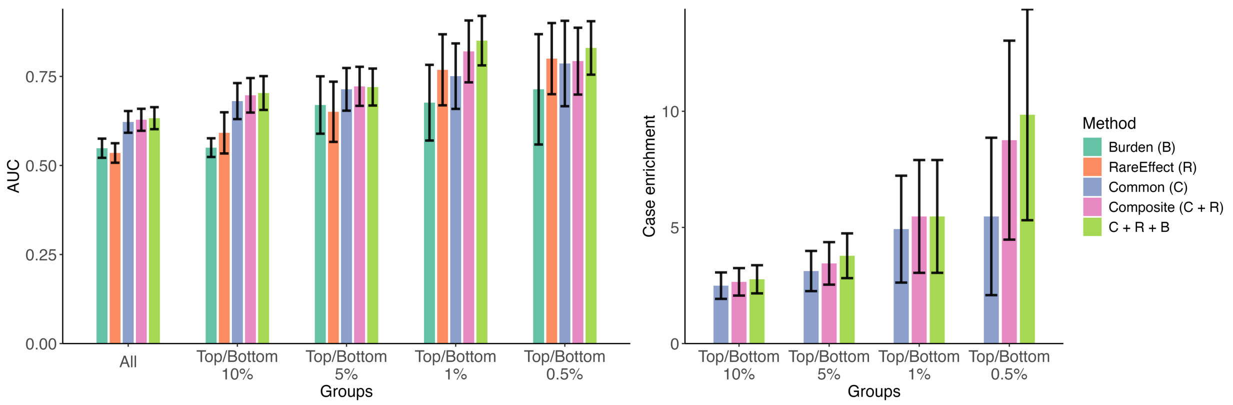

### Supplementary Figure 3. Relationship between common variant PRS ($PRS_{common}$) and RareEffect PRS ($PRS_{RE}$)

The $x$-axis represents $PRS_{RE}$, and the $y$-axis represents $PRS_{common}$ of each individual.

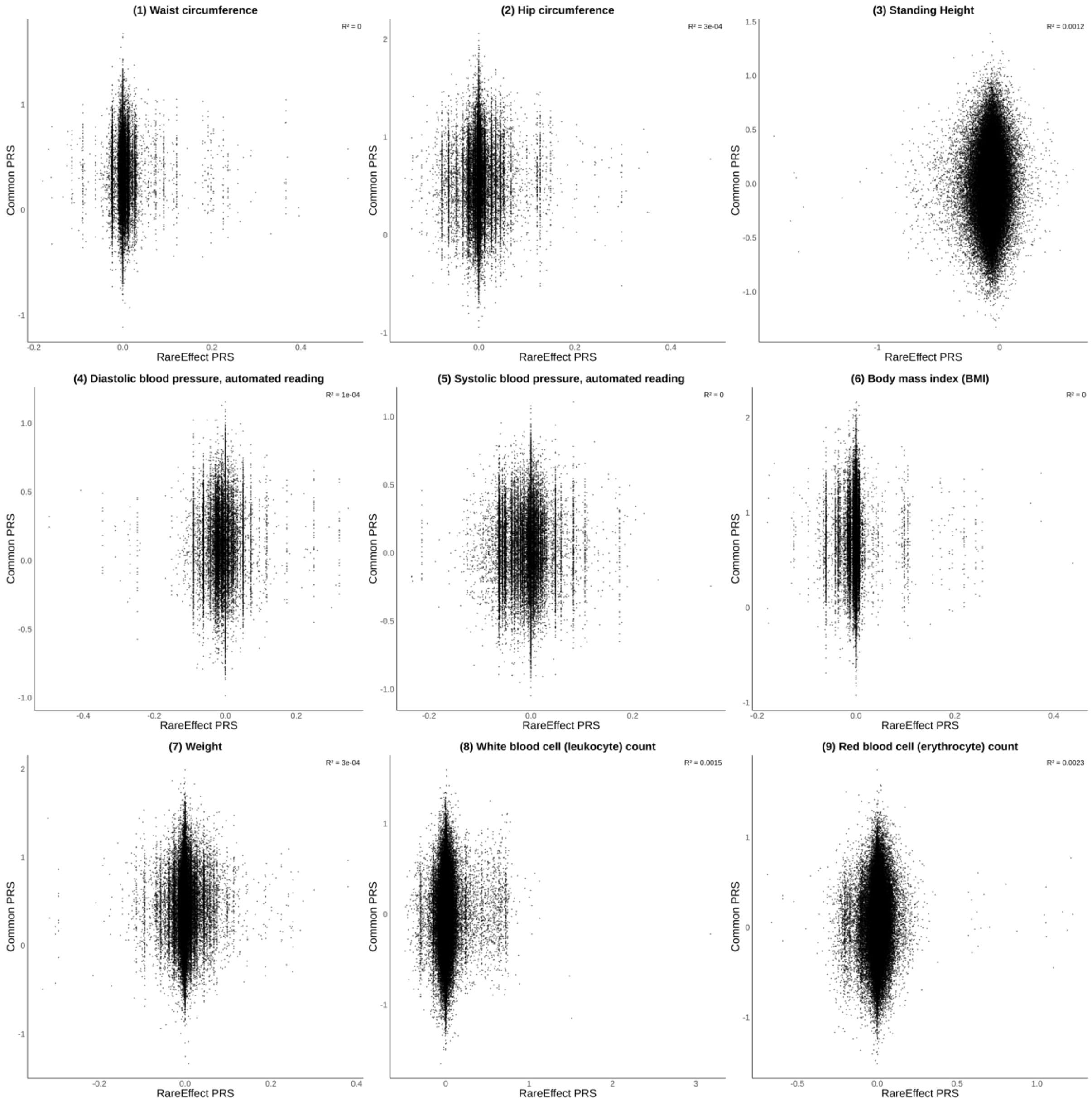

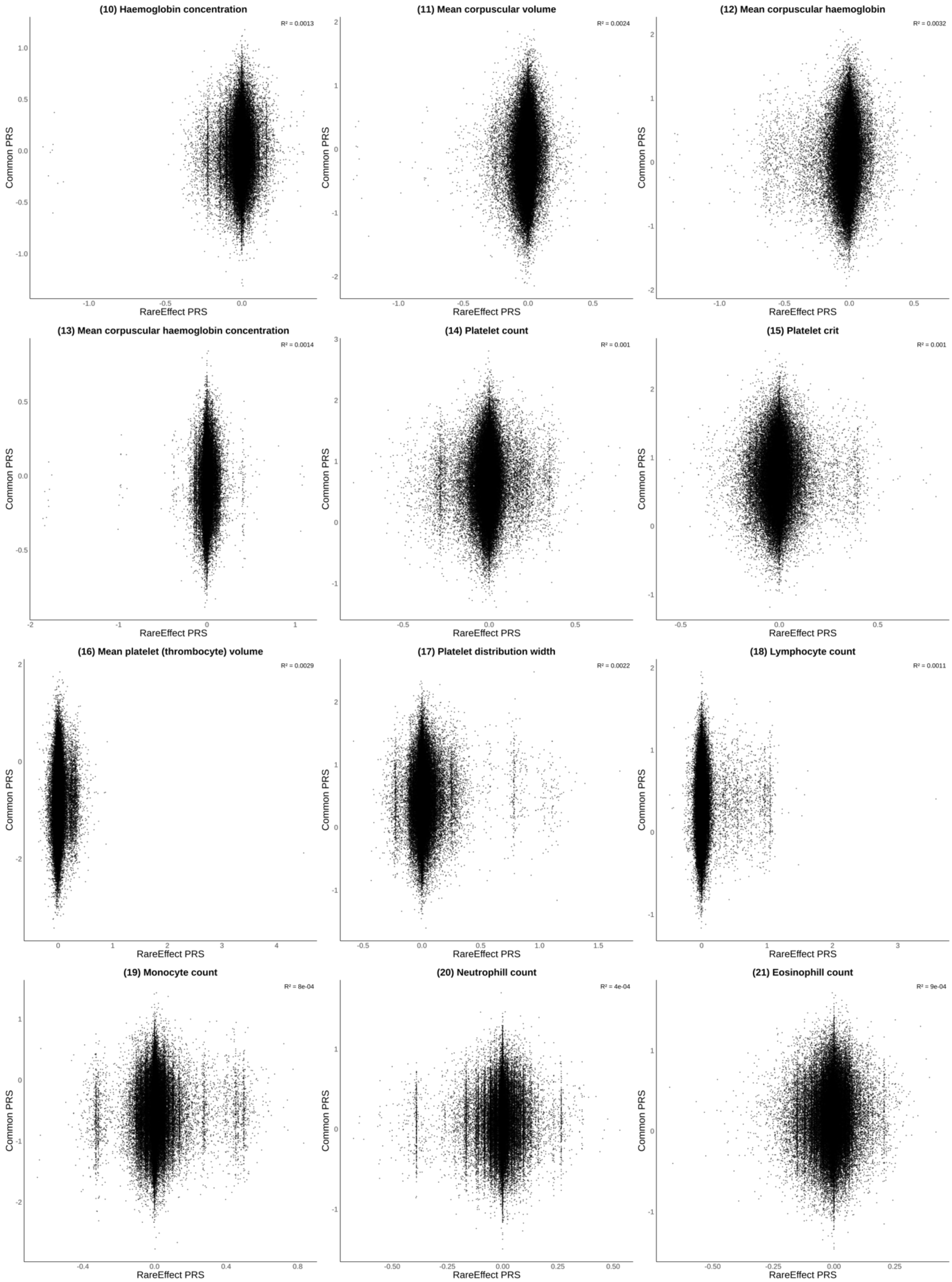
(10) Haemoglobin concentration
R² = 0.0013
(11) Mean corpuscular volume
R² = 0.0024
(12) Mean corpuscular haemoglobin
R² = 0.0032
(13) Mean corpuscular haemoglobin concentration
R² = 0.0014
(14) Platelet count
R² = 0.001
(15) Platelet crit
R² = 0.001
(16) Mean platelet (thrombocyte) volume
R² = 0.0029
(17) Platelet distribution width
R² = 0.0022
(18) Lymphocyte count
R² = 0.0011
(19) Monocyte count
R² = 8e-04
(20) Neutrophill count
R² = 4e-04
(21) Eosinophill count
R² = 9e-04
Common PRS
RareEffect PRS

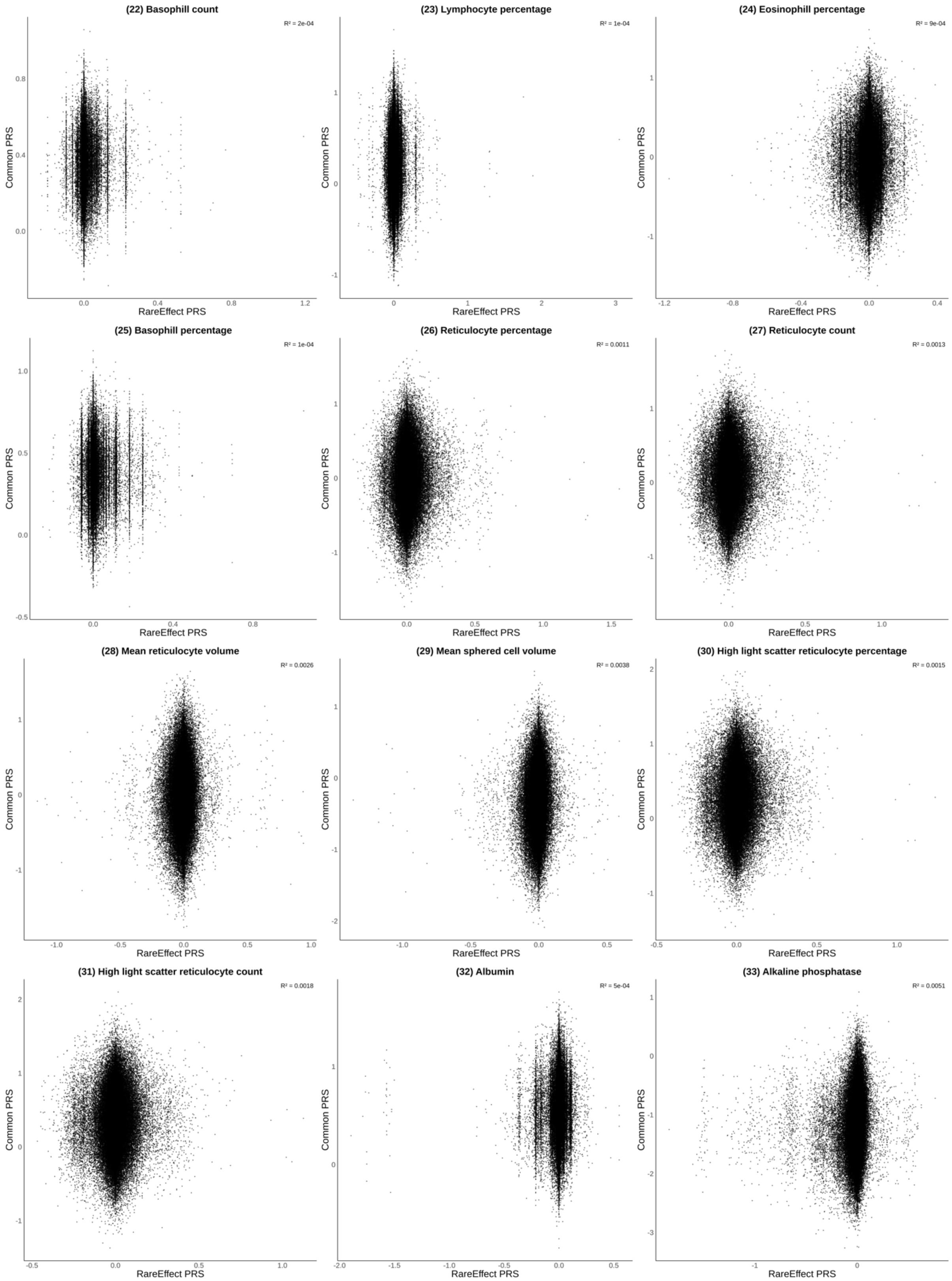
(22) Basophill count
R² = 2e-04
(23) Lymphocyte percentage
R² = 1e-04
(24) Eosinophill percentage
R² = 9e-04
(25) Basophill percentage
R² = 1e-04
(26) Reticulocyte percentage
R² = 0.0011
(27) Reticulocyte count
R² = 0.0013
(28) Mean reticulocyte volume
R² = 0.0026
(29) Mean sphered cell volume
R² = 0.0038
(30) High light scatter reticulocyte percentage
R² = 0.0015
(31) High light scatter reticulocyte count
R² = 0.0018
(32) Albumin
R² = 5e-04
(33) Alkaline phosphatase
R² = 0.0051
Common PRS
RareEffect PRS

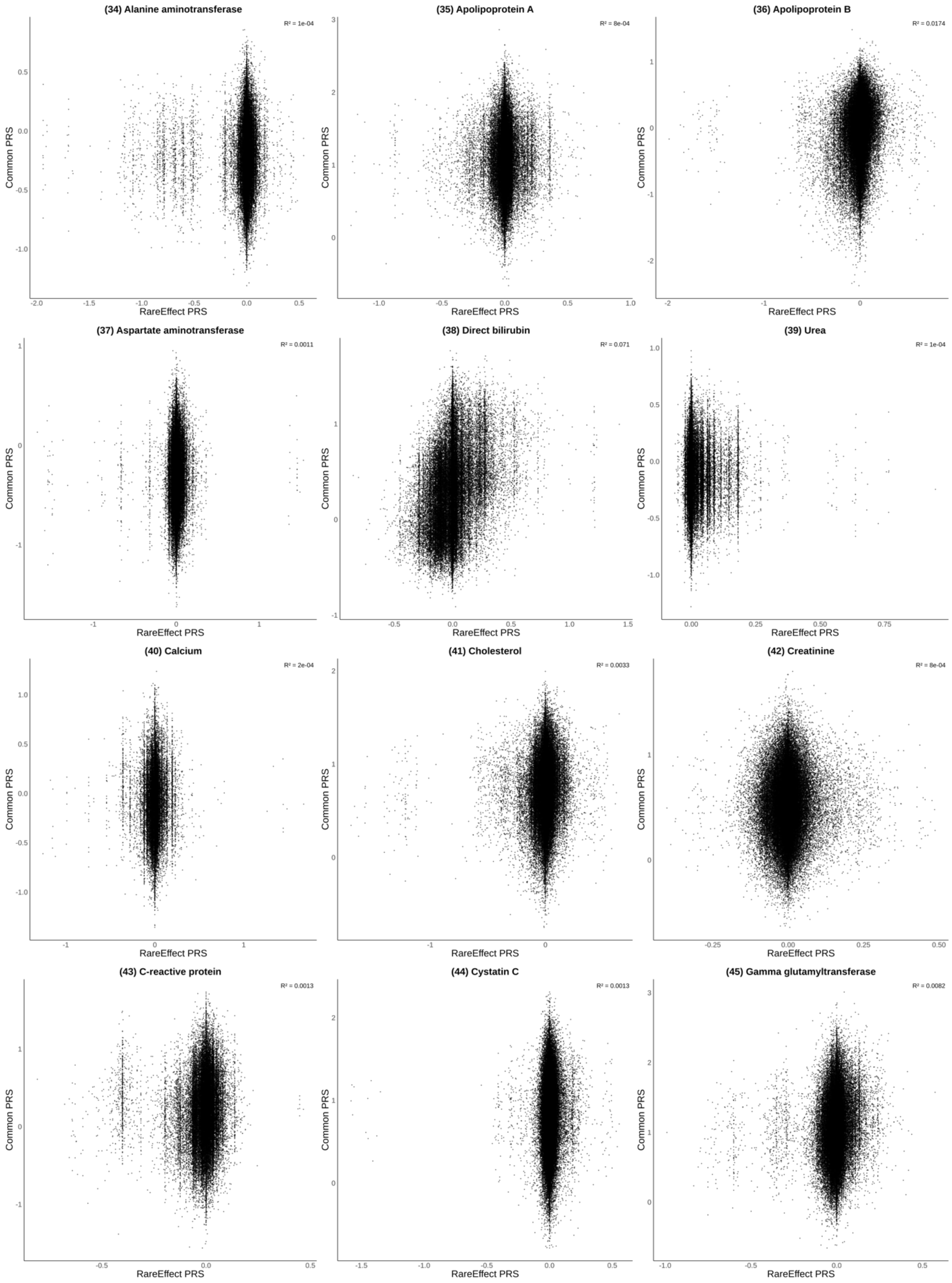
(34) Alanine aminotransferase
R² = 1e-04
(35) Apolipoprotein A
R² = 8e-04
(36) Apolipoprotein B
R² = 0.0174
(37) Aspartate aminotransferase
R² = 0.0011
(38) Direct bilirubin
R² = 0.071
(39) Urea
R² = 1e-04
(40) Calcium
R² = 2e-04
(41) Cholesterol
R² = 0.0033
(42) Creatinine
R² = 8e-04
(43) C-reactive protein
R² = 0.0013
(44) Cystatin C
R² = 0.0013
(45) Gamma glutamyltransferase
R² = 0.0082
Common PRS
RareEffect PRS

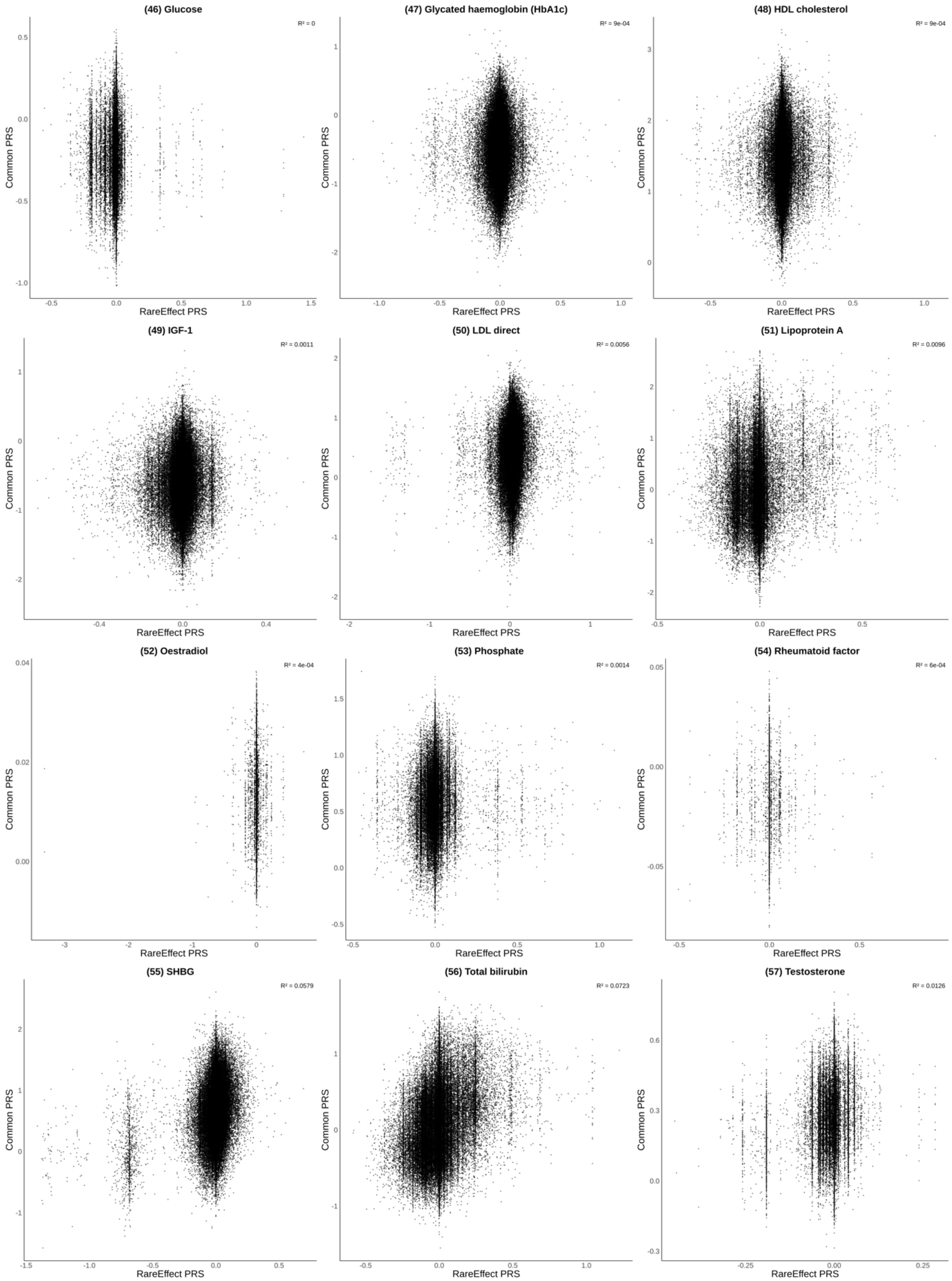
(46) Glucose
R² = 0
(47) Glycated haemoglobin (HbA1c)
R² = 9e-04
(48) HDL cholesterol
R² = 9e-04
(49) IGF-1
R² = 0.0011
(50) LDL direct
R² = 0.0056
(51) Lipoprotein A
R² = 0.0096
(52) Oestradiol
R² = 4e-04
(53) Phosphate
R² = 0.0014
(54) Rheumatoid factor
R² = 6e-04
(55) SHBG
R² = 0.0579
(56) Total bilirubin
R² = 0.0723
(57) Testosterone
R² = 0.0126
Common PRS
RareEffect PRS

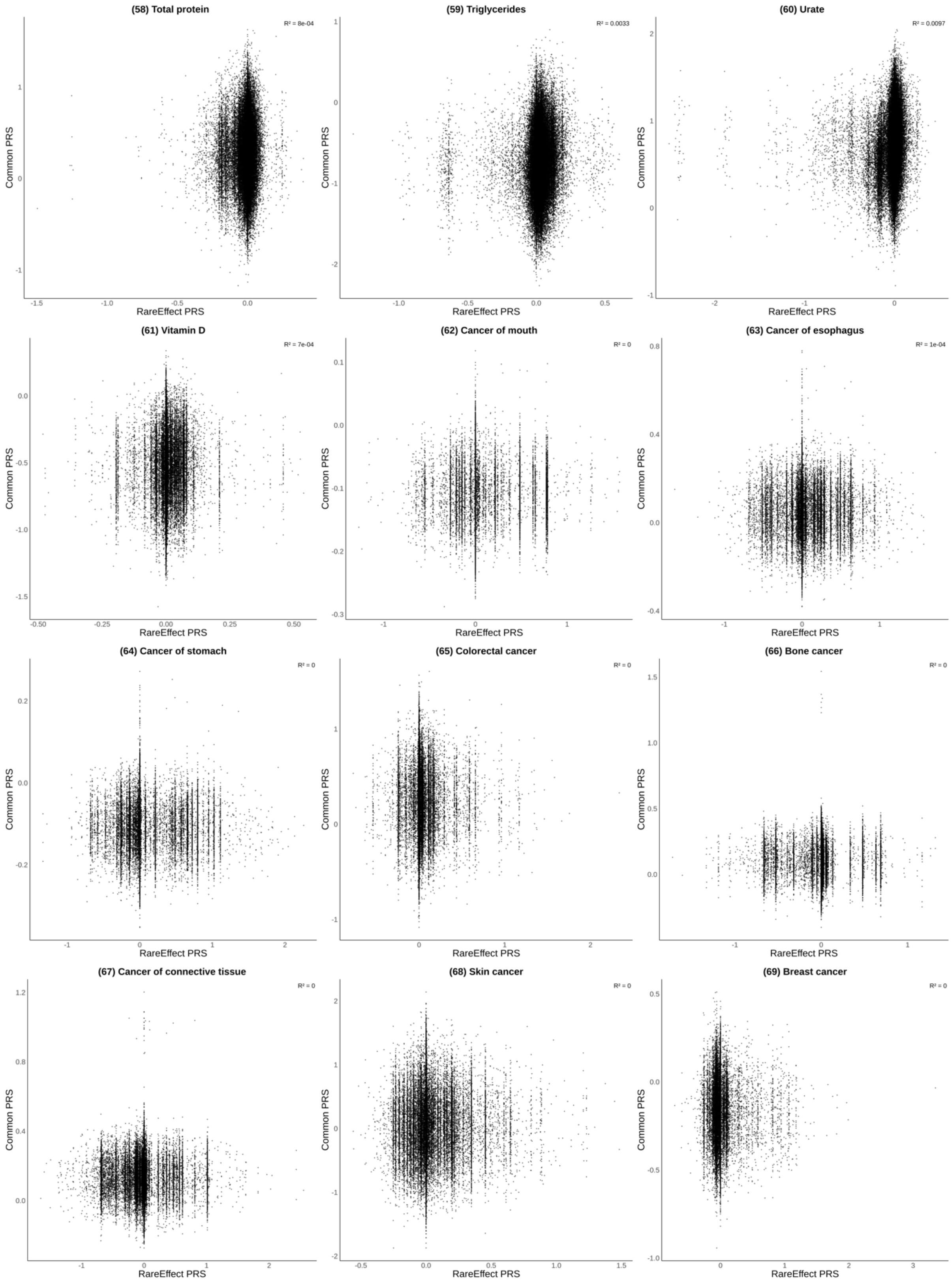
(58) Total protein
R² = 8e-04
(59) Triglycerides
R² = 0.0033
(60) Urate
R² = 0.0097
(61) Vitamin D
R² = 7e-04
(62) Cancer of mouth
R² = 0
(63) Cancer of esophagus
R² = 1e-04
(64) Cancer of stomach
R² = 0
(65) Colorectal cancer
R² = 0
(66) Bone cancer
R² = 0
(67) Cancer of connective tissue
R² = 0
(68) Skin cancer
R² = 0
(69) Breast cancer
R² = 0
Common PRS
RareEffect PRS

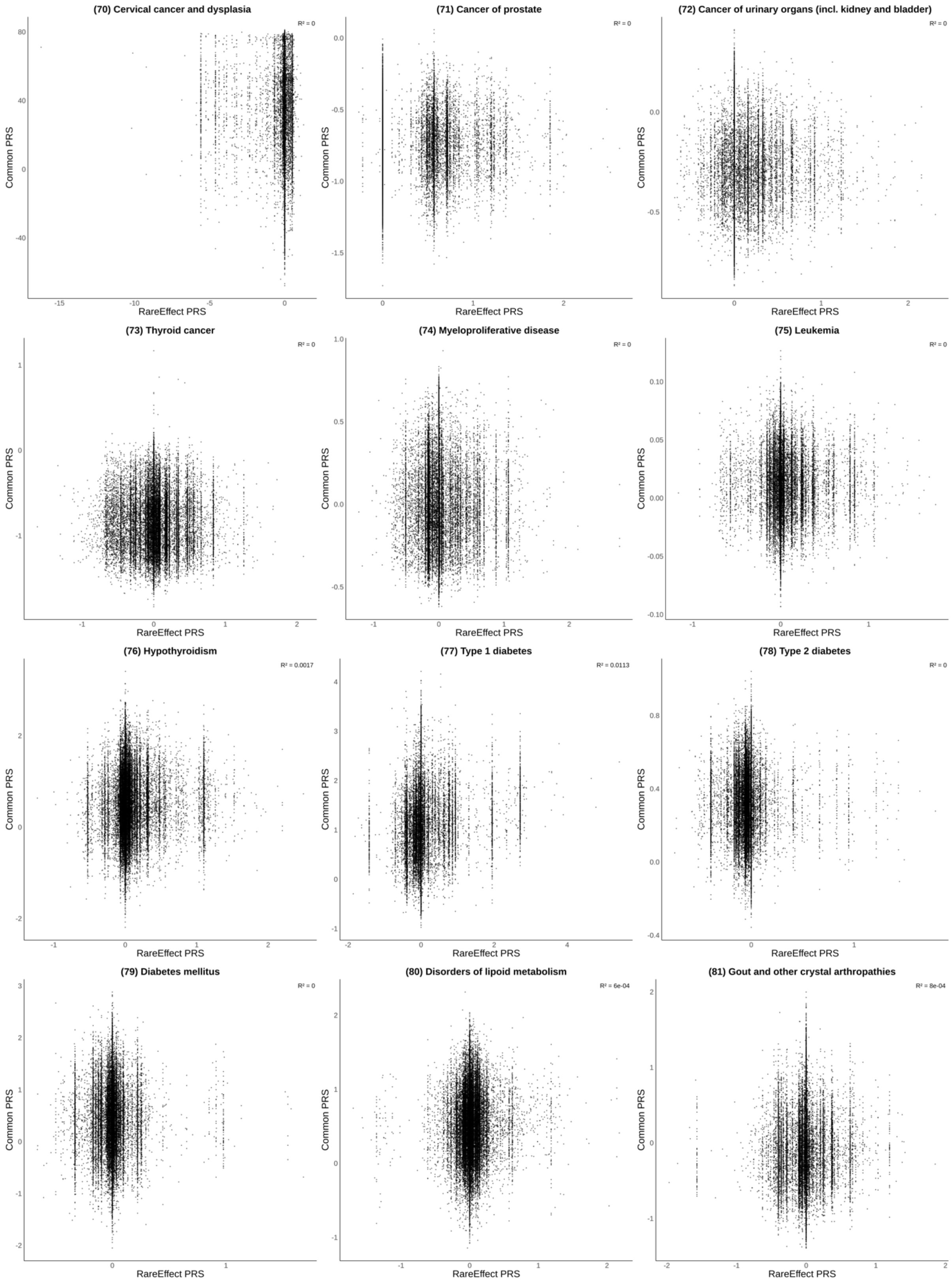
(70) Cervical cancer and dysplasia
R² = 0
(71) Cancer of prostate
R² = 0
(72) Cancer of urinary organs (incl. kidney and bladder)
R² = 0
(73) Thyroid cancer
R² = 0
(74) Myeloproliferative disease
R² = 0
(75) Leukemia
R² = 0
(76) Hypothyroidism
R² = 0.0017
(77) Type 1 diabetes
R² = 0.0113
(78) Type 2 diabetes
R² = 0
(79) Diabetes mellitus
R² = 0
(80) Disorders of lipoid metabolism
R² = 6e-04
(81) Gout and other crystal arthropathies
R² = 8e-04
Common PRS
RareEffect PRS

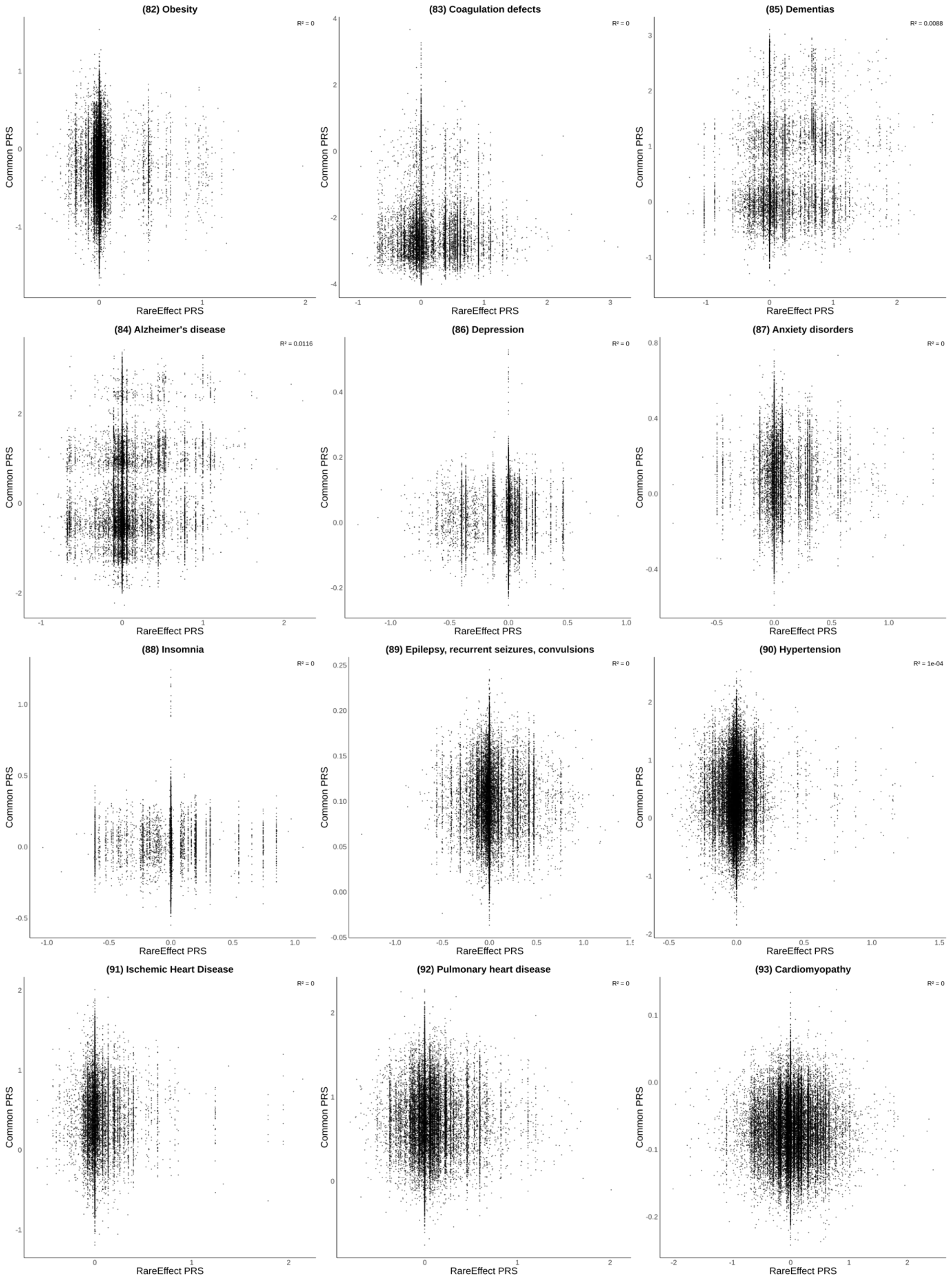
(82) Obesity
R² = 0
(83) Coagulation defects
R² = 0
(85) Dementias
R² = 0.0088
(84) Alzheimer's disease
R² = 0.0116
(86) Depression
R² = 0
(87) Anxiety disorders
R² = 0
(88) Insomnia
R² = 0
(89) Epilepsy, recurrent seizures, convulsions
R² = 0
(90) Hypertension
R² = 1e-04
(91) Ischemic Heart Disease
R² = 0
(92) Pulmonary heart disease
R² = 0
(93) Cardiomyopathy
R² = 0
Common PRS
RareEffect PRS

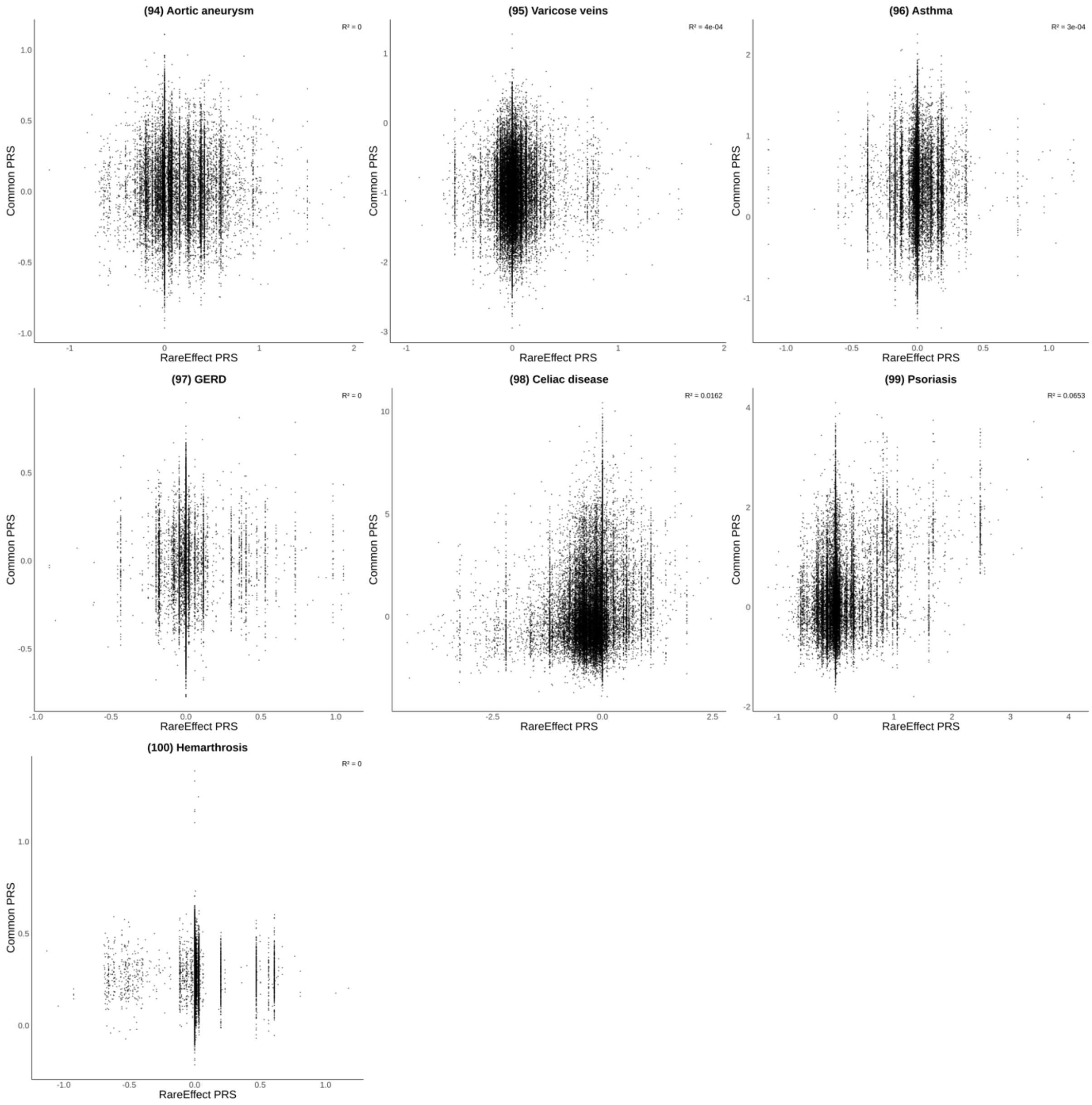
(94) Aortic aneurysm
R² = 0
(95) Varicose veins
R² = 4e-04
(96) Asthma
R² = 3e-04
(97) GERD
R² = 0
(98) Celiac disease
R² = 0.0162
(99) Psoriasis
R² = 0.0653
(100) Hemarthrosis
R² = 0
Common PRS
RareEffect PRS

**Supplementary Figure 4. Comparison of the performance of rare variant PRS methods for lipid phenotypes**

Performance was assessed using the coefficient of determination ($R^2$) of the risk prediction models. We evaluated the $R^2$ of each model within five subgroups: (1) all samples, (2) samples with top/bottom 10% PRS, (3) samples with top/bottom 5% PRS, (4) samples with top/bottom 1% PRS, and (5) samples with top/bottom 0.5% PRS. The black vertical lines represent the 95% confidence interval of the $R^2$ estimates.

**(A) HDL Cholesterol**

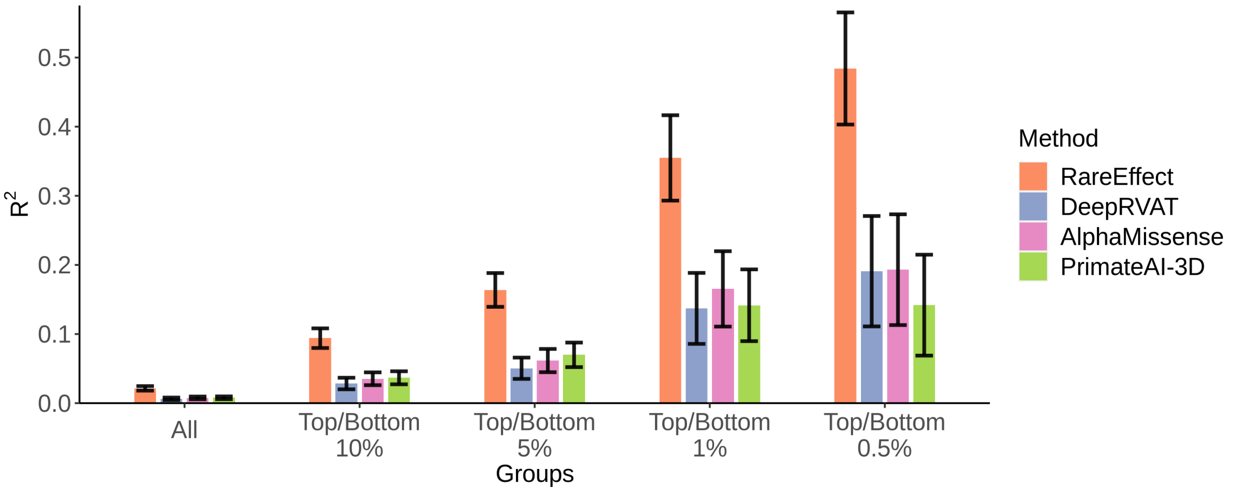


**(B) LDL Cholesterol**

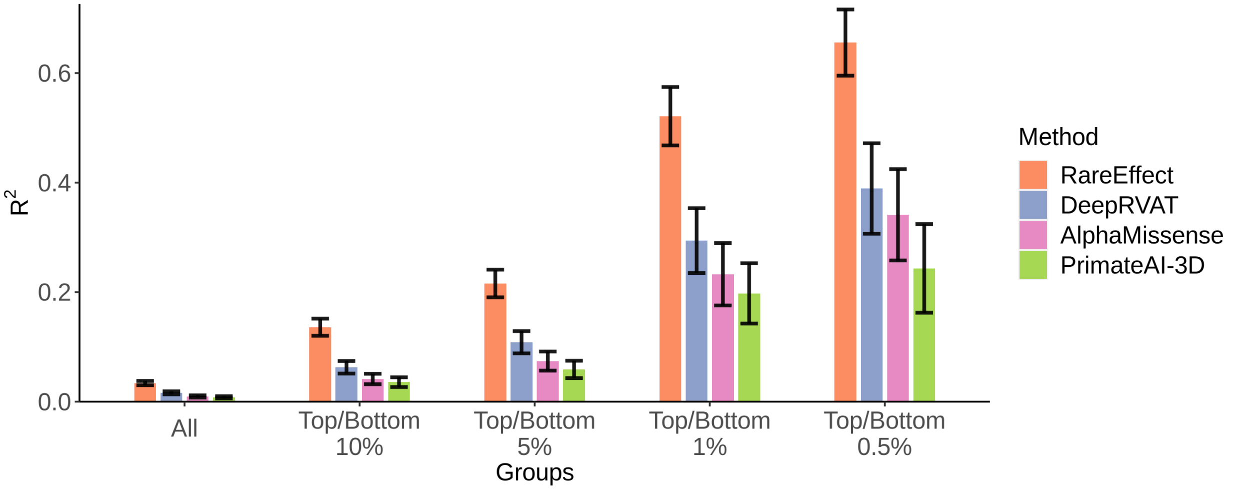

### (C) Triglycerides

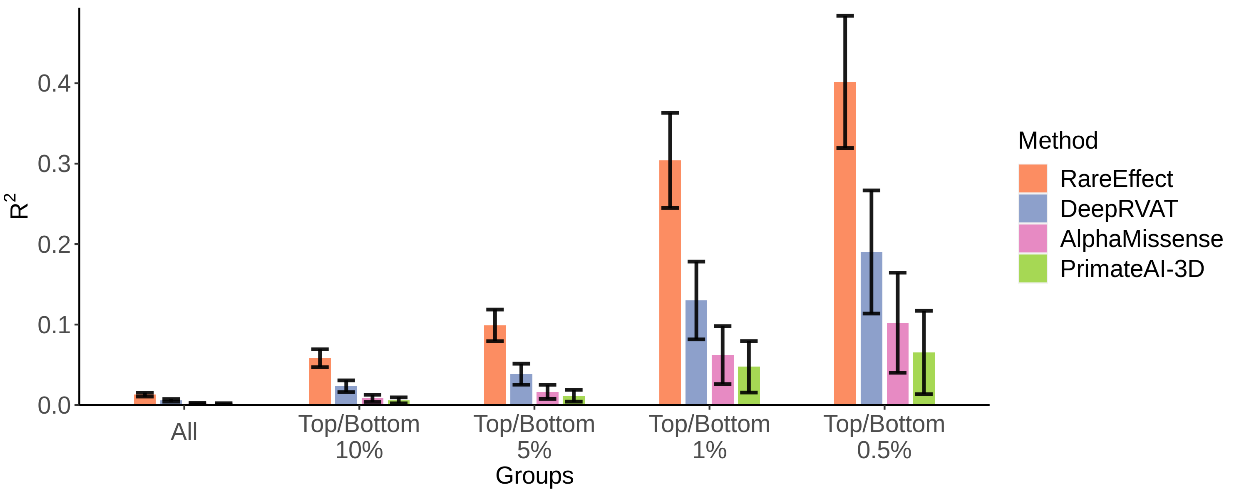

**Supplementary Figure 5. Comparison of the performance of rare variant PRS methods when combined with common variant PRS (SBayesRC) for lipid phenotypes**

Performance was assessed using the coefficient of determination ($R^2$) of the risk prediction models. We evaluated the $R^2$ of each model within five subgroups: (1) all samples, (2) samples with top/bottom 10% PRS, (3) samples with top/bottom 5% PRS, (4) samples with top/bottom 1% PRS, and (5) samples with top/bottom 0.5% PRS. The black vertical lines represent the 95% confidence interval of the $R^2$ estimates.

**(A) HDL Cholesterol**

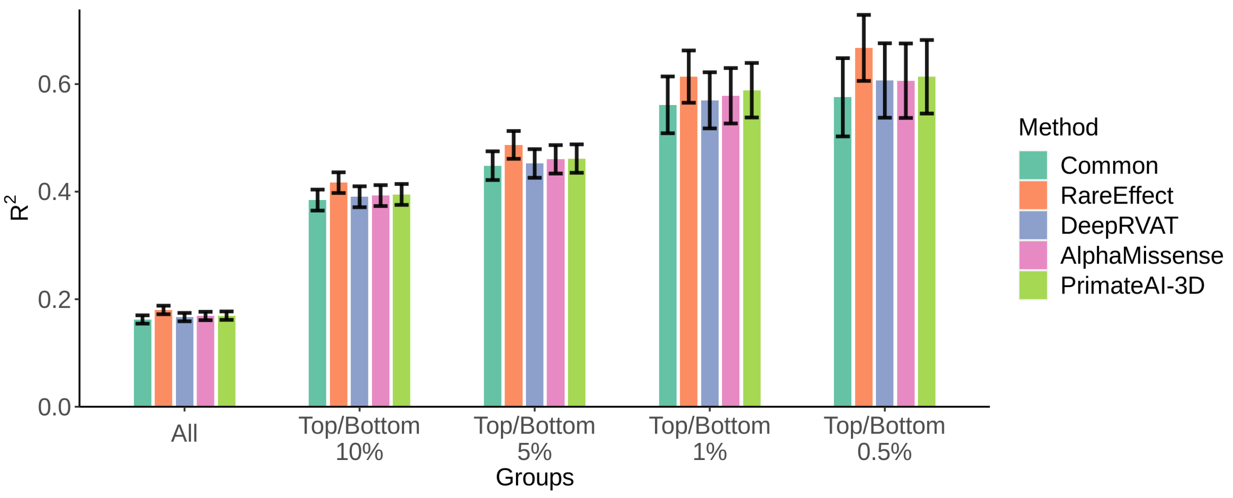


**(B) LDL Cholesterol**

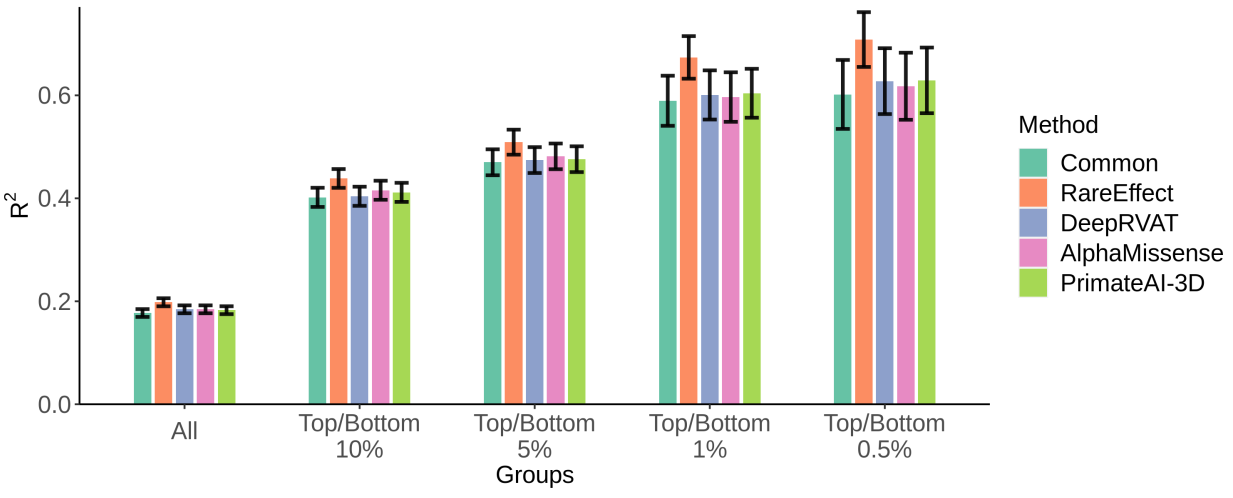

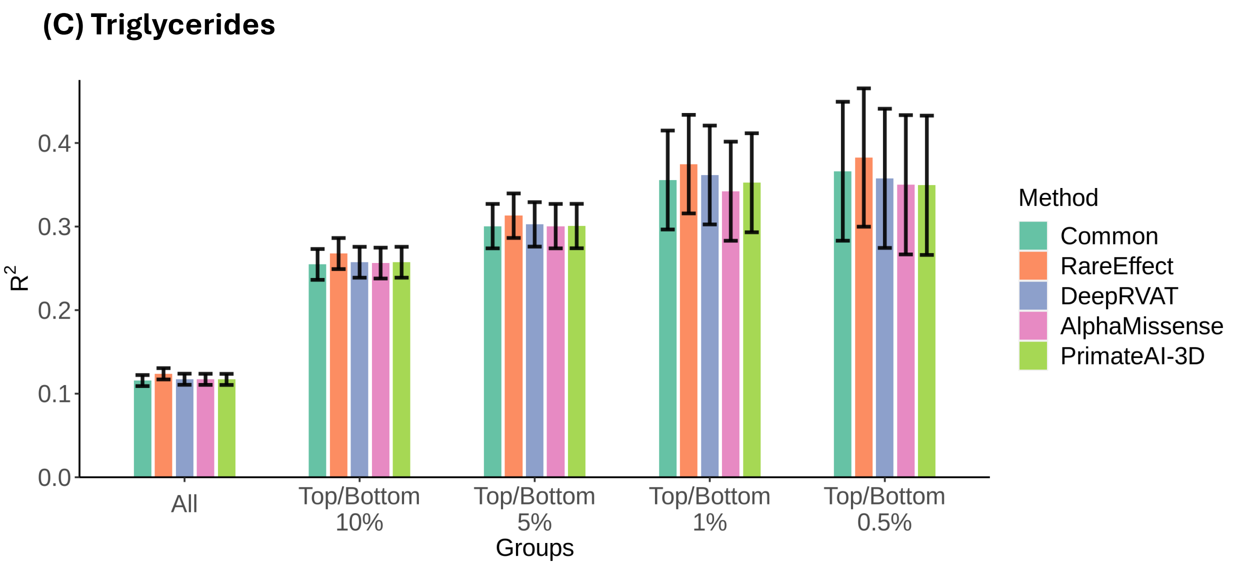

(C) Triglycerides
R2
0.0
0.1
0.2
0.3
0.4
All
Top/Bottom 10%
Top/Bottom 5%
Top/Bottom 1%
Top/Bottom 0.5%
Groups
Method
Common
RareEffect
DeepRVAT
AlphaMissense
PrimateAI-3D

**Supplementary Figure 6. Enrichment of high-risk individuals based on RareEffect PRS ($PRS_{RE}$), common variant PRS ($PRS_{common}$) and the composite score among phenotype outliers.**

The phenotype outliers were defined as samples with phenotype value exceeding a z-score threshold ($x$-axis). The $y$-axis shows the enrichment of PRS of the phenotype outliers compared to the baseline population, which is defined as the individuals with phenotype value less than zero. Lines are color-coded based on their PRS type: blue for $PRS_{common}$, orange for composite score, and green for $PRS_{RE}$. The solid line indicates the 99% quantile (top 1%) in each score, and the dashed line indicated indicates the 99.9% quantile (top 0.1%) in each score.

**(A) Height**

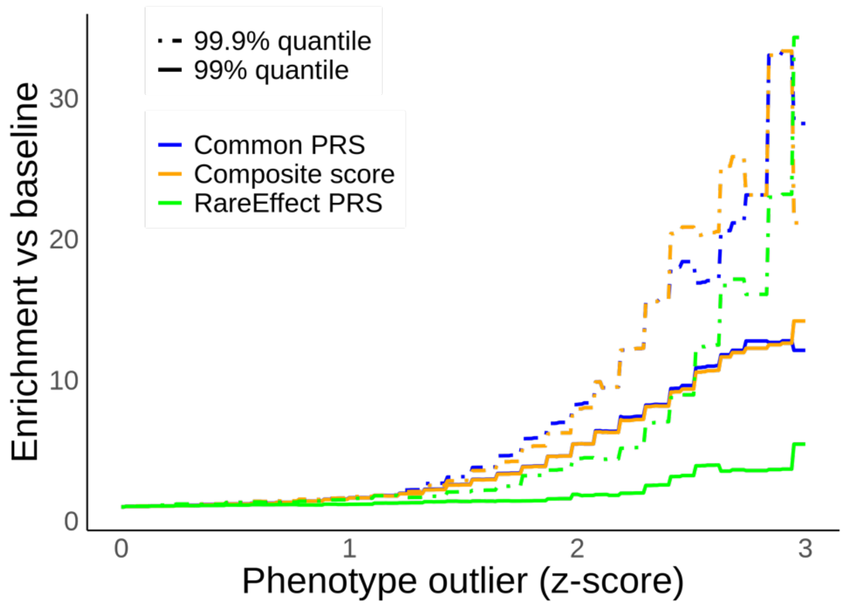

**(B) Body Mass Index (BMI)**

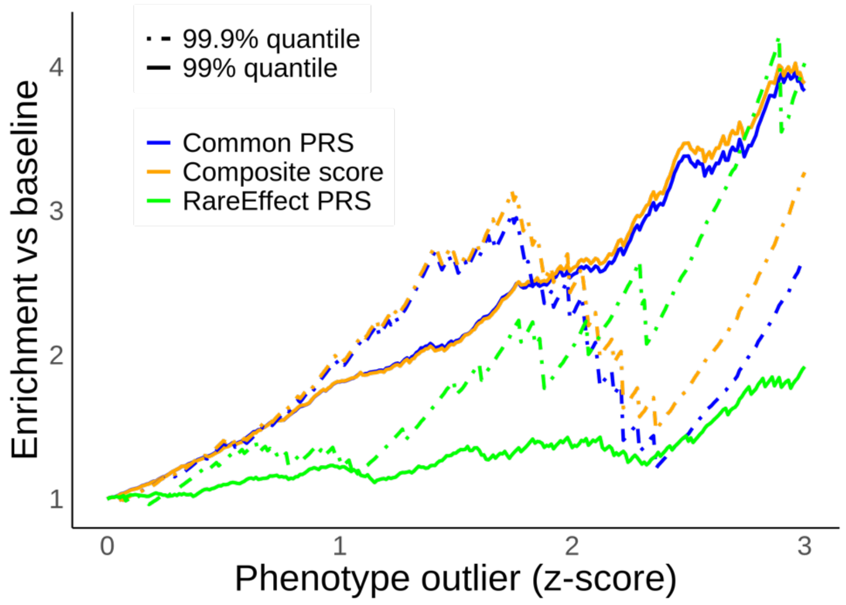


**(C) HDL Cholesterol**

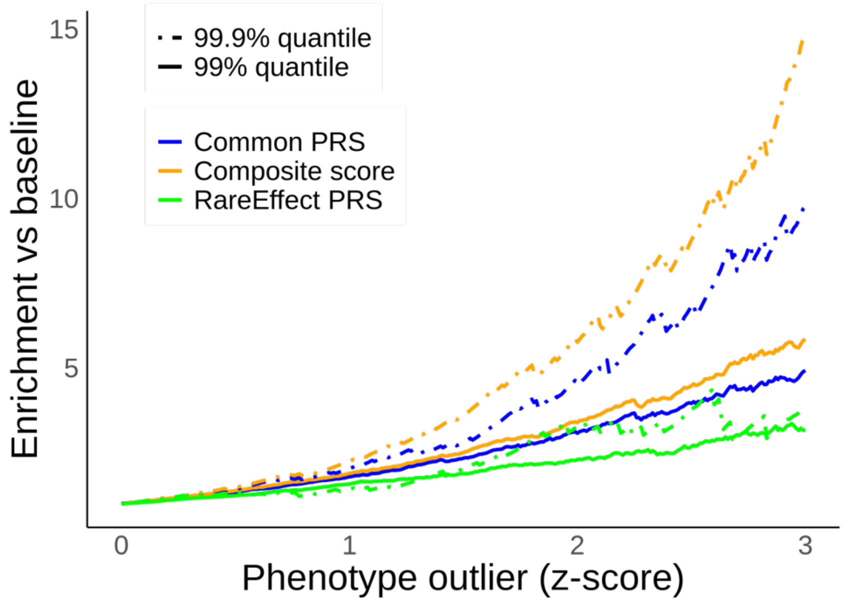

**(D) LDL Cholesterol**

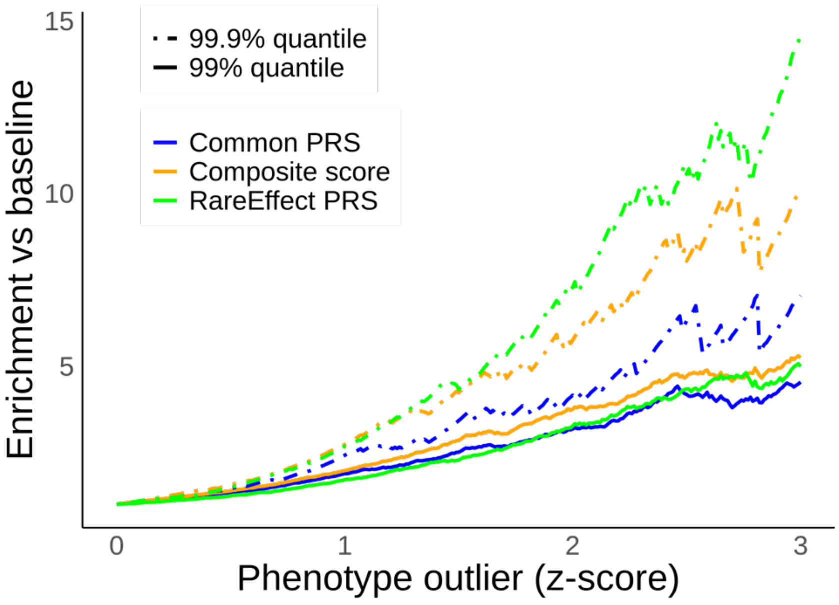


**(E) Triglycerides**

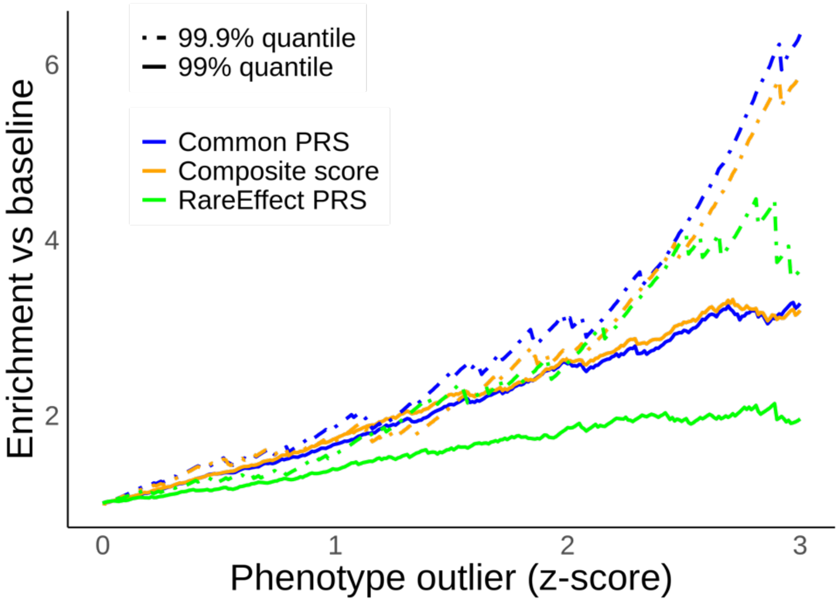


**(e) Triglycerides**

**Supplementary Figure 7. Computation time of RareEffect, linear regression, and ridge regression.**

Computation time was evaluated using an AMD EPYC 7542 CPU (2.9 GHz, 32 cores). Computation time for linear regression and ridge regression was measured using lm and glm functions in R 4.3.2, respectively.

**(A) Computation time in seconds (per gene) by number of variants in a gene measured using White British in UK Biobank 200K WES data (n=166,960)**

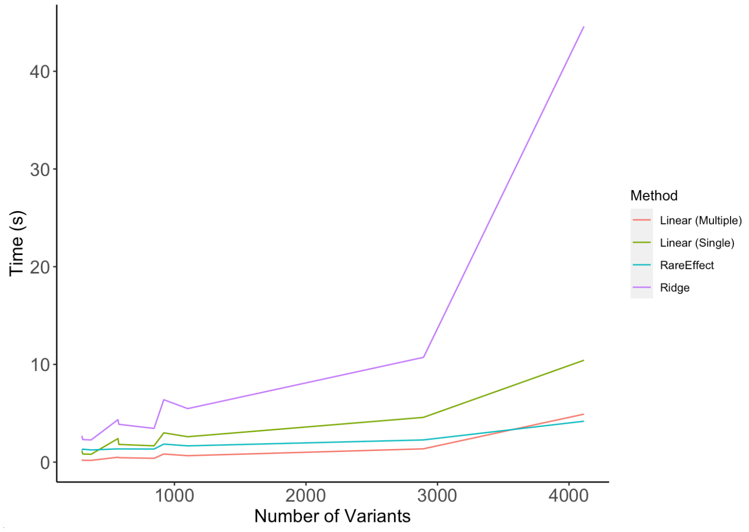


**(B) Computation time in seconds (per gene) by number of samples measured using a gene with 1,101 variants**

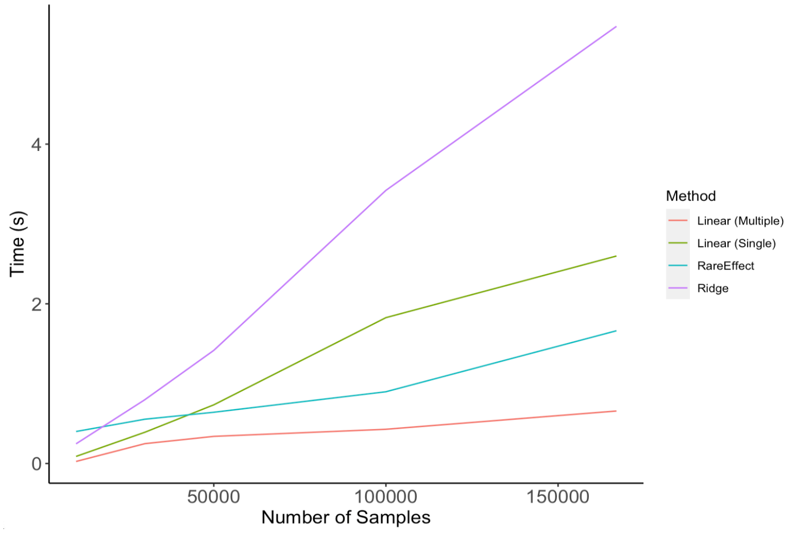

**Supplementary Figure 8. Memory usage of RareEffect by number of variants (UKB 200K WES, n = 166,960).**

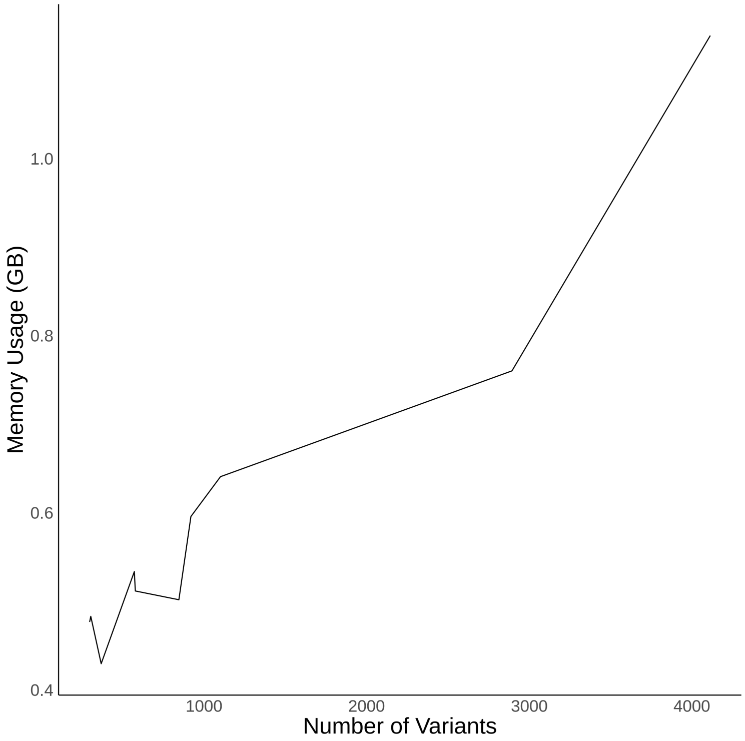

**Supplementary Figure 9. Computation time of the fast implementation Firth bias correction and the standard Firth correction.**

Computation time was evaluated using an AMD EPYC 7542 CPU (2.9 GHz, 32 cores).

**(A) Computation time for a single gene using 342,409 individuals by the number of variants**

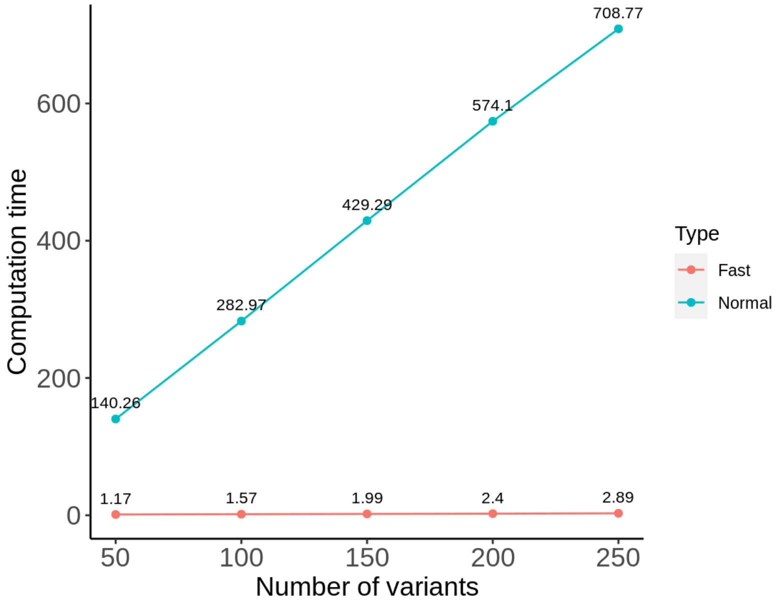


**(B) Computation time for a single gene with 100 variants by the number of samples**

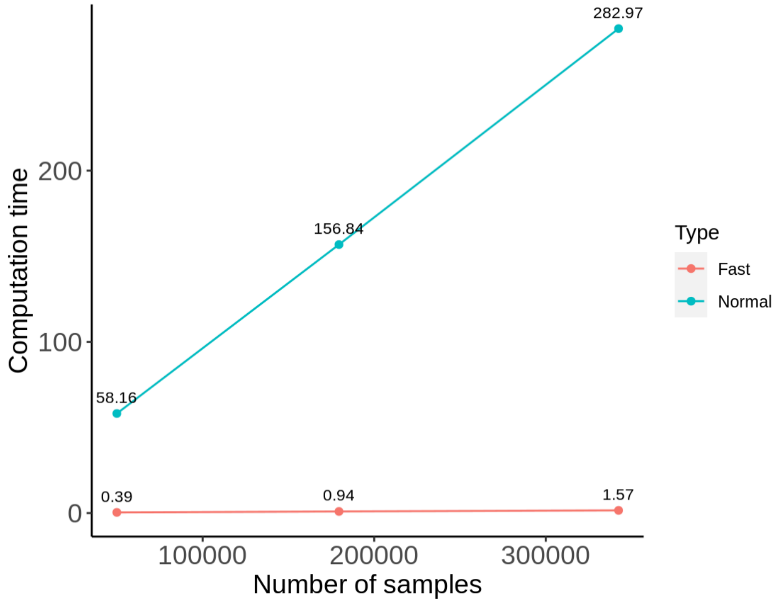

**Supplementary Figure 10. Relationship between RareEffect PRS ($PRS_{RE}$) and phenotype values.**

The $x$-axis represents $PRS_{RE}$ of each individuals, and the $y$-axis represents the log-transformed phenotype value.

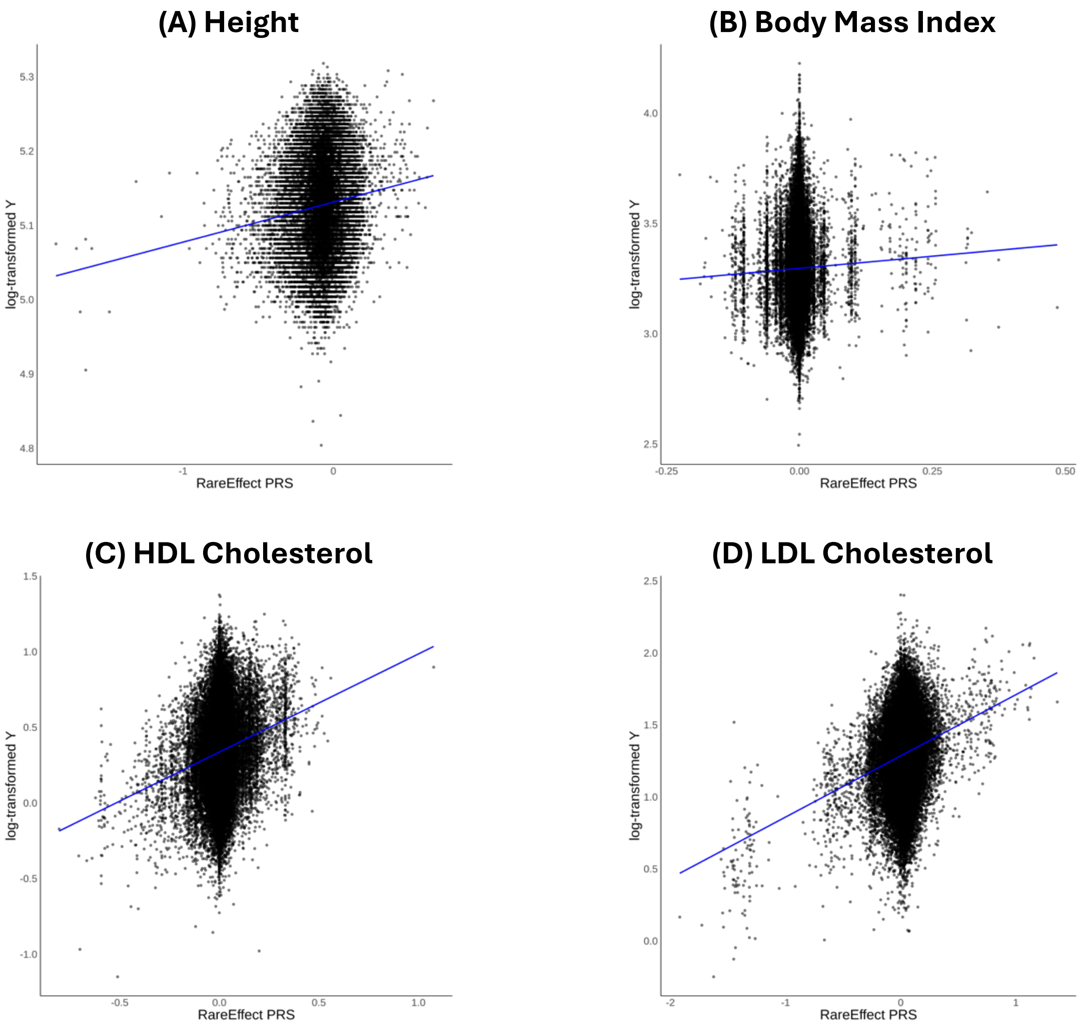

### (E) Triglycerides

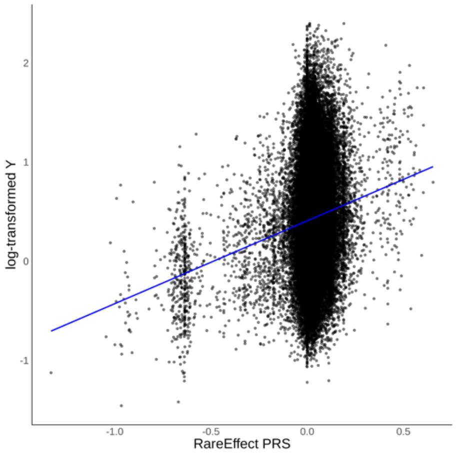

**Supplementary Figure 11. Comparison of estimated variance components between RareEffect and the MoM estimator.**

The $x$-axis represents the estimated variance components (likelihood-based) using FaST-LMM, and the $y$-axis represents the MoM estimates of variance components, which may include negative values.

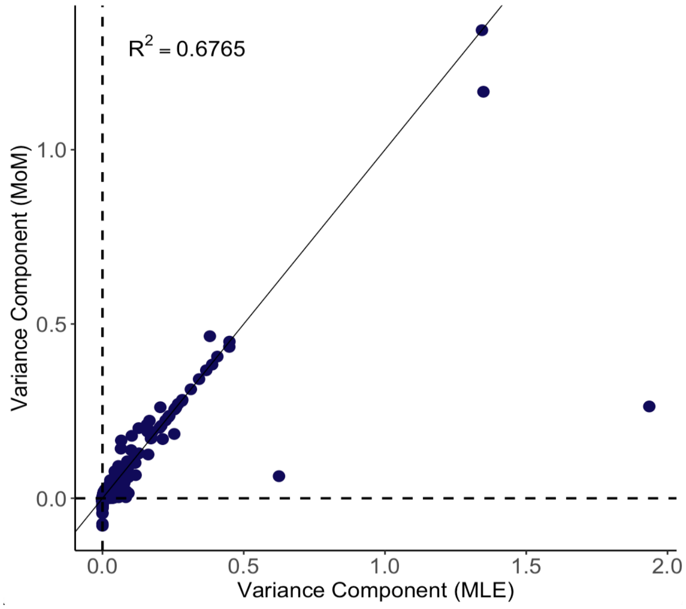

**Supplementary Figure 12. Comparison of estimated effect sizes between computing the hat matrix at every iteration and computing it only once during the Firth bias correction (estimated using UKB 470K WES data).**

The $x$-axis represents the estimated effect size when computing the hat matrix at every iteration, and the $y$-axis represents the estimated effect size when computing the hat matrix only once during the Firth bias correction step.

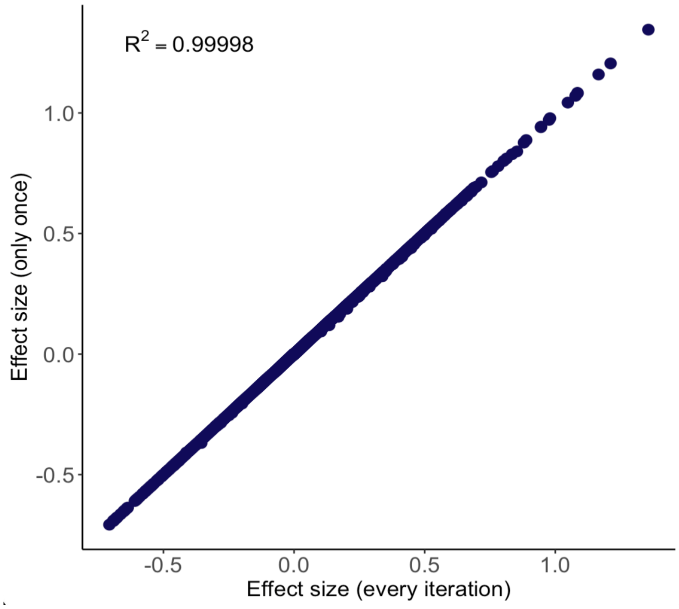

### Supplementary Figure 13. Distribution of RareEffect PRS

The $x$-axis represents $PRS_{RE}$ of each individuals, and the $y$-axis represents the frequency. Individuals with zero $PRS_{RE}$ were excluded from the histogram, and the number of zero-score samples is indicated in the top-right corner.

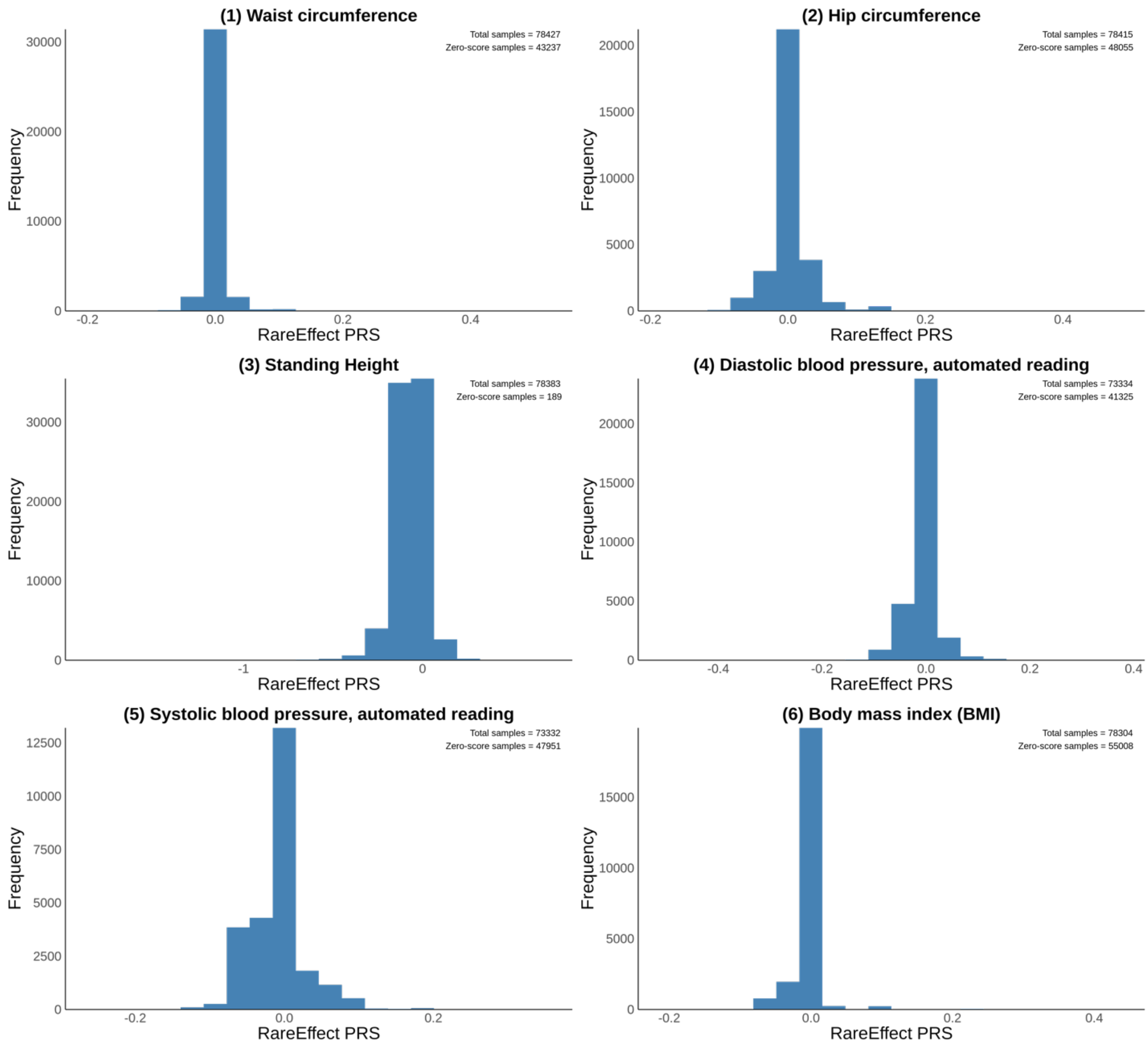

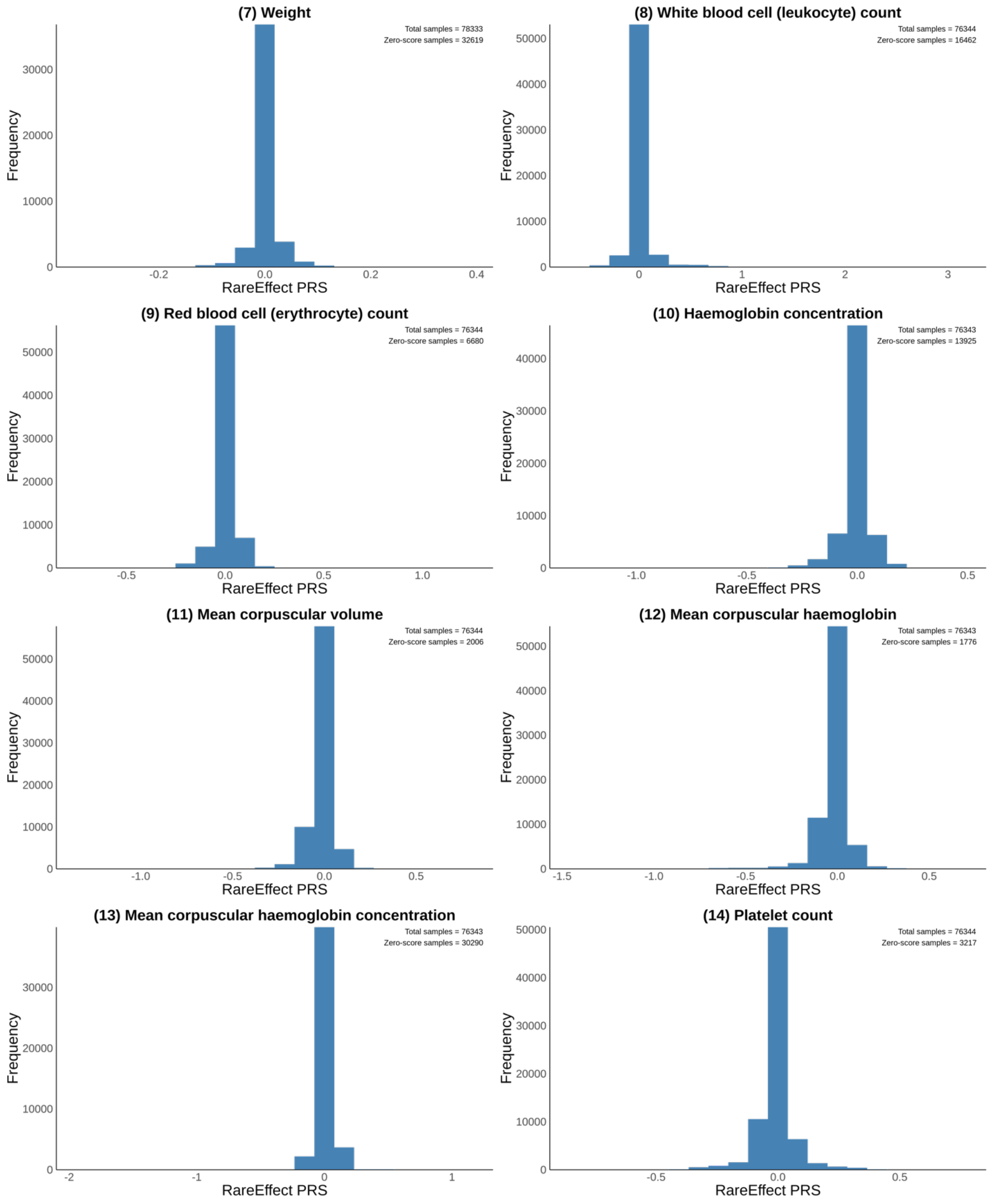
(7) Weight
Total samples = 78333
Zero-score samples = 32619
Frequency
RareEffect PRS
(8) White blood cell (leukocyte) count
Total samples = 76344
Zero-score samples = 16462
Frequency
RareEffect PRS
(9) Red blood cell (erythrocyte) count
Total samples = 76344
Zero-score samples = 6680
Frequency
RareEffect PRS
(10) Haemoglobin concentration
Total samples = 76343
Zero-score samples = 13925
Frequency
RareEffect PRS
(11) Mean corpuscular volume
Total samples = 76344
Zero-score samples = 2006
Frequency
RareEffect PRS
(12) Mean corpuscular haemoglobin
Total samples = 76343
Zero-score samples = 1776
Frequency
RareEffect PRS
(13) Mean corpuscular haemoglobin concentration
Total samples = 76343
Zero-score samples = 30290
Frequency
RareEffect PRS
(14) Platelet count
Total samples = 76344
Zero-score samples = 3217
Frequency
RareEffect PRS

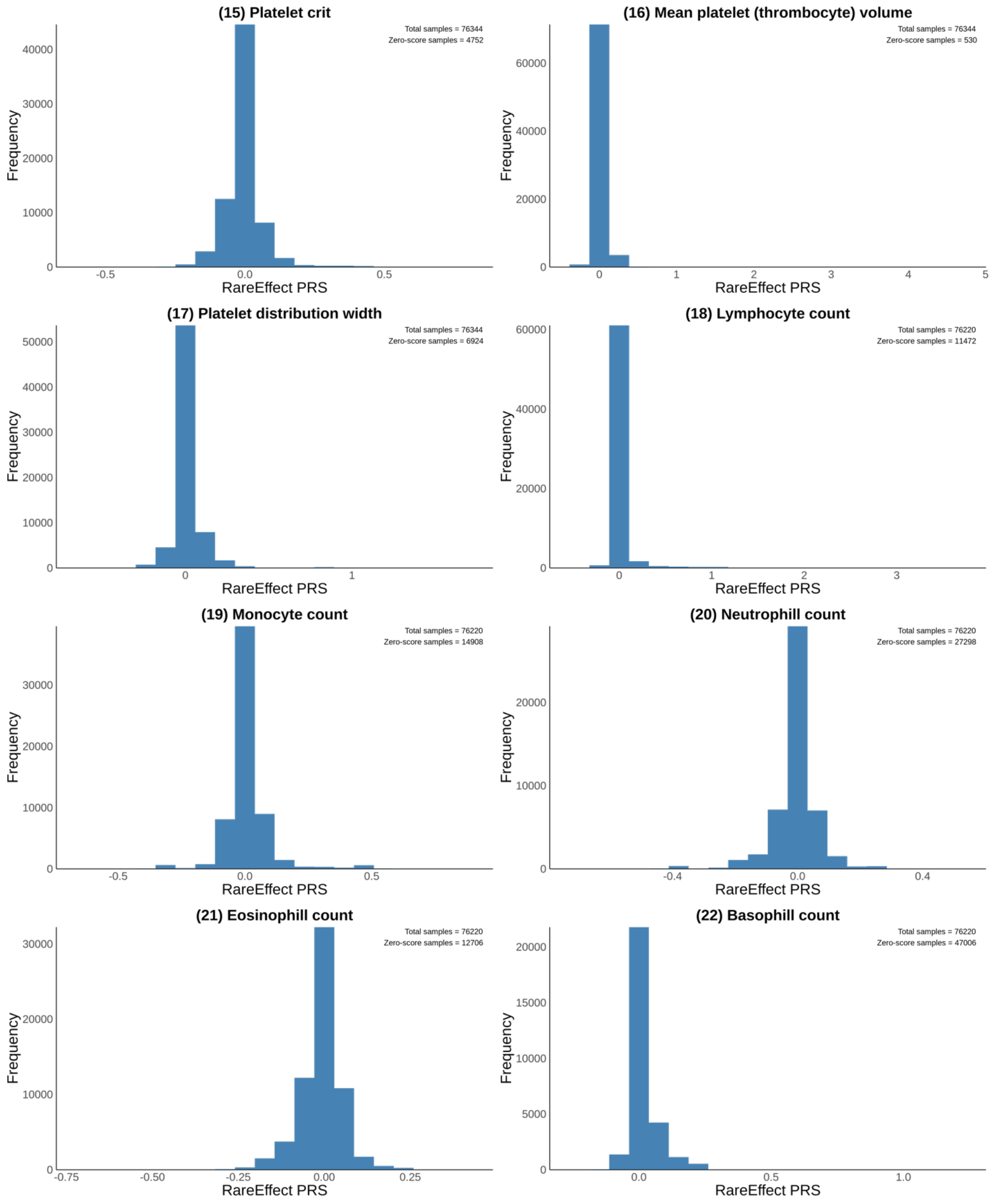

(15) Platelet crit
Total samples = 76344
Zero-score samples = 4752
Frequency
RareEffect PRS
(16) Mean platelet (thrombocyte) volume
Total samples = 76344
Zero-score samples = 530
Frequency
RareEffect PRS
(17) Platelet distribution width
Total samples = 76344
Zero-score samples = 6924
Frequency
RareEffect PRS
(18) Lymphocyte count
Total samples = 76220
Zero-score samples = 11472
Frequency
RareEffect PRS
(19) Monocyte count
Total samples = 76220
Zero-score samples = 14908
Frequency
RareEffect PRS
(20) Neutrophill count
Total samples = 76220
Zero-score samples = 27298
Frequency
RareEffect PRS
(21) Eosinophill count
Total samples = 76220
Zero-score samples = 12706
Frequency
RareEffect PRS
(22) Basophill count
Total samples = 76220
Zero-score samples = 47006
Frequency
RareEffect PRS

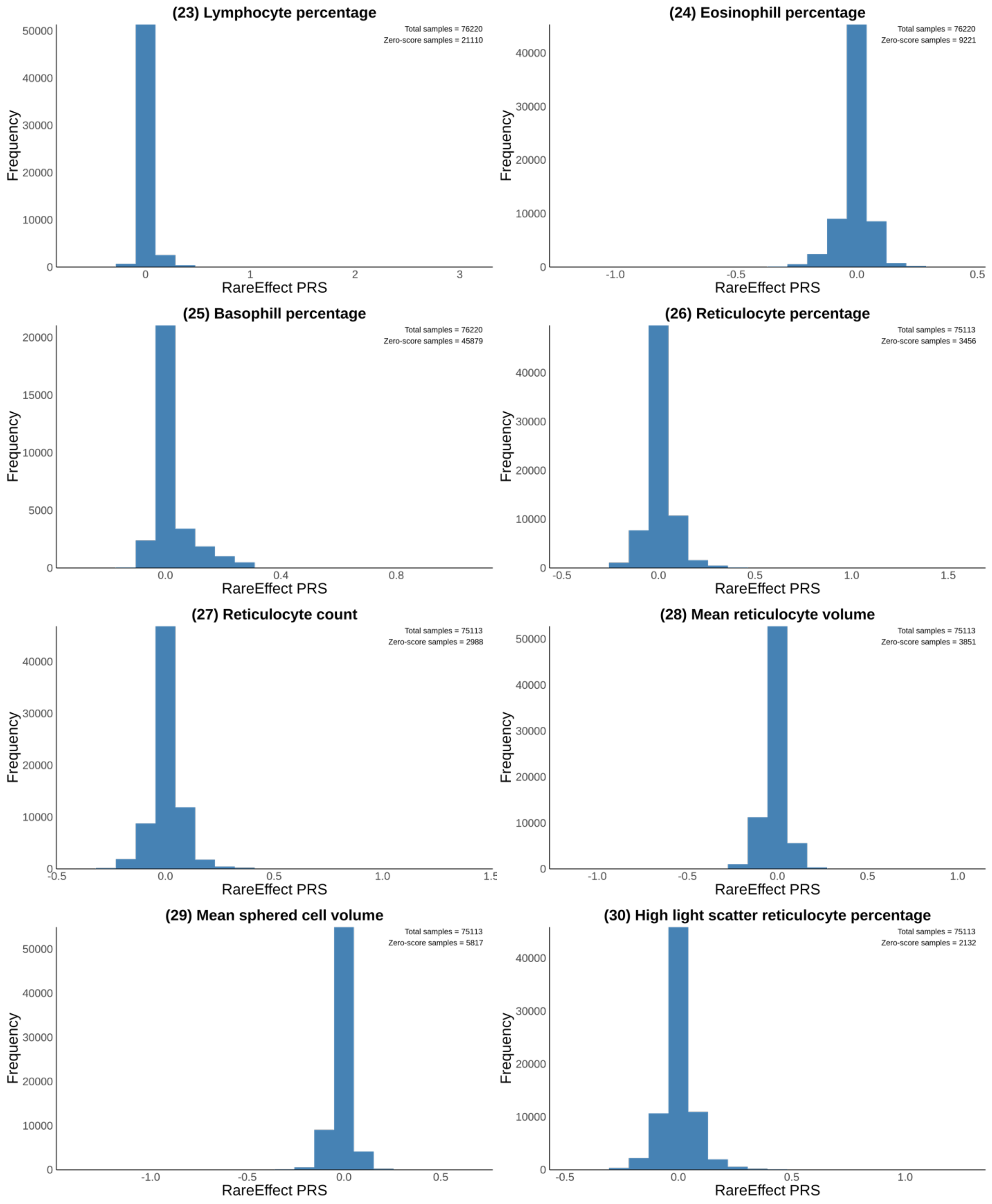
(23) Lymphocyte percentage
Total samples = 76220
Zero-score samples = 21110
(24) Eosinophill percentage
Total samples = 76220
Zero-score samples = 9221
(25) Basophill percentage
Total samples = 76220
Zero-score samples = 45879
(26) Reticulocyte percentage
Total samples = 75113
Zero-score samples = 3456
(27) Reticulocyte count
Total samples = 75113
Zero-score samples = 2988
(28) Mean reticulocyte volume
Total samples = 75113
Zero-score samples = 3851
(29) Mean sphered cell volume
Total samples = 75113
Zero-score samples = 5817
(30) High light scatter reticulocyte percentage
Total samples = 75113
Zero-score samples = 2132
Frequency
RareEffect PRS

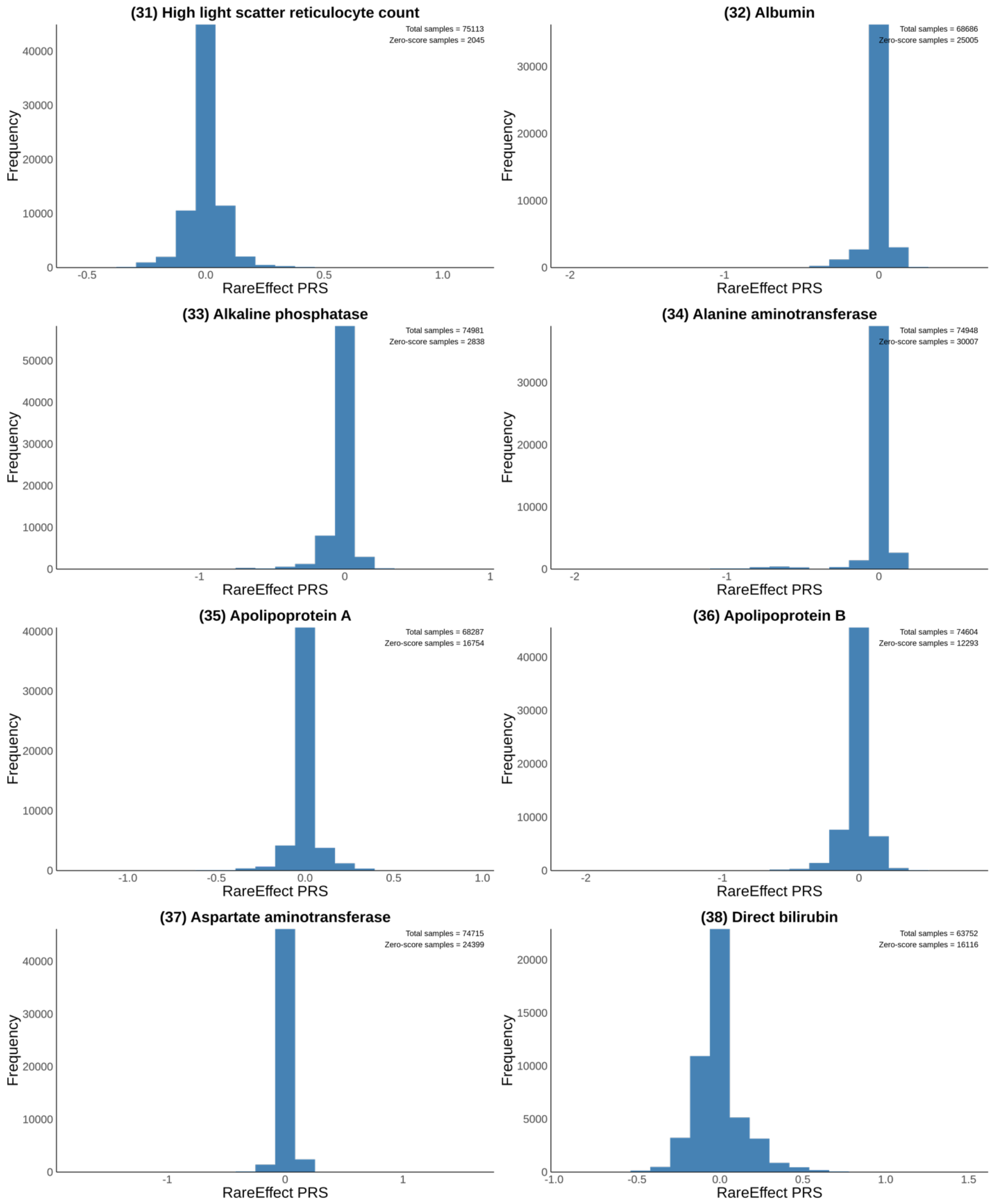
(31) High light scatter reticulocyte count
Total samples = 75113
Zero-score samples = 2045
(32) Albumin
Total samples = 68686
Zero-score samples = 25005
(33) Alkaline phosphatase
Total samples = 74981
Zero-score samples = 2838
(34) Alanine aminotransferase
Total samples = 74948
Zero-score samples = 30007
(35) Apolipoprotein A
Total samples = 68287
Zero-score samples = 16754
(36) Apolipoprotein B
Total samples = 74604
Zero-score samples = 12293
(37) Aspartate aminotransferase
Total samples = 74715
Zero-score samples = 24399
(38) Direct bilirubin
Total samples = 63752
Zero-score samples = 16116
Frequency
RareEffect PRS

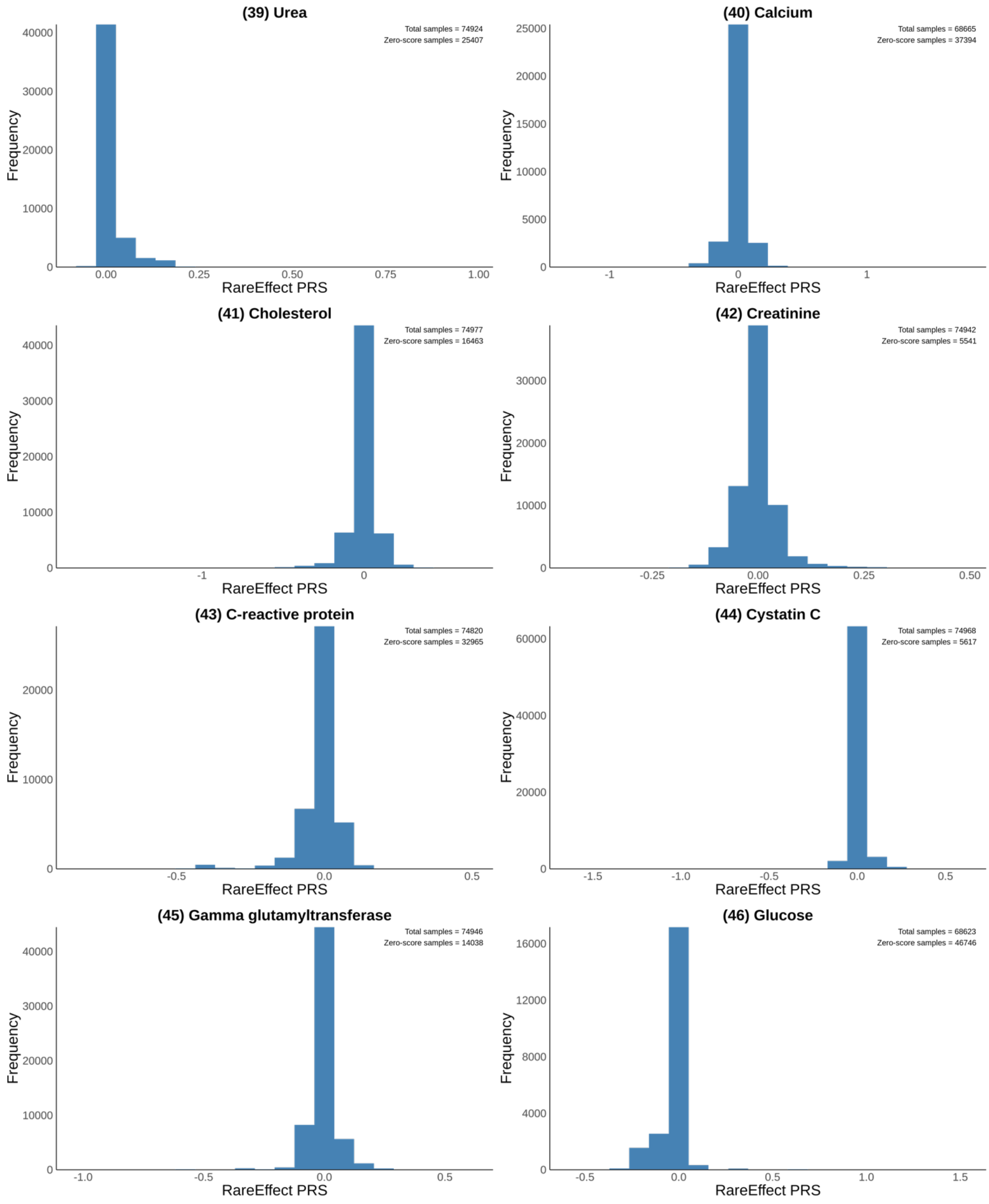

(39) Urea
Total samples = 74924
Zero-score samples = 25407
Frequency
RareEffect PRS
(40) Calcium
Total samples = 68665
Zero-score samples = 37394
Frequency
RareEffect PRS
(41) Cholesterol
Total samples = 74977
Zero-score samples = 16463
Frequency
RareEffect PRS
(42) Creatinine
Total samples = 74942
Zero-score samples = 5541
Frequency
RareEffect PRS
(43) C-reactive protein
Total samples = 74820
Zero-score samples = 32965
Frequency
RareEffect PRS
(44) Cystatin C
Total samples = 74968
Zero-score samples = 5617
Frequency
RareEffect PRS
(45) Gamma glutamyltransferase
Total samples = 74946
Zero-score samples = 14038
Frequency
RareEffect PRS
(46) Glucose
Total samples = 68623
Zero-score samples = 46746
Frequency
RareEffect PRS

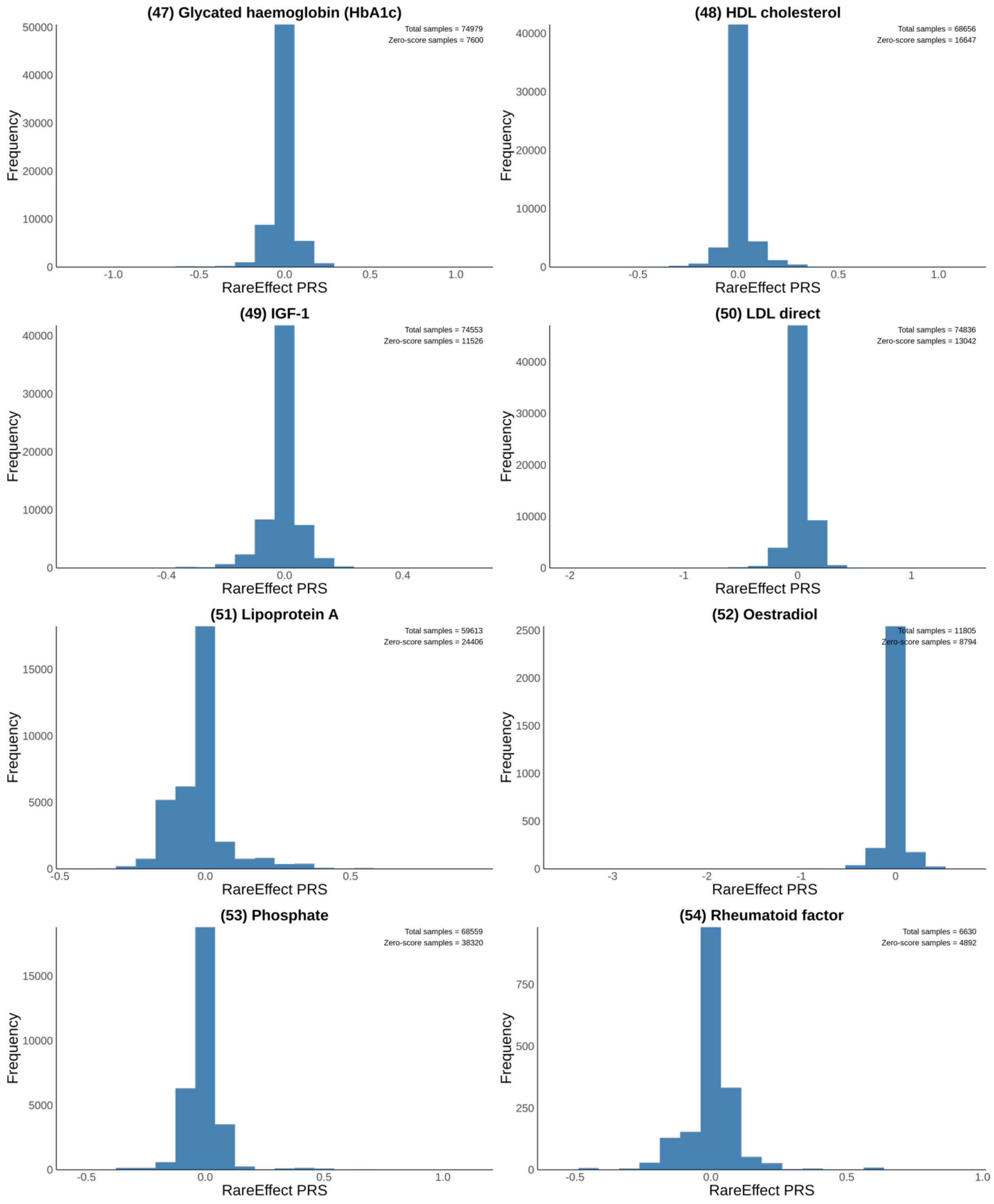
(47) Glycated haemoglobin (HbA1c)
Total samples = 74979
Zero-score samples = 7600
(48) HDL cholesterol
Total samples = 68656
Zero-score samples = 16647
(49) IGF-1
Total samples = 74553
Zero-score samples = 11526
(50) LDL direct
Total samples = 74836
Zero-score samples = 13042
(51) Lipoprotein A
Total samples = 59613
Zero-score samples = 24406
(52) Oestradiol
Total samples = 11805
Zero-score samples = 8794
(53) Phosphate
Total samples = 68559
Zero-score samples = 38320
(54) Rheumatoid factor
Total samples = 6630
Zero-score samples = 4892
Frequency
RareEffect PRS

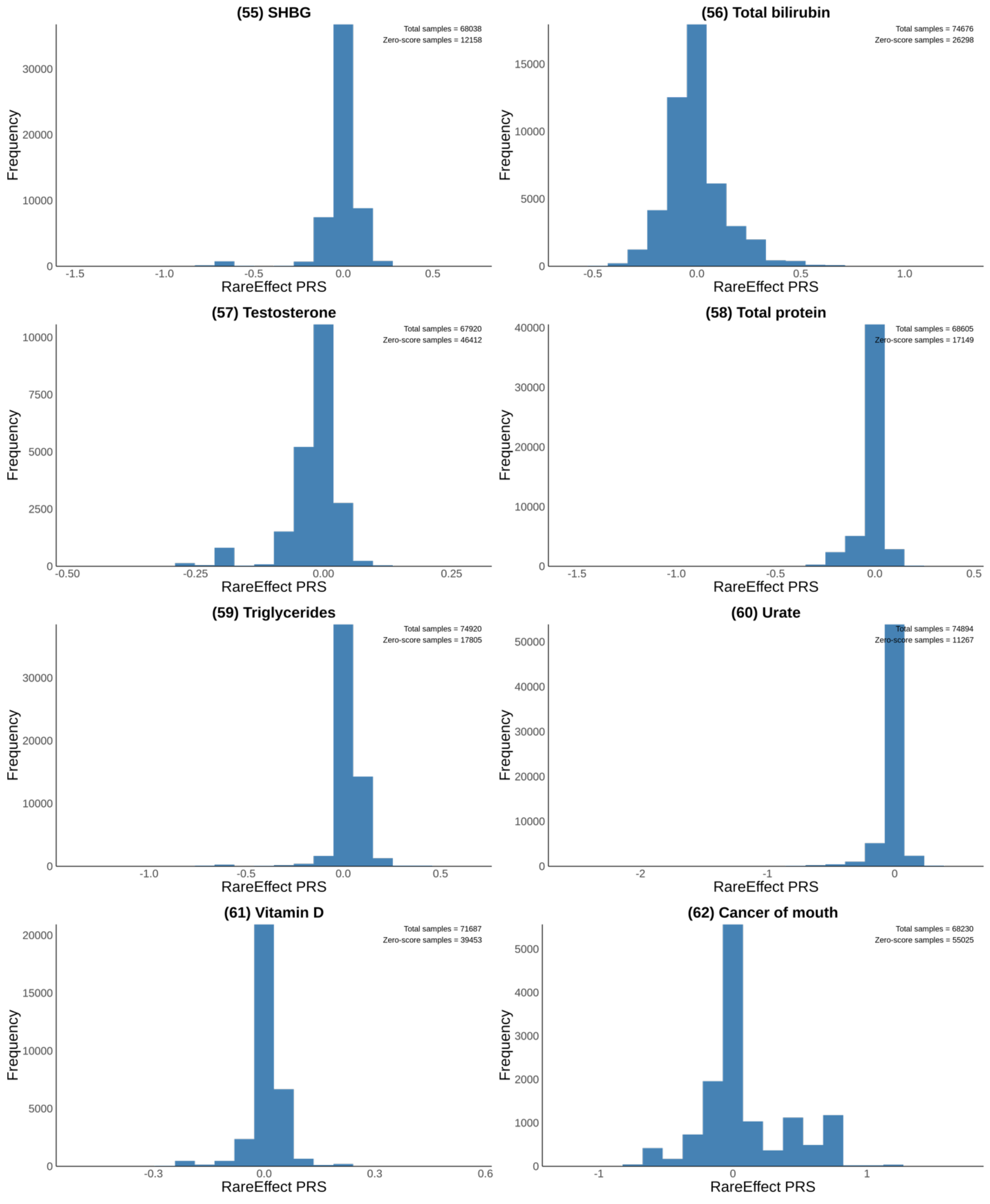
(55) SHBG
Total samples = 68038
Zero-score samples = 12158
Frequency
RareEffect PRS
(56) Total bilirubin
Total samples = 74676
Zero-score samples = 26298
Frequency
RareEffect PRS
(57) Testosterone
Total samples = 67920
Zero-score samples = 46412
Frequency
RareEffect PRS
(58) Total protein
Total samples = 68605
Zero-score samples = 17149
Frequency
RareEffect PRS
(59) Triglycerides
Total samples = 74920
Zero-score samples = 17805
Frequency
RareEffect PRS
(60) Urate
Total samples = 74894
Zero-score samples = 11267
Frequency
RareEffect PRS
(61) Vitamin D
Total samples = 71687
Zero-score samples = 39453
Frequency
RareEffect PRS
(62) Cancer of mouth
Total samples = 68230
Zero-score samples = 55025
Frequency
RareEffect PRS

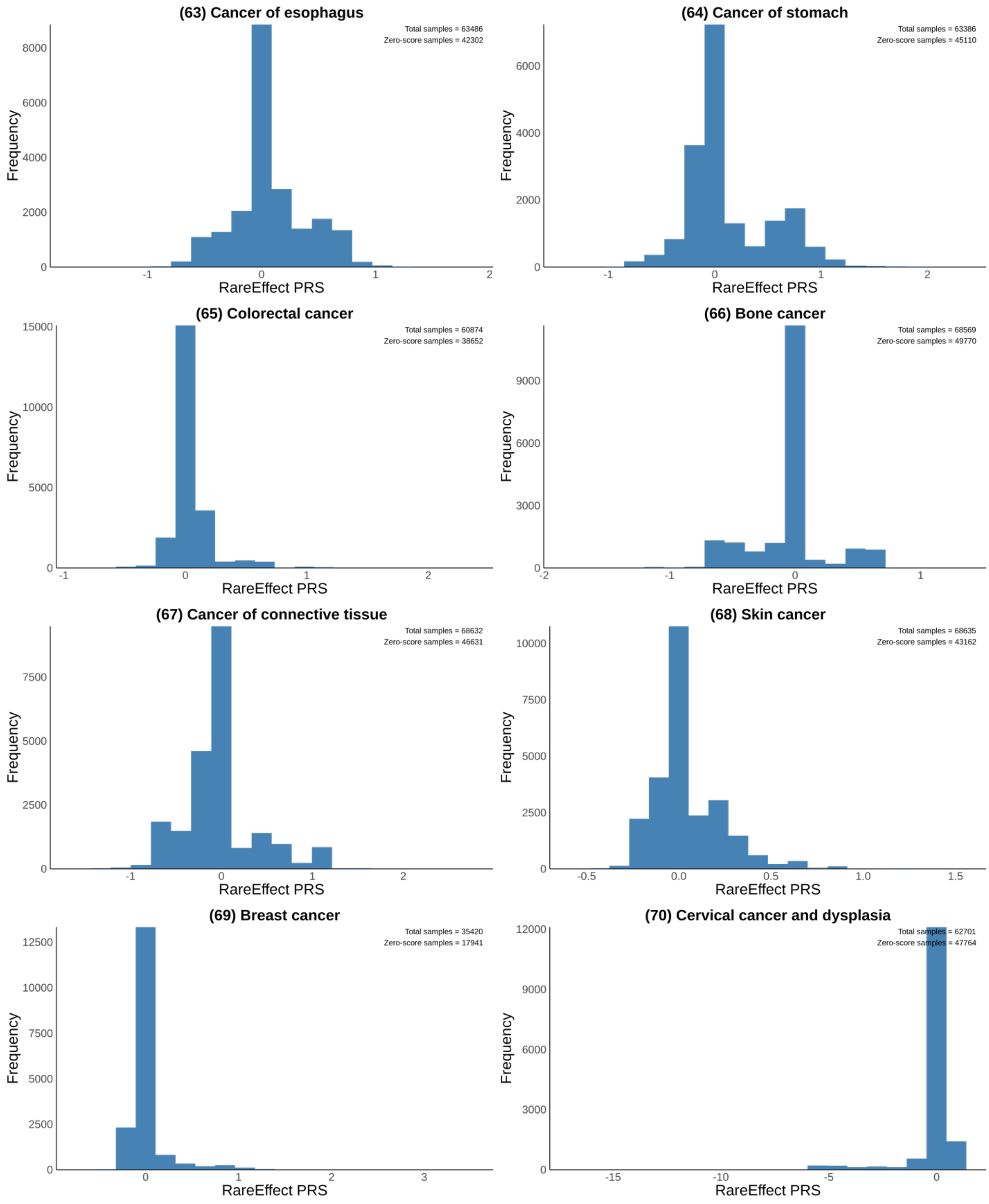
(63) Cancer of esophagus
Total samples = 63486
Zero-score samples = 42302
(64) Cancer of stomach
Total samples = 63386
Zero-score samples = 45110
(65) Colorectal cancer
Total samples = 60874
Zero-score samples = 38652
(66) Bone cancer
Total samples = 68569
Zero-score samples = 49770
(67) Cancer of connective tissue
Total samples = 68632
Zero-score samples = 46631
(68) Skin cancer
Total samples = 68635
Zero-score samples = 43162
(69) Breast cancer
Total samples = 35420
Zero-score samples = 17941
(70) Cervical cancer and dysplasia
Total samples = 62701
Zero-score samples = 47764
Frequency
RareEffect PRS

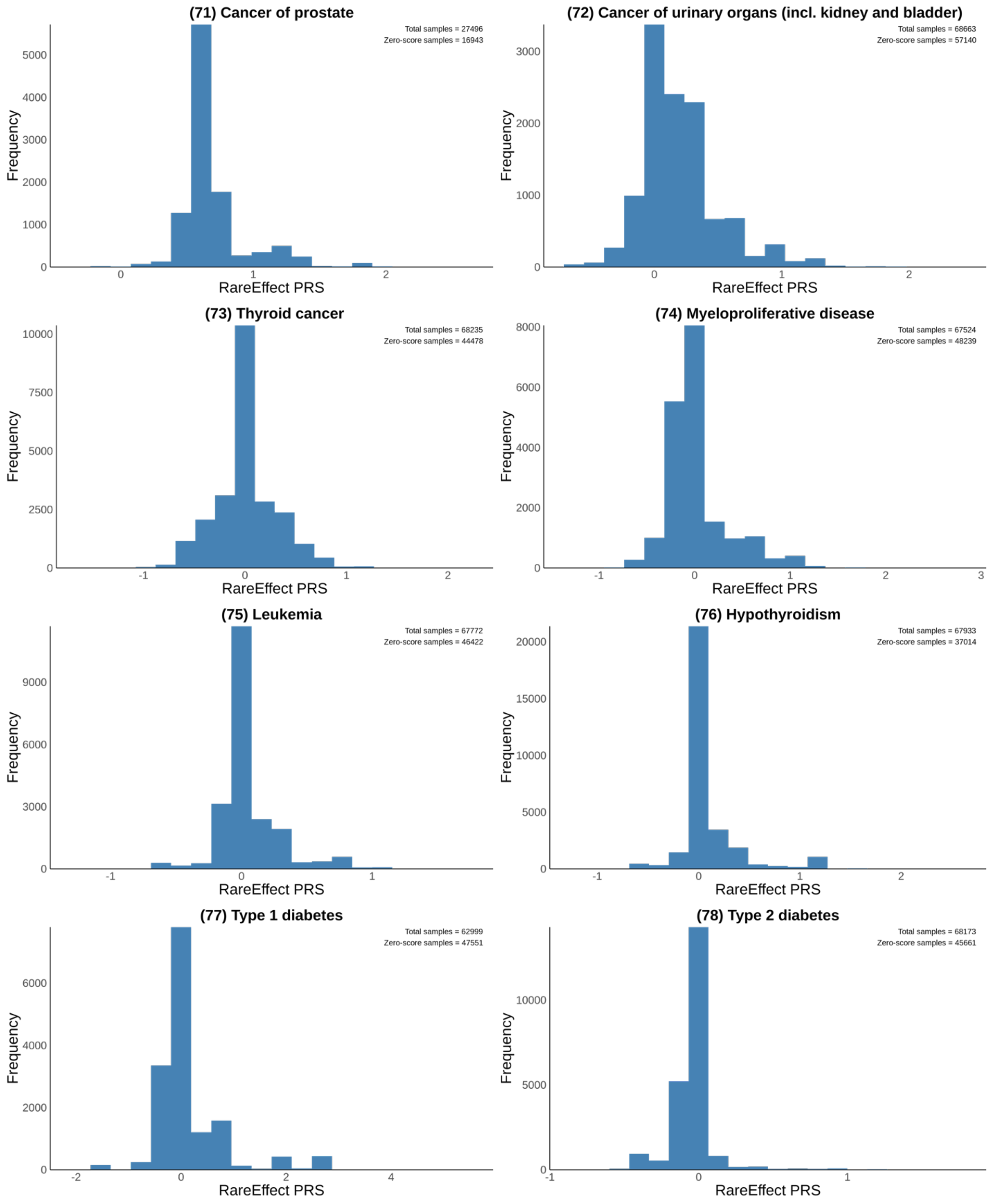
(71) Cancer of prostate
Total samples = 27496
Zero-score samples = 16943
Frequency
RareEffect PRS
(72) Cancer of urinary organs (incl. kidney and bladder)
Total samples = 68663
Zero-score samples = 57140
Frequency
RareEffect PRS
(73) Thyroid cancer
Total samples = 68235
Zero-score samples = 44478
Frequency
RareEffect PRS
(74) Myeloproliferative disease
Total samples = 67524
Zero-score samples = 48239
Frequency
RareEffect PRS
(75) Leukemia
Total samples = 67772
Zero-score samples = 46422
Frequency
RareEffect PRS
(76) Hypothyroidism
Total samples = 67933
Zero-score samples = 37014
Frequency
RareEffect PRS
(77) Type 1 diabetes
Total samples = 62999
Zero-score samples = 47551
Frequency
RareEffect PRS
(78) Type 2 diabetes
Total samples = 68173
Zero-score samples = 45661
Frequency
RareEffect PRS

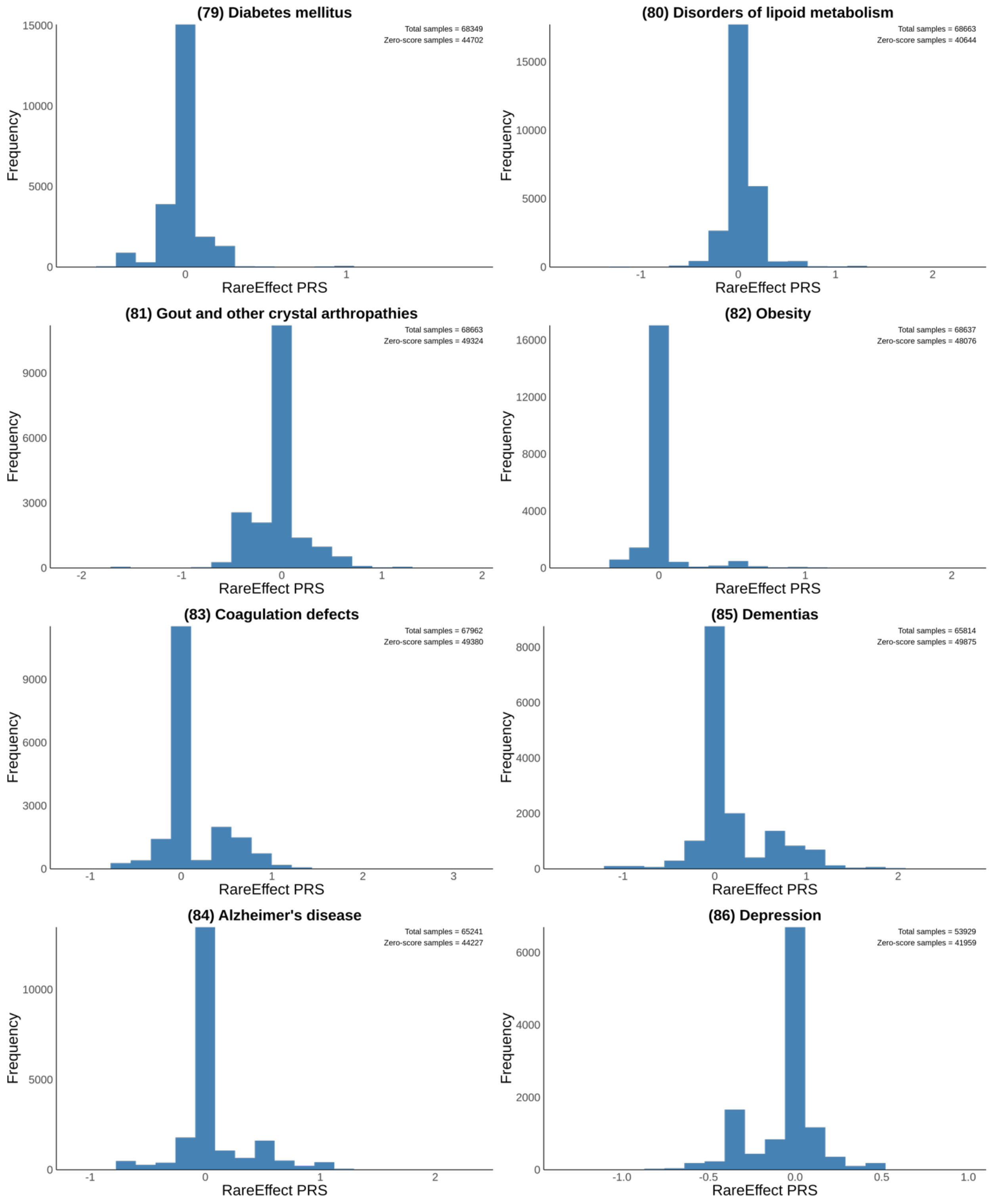

(79) Diabetes mellitus
Total samples = 68349
Zero-score samples = 44702
Frequency
RareEffect PRS
(80) Disorders of lipoid metabolism
Total samples = 68663
Zero-score samples = 40644
Frequency
RareEffect PRS
(81) Gout and other crystal arthropathies
Total samples = 68663
Zero-score samples = 49324
Frequency
RareEffect PRS
(82) Obesity
Total samples = 68637
Zero-score samples = 48076
Frequency
RareEffect PRS
(83) Coagulation defects
Total samples = 67962
Zero-score samples = 49380
Frequency
RareEffect PRS
(85) Dementias
Total samples = 65814
Zero-score samples = 49875
Frequency
RareEffect PRS
(84) Alzheimer's disease
Total samples = 65241
Zero-score samples = 44227
Frequency
RareEffect PRS
(86) Depression
Total samples = 53929
Zero-score samples = 41959
Frequency
RareEffect PRS

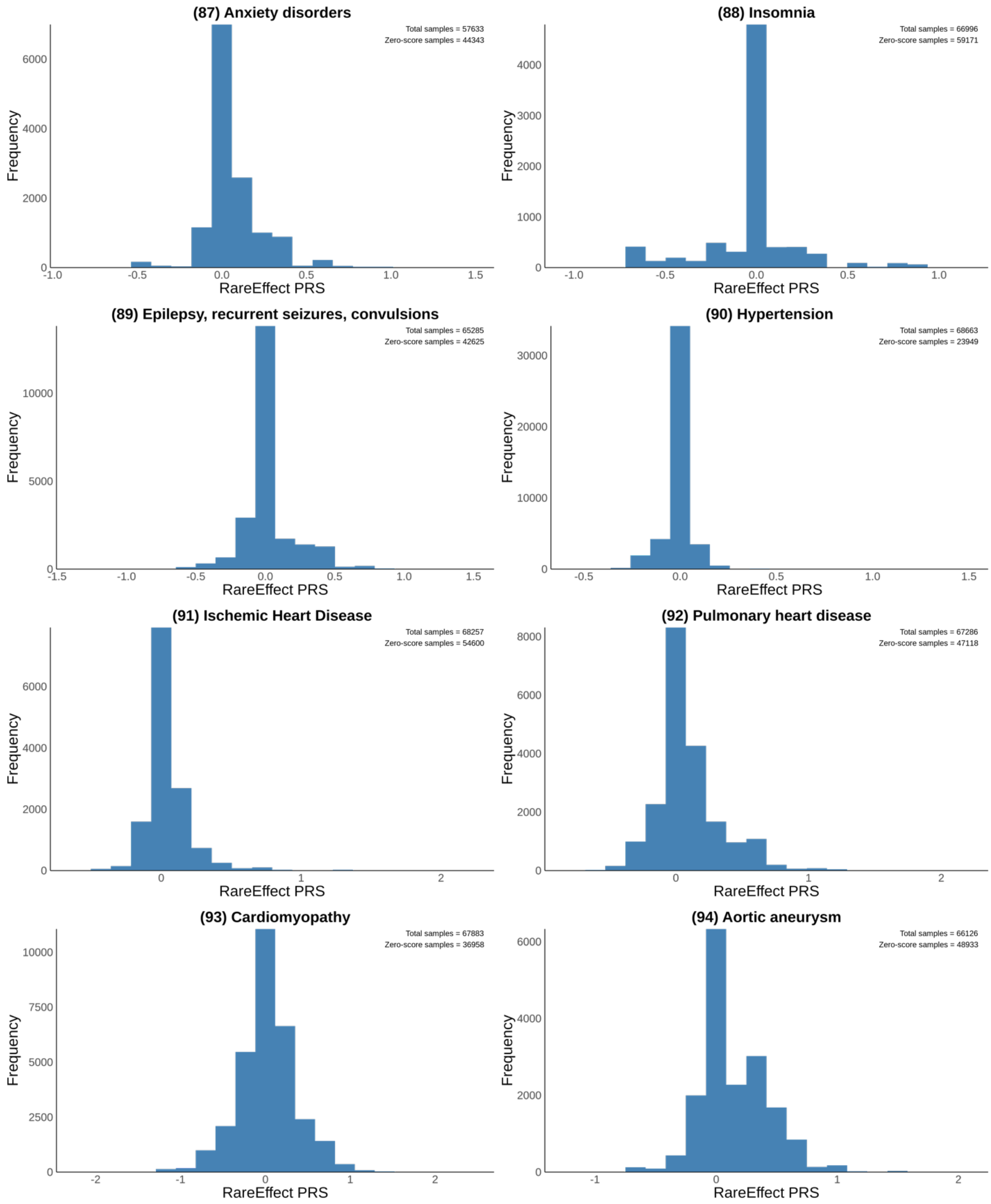
(87) Anxiety disorders
Total samples = 57633
Zero-score samples = 44343
(88) Insomnia
Total samples = 66996
Zero-score samples = 59171
(89) Epilepsy, recurrent seizures, convulsions
Total samples = 65285
Zero-score samples = 42625
(90) Hypertension
Total samples = 68663
Zero-score samples = 23949
(91) Ischemic Heart Disease
Total samples = 68257
Zero-score samples = 54600
(92) Pulmonary heart disease
Total samples = 67286
Zero-score samples = 47118
(93) Cardiomyopathy
Total samples = 67883
Zero-score samples = 36958
(94) Aortic aneurysm
Total samples = 66126
Zero-score samples = 48933
Frequency
RareEffect PRS

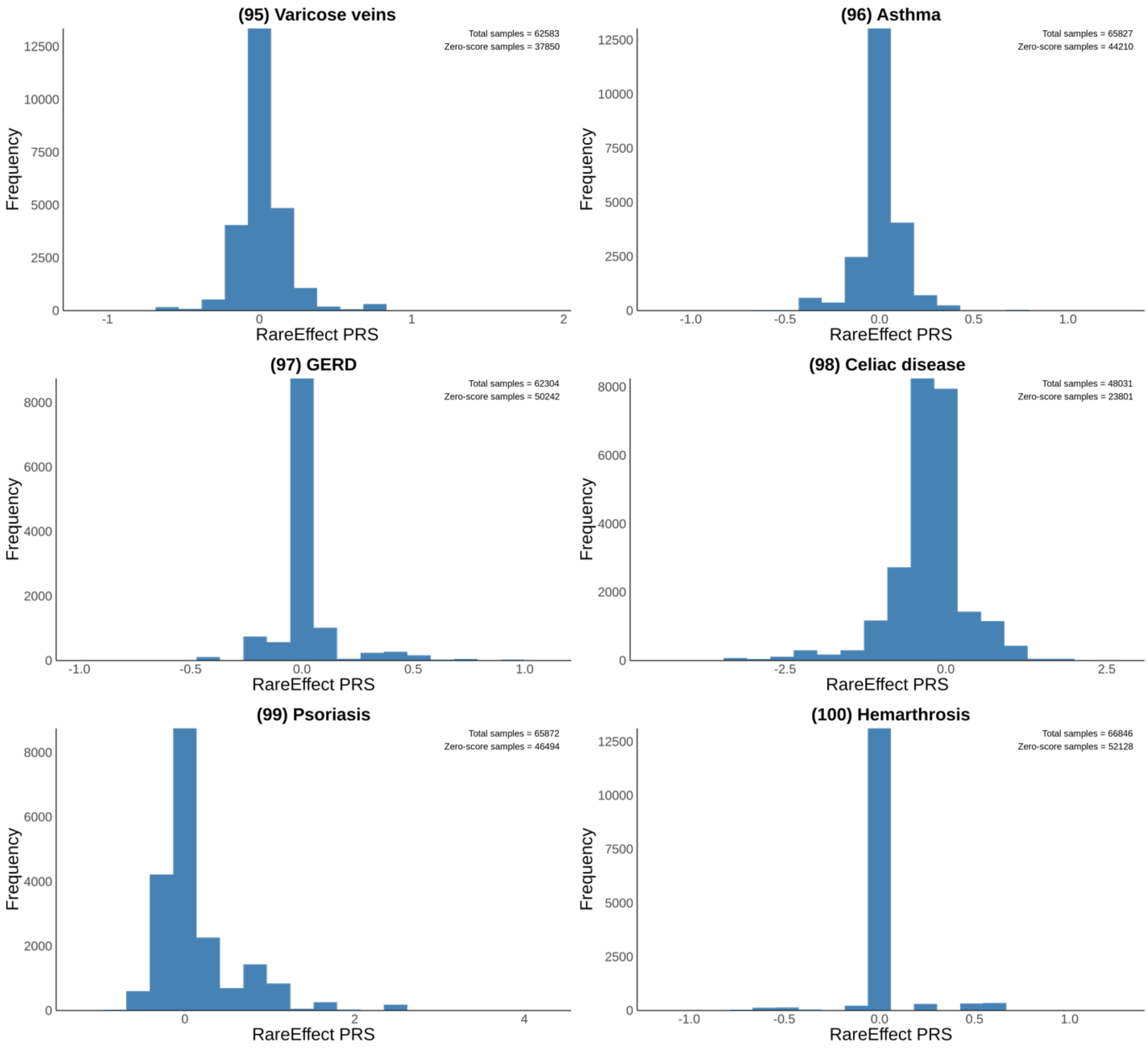
(95) Varicose veins
Total samples = 62583
Zero-score samples = 37850
Frequency
RareEffect PRS
(96) Asthma
Total samples = 65827
Zero-score samples = 44210
Frequency
RareEffect PRS
(97) GERD
Total samples = 62304
Zero-score samples = 50242
Frequency
RareEffect PRS
(98) Celiac disease
Total samples = 48031
Zero-score samples = 23801
Frequency
RareEffect PRS
(99) Psoriasis
Total samples = 65872
Zero-score samples = 46494
Frequency
RareEffect PRS
(100) Hemarthrosis
Total samples = 66846
Zero-score samples = 52128
Frequency
RareEffect PRS
